## Supplementary material for "Spatio-temporal estimates of HIV risk group proportions for adolescent girls and young women across 13 priority countries in sub-Saharan Africa": S1 Text

**References** **12**

### 1 Modelling of risk group proportions

#### 1.1 Overview

Let $i \in \{1, \dots, n\}$ denote subnational units (Table E in S2 Text) which partition the 13 studied AGYW priority countries $c[i] \in \{1, \dots, 13\}$. We considered the years 1999-2018 denoted as $t \in \{1, \dots, T\}$, and age groups $a \in \{15\text{–}19, 20\text{–}24, 25\text{–}29\}$. Let the four risk groups be $k \in \{1, 2, 3, 4\}$ and denote being in either the third or fourth risk group by $k = 3^+$.

First, we used a multinomial logistic regression model to infer the proportion of AGYW in the risk groups $k \in \{1, 2, 3^+\}$. One way to specify this model is using a multinomial likelihood

$$\mathbf{y}_{ita} = (y_{ita1}, \dots, y_{ita3^+})^\top \sim \text{Multinomial}(m_{ita}; p_{ita1}, \dots, p_{ita3^+}), \tag{1}$$

where the number of women in risk group $k$ is $y_{itak}$, the sample size is $m_{ita} = \sum_{k=1}^{3^+} y_{itak}$, $p_{itak} > 0$ is the probability of membership of the $k$th risk group with $\sum_{k=1}^{3^+} p_{itak} = 1$, and taking $k = 1$ to be the baseline category, linear predictors may be specified for the additive log ratios $\log(p_{itak}/p_{ita1})$. To facilitate inference in the `R-INLA`, we used an equivalent Poisson log-linear model via the multinomial-Poisson transformation (Baker 1994). This transformation, and further details about this model, are presented in Section 1.2.

Next, we fit a logistic regression model to estimate the proportion of those in the $k = 3^+$ risk group that were in the $k = 3$ and $k = 4$ risk groups respectively. This model was of the form

$$y_{ia4} \sim \text{Binomial}\left(y_{ia3} + y_{ia4}, q_{ia}\right), \tag{2}$$

$$q_{ia} = \text{logit}^{-1}\left(\eta_{ia}\right), \tag{3}$$

where $q_{ia} = p_{ia4}/(p_{ia3} + p_{ia4}) = p_{ia4}/p_{ia3^+}$ and the linear predictor $\eta_{ia}$ is chosen suitably, described in more detail in Section 1.3. Taking this two-step approach allowed us to include all surveys in the multinomial regression model, but only those surveys with a specific transactional sex question in Equation 2. As all such surveys occurred in 2013-2018, in the logistic regression model we assumed $q_{ia}$ to be constant over time.

To facilitate uncertainty quantification, we took 1000 posterior samples, indexed by $s$, from each of the multinomial $\{p^s_{itak}\}$ and logistic regression $\{q^s_{ia}\}$ models. Samples $p^s_{ita3}$ and $p^s_{ita4}$ were then generated by $p^s_{ita3} = (1 - q^s_{ia})p_{ita3^{+s}}$ and $p^s_{ita4} = q^s_{ia}p^s_{ita3^+}$.

As transactional sex does not directly correspond to sex work (Wamoyi et al. 2016), we adjusted our samples so that at a national level, the population size estimates match FSW population size estimates by age. Our methodology for producing these estimates is described in Section 2. In making this adjustment, we assumed that subnational variation in the FSW proportions corresponds to that of the transactional sex proportions.

We calculated the mean, median, and 2.5% and 97.5% quantiles for the risk group probabilities empirically using the adjusted samples. To produce aggregated estimates, such as the age category 15-24 or any national estimates, we weighted the adjusted samples by population sizes $N_{ita} = \sum_k N_{itak}$ obtained from the Naomi model (Eaton et al. 2021).

#### 1.2 Multinomial regression model

##### 1.2.1 The multinomial-Poisson transformation

The multinomial-Poisson transformation reframes a given multinomial logisitic regression model as an equivalent Poisson log-linear model of the form

$$y_{itak} \sim \text{Poisson}(\kappa_{itak}), \tag{4}$$

$$\log(\kappa_{itak}) = \eta_{itak}, \tag{5}$$

for certain choice of the linear predictor $\eta_{itak}$. The basis of the transformation is that, conditional on their sum, Poisson counts are jointly multinomially distributed (McCullagh and Nelder 1989) as follows

$$\mathbf{y}_{ita} \,|\, m_{ita} \sim \text{Multinomial}\left(m_{ita}; \frac{\kappa_{ita1}}{\kappa_{ita}}, \dots, \frac{\kappa_{ita3^+}}{\kappa_{ita}}\right), \tag{6}$$

where $\kappa_{ita} = \sum_{k=1}^{3^+} \kappa_{itak}$ such that category probabilites are obtained by the softmax funciton

$$p_{itak} = \frac{\exp(\eta_{itak})}{\sum_{k=1}^{3^+} \exp(\eta_{itak})} = \frac{\kappa_{itak}}{\sum_{k=1}^{3^+} \kappa_{itak}} = \frac{\kappa_{itak}}{\kappa_{ita}}. \tag{7}$$

In the equivalent model, the sample sizes $m_{ita} = \sum_k y_{itak}$ are treated as random, rather than fixed as they would be in the multinomial logistic regression model, taking a Poisson distribution

$$m_{ita} \sim \text{Poisson}\left(\kappa_{ita}\right). \tag{8}$$

In the equivalent model, the joint distribution of $p(\mathbf{y}_{ita}, m_{ita}) = p(\mathbf{y}_{ita} \,|\, m_{ita}) p(m_{ita})$ is

$$p(\mathbf{y}_{ita}, m_{ita}) = \exp(-\kappa_{ita}) \frac{(\kappa_{ita})^{m_{ita}}}{m_{ita}!} \times \frac{m_{ita}!}{\prod_k y_{itak}!} \prod_k \left(\frac{\kappa_{itak}}{\kappa_{ita}}\right)^{y_{itak}} \tag{9}$$

$$= \prod_k \left(\frac{\exp(-\kappa_{itak})\left(\kappa_{itak}\right)^{y_{itak}}}{y_{itak}!}\right) \tag{10}$$

$$= \prod_k \text{Poisson}\left(y_{itak} \,|\, \kappa_{itak}\right). \tag{11}$$

corresponding to the product of independent Poisson likelihoods as in Equation 4. This model, including random sample sizes, is equivalent to the multinomial logistic regression only when these normalisation constants are recovered exactly. To ensure that this is the case, one approach is to include observation-specific random effects $\theta_{ita}$ in the equation for the linear predictor. Multiplying each of $\{\kappa_{itak}\}_{k=1}^{3^+}$ by $\exp(\theta_{ita})$ has no effect on the category probabilities, but does provide the necessary flexibility for $\kappa_{ita}$ to recover $m_{ita}$ exactly. Although in theory an improper prior $\theta_{ita} \propto 1$ should be used, in practise, by keeping $\eta_{ita}$ otherwise small using appropriate constraints, so that arbitrarily large values of $\theta_{ita}$ are not required, it is sufficient (and practically preferable for inference) to instead use a vague prior.

##### 1.2.2 Model specifications

| Model ID | Category ($\beta_k$) | Country ($\zeta_{ck}$) | Age ($\alpha_{ack}$) | Spatial ($\phi_{ik}$) | Temporal ($\gamma_{tk}$) | Spatio-temporal ($\delta itk$) |
|---|---|---|---|---|---|---|
| M1 | IID | IID | IID | IID | IID | Not included |
| M2 | IID | IID | IID | Besag | IID | Not included |
| M3 | IID | IID | IID | IID | AR1 | Not included |
| M4 | IID | IID | IID | Besag | AR1 | Not included |

Table A: The multinomial regression models that we considered. Observation random effects $\theta_{ita}$, included in all models, are omitted from this table.

We considered four models (Table A) for $\eta_{ita}$ of the form

$$\eta_{ita} = \theta_{ita} + \beta_k + \zeta_{c[i]k} + \alpha_{ac[i]k} + \phi_{ik} + \gamma_{tk}.$$

Observation random effects $\theta_{ita} \sim \mathcal{N}(0, 1000^2)$ were included in all models we considered. To capture country-specific proportion estimates for each category, we included category random effects $\beta_k \sim \mathcal{N}(0, \tau_\beta^{-1})$ and country-category random effects $\zeta_{ck} \sim \mathcal{N}(0, \tau_\zeta^{-1})$. Heterogeneity in risk group proportions by age was allowed by including age-country-category random effects $\alpha_{ack} \sim \mathcal{N}(0, \tau_\alpha^{-1})$. We considered two specifications, independent and identically distributed (IID) and Besag (Besag, York, and Mollié 1991), for the space-category $\phi_{ik}$ random effects (Section 1.2.3) and two specifications, IID and first order autoregressive (AR1), for the year-category $\gamma_{tk}$ random effects (Section 1.2.4). All random effect precision parameters $\tau \in \{\tau_\beta, \tau_\zeta, \tau_\alpha, \tau_\phi, \tau_\gamma\}$ were given independent penalised complexity (PC) priors (Simpson et al. 2017) with base model $\sigma = 0$ given by $p(\tau) = 0.5\nu\tau^{-3/2} \exp\left(-\nu\tau^{-1/2}\right)$ where $\nu = -\ln(0.01)/2.5$ such that $\mathbb{P}(\sigma > 2.5) = 0.01$.

##### 1.2.3 Spatial random effects

The specifications we considered were IID

$$\phi_{ik} \sim \mathcal{N}(0, \tau_\phi^{-1}),$$

and Besag grouped by category

$$\boldsymbol{\phi} = (\phi_{11}, \ldots, \phi_{n1}, \ldots, \phi_{13^+}, \ldots \phi_{n3^+})^\top \sim \mathcal{N}(\mathbf{0}, (\tau_\phi \mathbf{R}_\phi^\star)^-),$$

where the scaled structure matrix $\mathbf{R}_\phi^\star = \mathbf{R}_b^\star \otimes \mathbf{I}$ is given by the Kronecker product of the scaled Besag structure matrix $\mathbf{R}_b^\star$ and the identity matrix $\mathbf{I}$, and $-$ denotes the generalised matrix inverse. Scaling of the structure matrix to have generalised variance one ensures interpretable priors may be placed on the precision parameter (Sørbye and Rue 2014). We followed the further recommendations of Freni-Sterrantino, Ventrucci, and Rue (2018) with regard to disconnected adjacency graphs, singletons and constraints. The Besag structure matrix $\mathbf{R}_b$ is obtained by the precision matrix of the random effects $\mathbf{b} = (b_1, \ldots, b_n)^\top$ with full conditionals

$$b_i \,|\, \mathbf{b}_{-i} \sim \mathcal{N}\left(\frac{\sum_{j:j\sim i} b_j}{n_{\delta i}}, \frac{1}{n_{\delta i}}\right), \tag{12}$$

where $j \sim i$ if the districts $A_i$ and $A_j$ are adjacent, and $n_{\delta i}$ is the number of districts adjacent to $A_i$.

In preliminary testing, we excluded spatial random effects from the model, but found that this negatively effected performance. We also tested using the BYM2 model (Simpson et al. 2017) in place of the Besag, but found that the proportion parameter posteriors tended to be highly peaked at the value one. For simplicity and to avoid numerical issues, by using Besag random effects we decided to fix this proportion to one.

##### 1.2.4 Temporal random effects

The specifications we considered were IID

$$\phi_{tk} \sim \mathcal{N}(0, \tau_\phi^{-1}),$$

and AR1 grouped by category

$$\boldsymbol{\gamma} = (\gamma_{11}, \ldots, \gamma_{13^+}, \ldots, \gamma_{T1}, \ldots, \gamma_{T3^+})^\top \sim \mathcal{N}(\mathbf{0}, (\tau_\phi \mathbf{R}_\gamma^\star)^-),$$

where the scaled structure matrix $\mathbf{R}_\gamma^\star = \mathbf{R}_r^\star \otimes \mathbf{I}$ is given by the Kronecker product of a scaled AR1 structure matrix $\mathbf{R}_r^\star$ and the identity matrix $\mathbf{I}$. The AR1 structure matrix $\mathbf{R}_r$ is obtained by precision matrix of the random effects $\mathbf{r} = (r_1, \ldots, r_T)^\top$ specified by

$$r_1 \sim \left(0, \frac{1}{1-\rho^2}\right), \tag{13}$$

$$r_t = \rho r_{t-1} + \epsilon_t, \quad t = 2, \ldots, T, \tag{14}$$

where $\epsilon_t \sim \mathcal{N}(0,1)$ and $|\rho| < 1$. For the lag-one correlation parameter $\rho$, we used the PC prior, as derived by Sørbye and Rue (2017), with base model $\rho = 1$ and condition $\mathbb{P}(\rho > 0 = 0.75)$. We chose the base model $\rho = 1$ corresponding to no change in behaviour over time, rather than the alternative $\rho = 0$ corresponding to no correlation in behaviour over time, as we judged the former to be more plausible a priori.

##### 1.2.5 Constraints

To ensure interpretable posterior inferences of random effect contribution, we applied sum-to-zero constraints such that none of the category interaction random effects altered overall category probabilities. For the space-year-category random effects, we applied analogous sum-to-zero constraints to maintain roles of the space-category and year-category random effects. Together, these were:

1. Category $\sum_k \beta_k = 0$
2. Country $\sum_c \zeta_{ck} = 0, \forall\, k$
3. Age-country $\sum_a \alpha_{ack} = 0, \forall\, c, k,$
4. Spatial $\sum_i \phi_{ik} = 0, \forall\, k$
5. Temporal $\sum_t \gamma_{tk} = 0, \forall\, k$

##### 1.2.6 Survey weighted likelihood

We included surveys which use a complex design, in which each individual has an unequal probability of being included in the sample. For example the DHS often employs a two-stage cluster design, first taking an urban rural stratified sample of ennumeration areas, before selecting households from each ennumeration area using systematic sampling (DHS 2012).

To account for this aspect of survey design, we use a weighted pseudo-likelihood where the observed counts $y$ are replaced by effective counts $y^\star$ calculated using the survey weights $w_j$ of all individuals $j$ in the corresponding strata. We multiplied direct estimates produced using the `survey` package (Lumley 2004) by the Kish effective sample size (Kish 1965)

$$m^\star = \frac{\left(\sum_j w_j\right)^2}{\sum_j {w_j}^2} \tag{15}$$

to obtain $y^\star$. These counts may not be integers, and as such the Poisson likelihood we used in Equation 4 is not appropriate. Instead, we used a generalised Poisson pseudo-likelihood $y^\star \sim \text{xPoisson}(\kappa)$, given by

$$p(y^\star) = \frac{\kappa^{y^\star}}{\lfloor y^\star ! \rfloor} \exp\left(-\kappa\right), \tag{16}$$

as implemented by `family = "xPoisson"` in `R-INLA`, which accepts non-integer input.

##### 1.2.7 Model selection

We performed model selection on the basis of the conditional predictive ordinate (CPO) criterion (Pettit 1990), selecting model M2. This model included Besag spatial random effects and IID temporal random effects. For reference, we also computed the deviance information criterion (DIC) (Spiegelhalter et al. 2002) and widely applicable information criterion (WAIC) (Watanabe 2013) (Table B and Fig A).

| | M1 | M2 | M3 | M4 |
|---|---|---|---|---|
| DIC | 100780 (300) | 101588 (317) | 100781 (300) | 101589 (317) |
| WAIC | 103763 (358) | 105008 (383) | 103763 (358) | 105009 (383) |
| CPO | 5573 (36) | 5772 (36) | 5574 (36) | 5771 (36) |

Table B: Multinomial regression model performance. For the CPO, higher values indicate better model performance. For the DIC and WAIC, lower values indicate better model performance.

#### 1.3 Logistic regression model

| Model ID | Intercept ($\beta_0$) | Country ($\zeta_c$) | Age ($\alpha_c a$) | Spatial ($\phi_i$) | Covariates |
|---|---|---|---|---|---|
| L1 | Constant | IID | IID | IID | Not included |
| L2 | Constant | IID | IID | Besag | Not included |
| L3 | Constant | IID | IID | IID | `cfswever` |
| L4 | Constant | IID | IID | Besag | `cfswever` |
| L5 | Constant | IID | IID | IID | `cfswrecent` |
| L6 | Constant | IID | IID | Besag | `cfswrecent` |

Table C: The logistic regression models that were considered. `cfswever` denotes the proportion of men who have ever paid for sex and `cfswrecent` denotes the proportion of men who have paid for sex in the past 12 months.

We considered six logistic regression models (Table C) each including a constant $\beta_0 \sim \mathcal{N}(-2, 1^2)$, country random effects $\zeta_c \sim \mathcal{N}(0, \tau_\zeta^{-1})$, and age-country random effects $\alpha_{ac} \sim \mathcal{N}(0, \tau_\alpha^{-1})$. The prior on $\beta_0$ placed

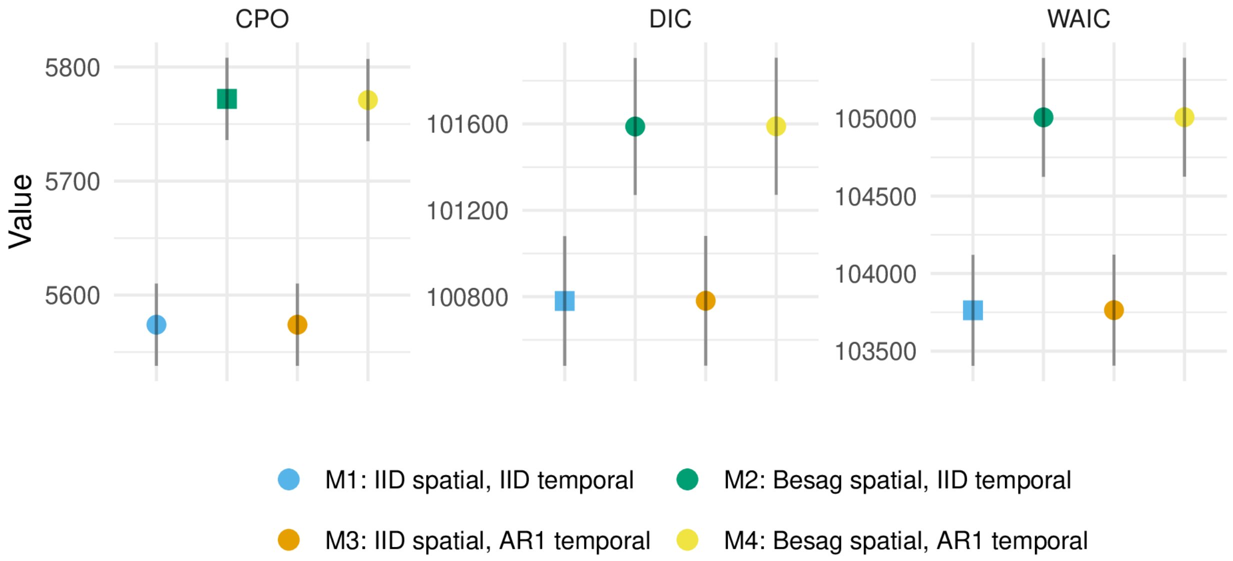

Figure A: Multinomial regression model performance, with the best performing model(s) according to each criterion shown as a square. For the CPO, higher values indicate better model performance for the CPO. For the DIC and WAIC, lower values indicate better model performance.

95% prior probability on the range 2-50% for the percentage of those with non-regular or multiple partners who report transactional sex. We considered two specifications (IID, Besag) for the spatial random effects $\phi_i$. To aid estimation with sparse data, we also considered national-level covariates for the proportion of men who have paid for sex ever `cfswever` or in the last twelve months `cfswrecent`, available from Hodgins et al. (2022). For both random effect precision parameters $\tau \in \{\tau_\alpha, \tau_\zeta\}$ we used the PC prior with base model $\sigma = 0$ and $\mathbb{P}(\sigma > 2.5 = 0.01)$. For the regression parameters $\beta \in \{\beta_{\texttt{cfswever}}, \beta_{\texttt{cfswrecent}}\}$ we used the prior $\beta \sim \mathcal{N}(0, 2.5^2)$.

##### 1.3.1 Survey weighted likelihood

As with the multinomial regression model, we used survey weighted counts $\{y^\star_{itak}\}$ and sample sizes $\{m^\star_{itak}\}$. We used a generalised binomial pseudo-likelihood $y^\star \sim \text{xBinomial}(m^\star, q)$, as implemented by `family = "xBinomial"` in `R-INLA`, given by

$$p(y^\star \mid m^\star, q) = \binom{\lfloor m^\star \rfloor}{\lfloor y^\star \rfloor} q^{y^\star} (1-q)^{m^\star - y^\star}. \tag{17}$$

to extend the binomial distribution to non-integer weighted counts and sample sizes.

##### 1.3.2 Model selection

We selected the best model according to the CPO statistic, which was model L6. CPO values, along with DIC and WAIC values for reference, are presented in Table D and Fig B. Inclusion of Besag spatial random effects, rather than IID, consistently improved performance. Benefits from inclusion of covariates were more marginal. That said, as some countries had no suitable surveys, we preferred to include covariate information such that the estimates in these countries are based on some country-specific data.

#### 1.4 Coverage assessment

To assess the calibration of our fitted model, we calculated the quantile $q$ of each observation within the posterior predictive distribution. For calibrated models, these quantiles, known as probability integral

| | L1 | L2 | L3 | L4 | L5 | L6 |
|---|---|---|---|---|---|---|
| DIC | 4662 (110) | 4605 (111) | 4662 (110) | 4605 (111) | 4662 (110) | 4605 (111) |
| WAIC | 4692 (115) | 4624 (115) | 4692 (115) | 4624 (115) | 4692 (115) | 4624 (115) |
| CPO | 950 (15) | 969 (15) | 951 (15) | 970 (15) | 950 (15) | 970 (15) |

Table D: Logistic regression model performance. For the CPO, higher values indicate better model performance. For the DIC and WAIC, lower values indicate better model performance.

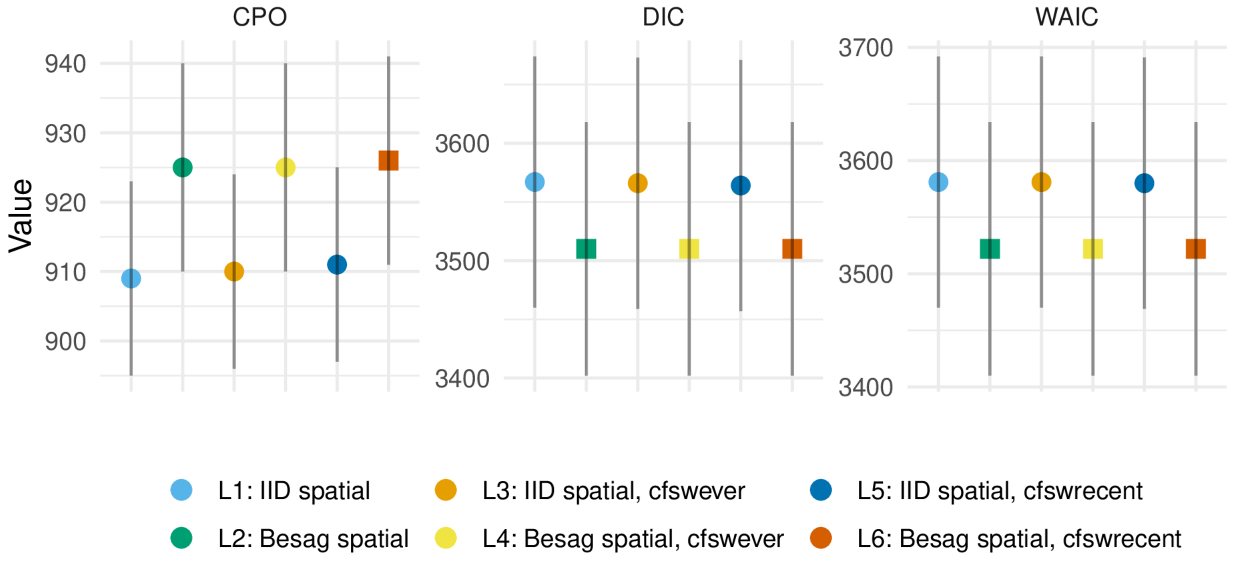

Figure B: Logistic regression model performance, with the best performing model(s) according to each criterion shown as a square.

transform (PIT) values (Dawid 1984; Bosse et al. 2022), should follow a uniform distribution $q \sim \mathcal{U}[0, 1]$. To generate samples from the posterior predictive distribution, we applied the multinomial likelihood to samples from the latent field, setting the sample size to be the floor of the Kish effective sample size.

Using the PIT values, it is possible to calculate the empirical coverage of all $(1 - \alpha)100\%$ (equal-tailed) posterior predictive credible intervals. These empirical coverages can be compared to the nominal coverage $(1 - \alpha)$ for each value of $\alpha \in [0, 1]$ to give empirical cumulative distribution function (ECDF) difference values. This approach has the advantage of considering all possible confidence values at once. Säilynoja, Bürkner, and Vehtari (2021) develop binomial distribution-based simultaneous confidence bands for ECDF difference values which test uniformity.

Fig C shows the PIT histogram and ECDF difference plots for our final model, faceted by risk group.

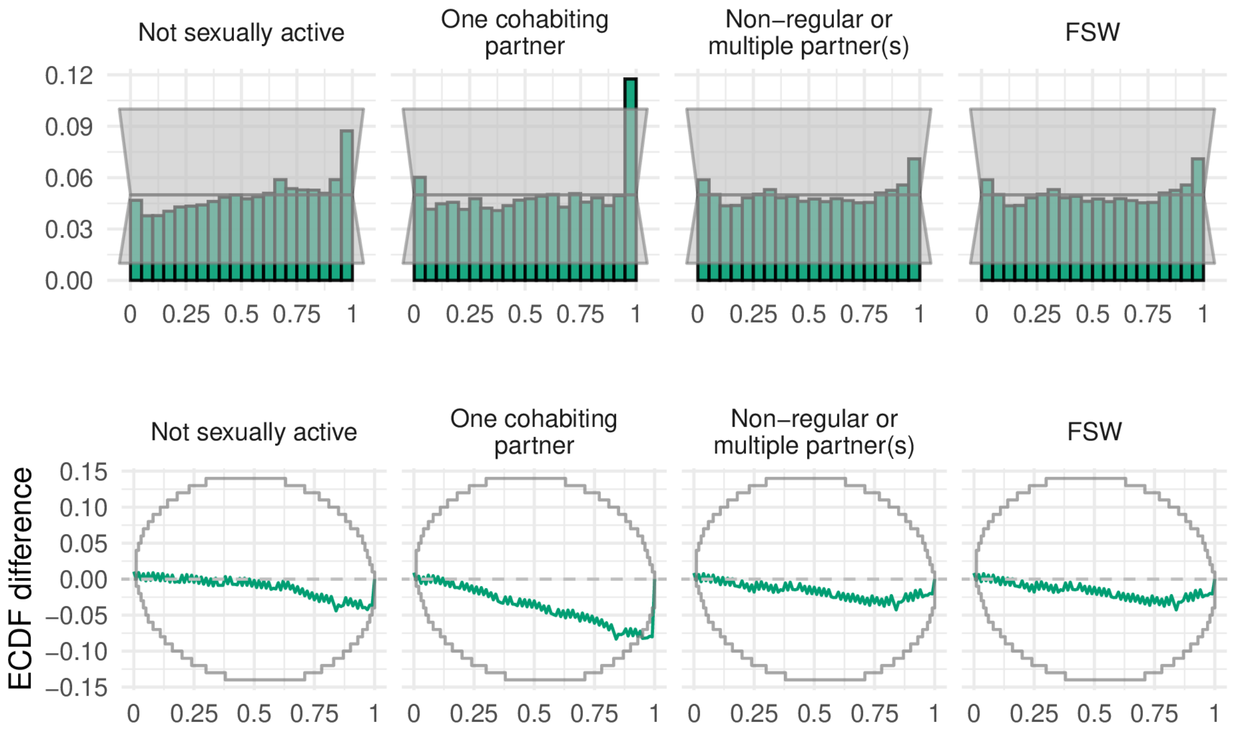

Figure C: Probability integral transform (PIT) histograms (top row) and empirical cumulative distribution function (ECDF) difference plots (bottom row) for the final selected model.

#### 2 FSW population size estimation

To estimate the number of FSW by age group and country, we disaggregated country-specific estimates of adult (15-49) FSW population size from Stevens et al. (2022) by age group.

First, we calculated the total sexually debuted population in each age group, in each country. To describe the distribution of age at first sex, we used skew logistic distributions (Nguyen and Eaton 2022) with cumulative distribution function given by

$$F(x) = (1 + \exp(\kappa_c(\mu_c - x)))^{-\gamma_c}, \tag{18}$$

where $\kappa_c, \mu_c, \gamma_c > 0$ are country-specific shape, shape and skewness parameters respectively.

Next, we used the assumed Gamma($\alpha = 10.4, \beta = 0.36$) FSW age distribution in South Africa from the Thembisa model (Johnson and Dorrington 2020) to calculate the implied ratio between the number of FSW and the sexually debuted population in each age group. We assumed these ratios in South Africa were applicable to every country to calculate the number of FSW by age group in all 13 countries (Fig D).

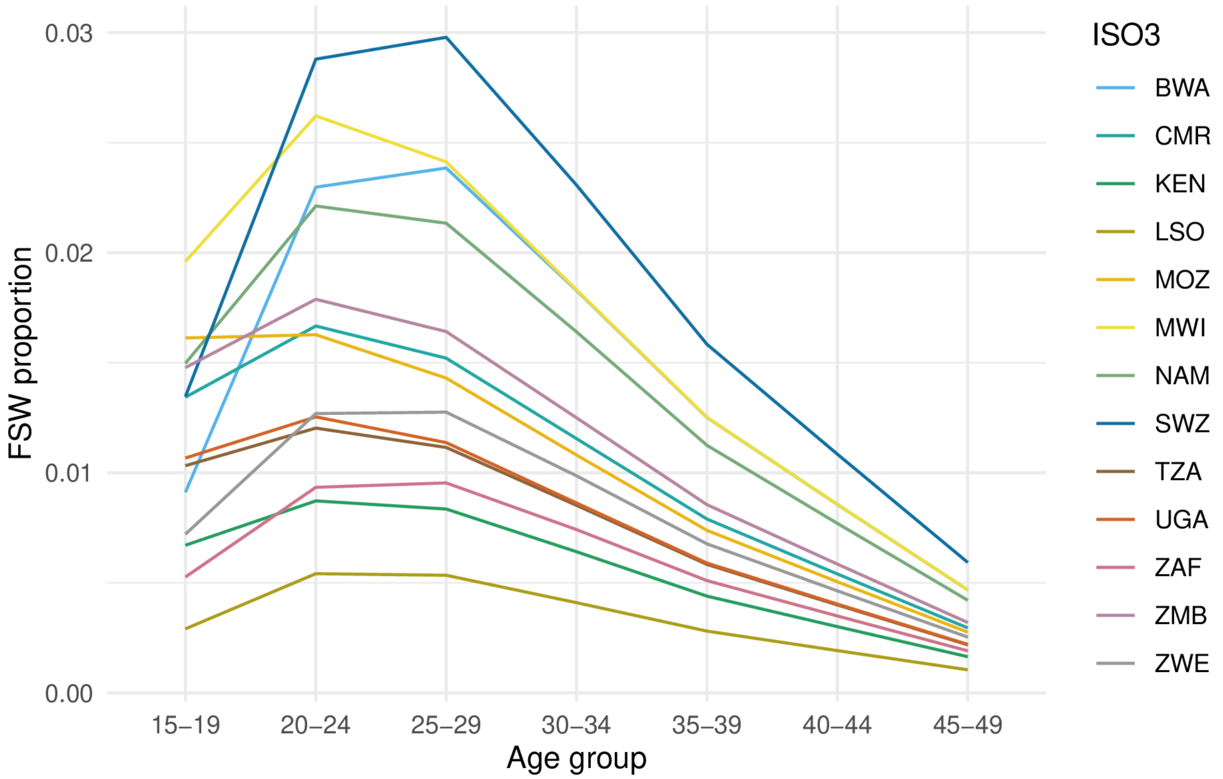

Figure D: Proportion of FSW by age group (including the age groups 30-34, 35-39, 40-44 and 45-49) as produced by our disaggregation procedure.

### 3 Prevalence, incidence and expected new infections reached

#### 3.1 Calculation of prevalence and PLHIV

We calculated HIV prevalence $\rho_{iak}$ and the number of people living with HIV (PLHIV) $H_{iak}$ stratified according to district, age group and risk group by disaggregating Naomi estimates by risk group.

To do so, we estimated HIV prevalence log odds ratios relative to the general population using age, country and risk group specific HIV prevalence bio-marker survey data. We also included general population HIV prevalence data, allowing us to fit a logistic regression model including indicator functions for each risk group, and an indicator for being in the general population. The regression coefficients in this model correspond to log odds, such that log odds ratios may be easily obtained by taking the difference.

To allow the log odds ratio for the highest risk group to vary based on general population prevalence we fit a linear regression of the FSW log odds against the general population log odds. We ensured that the log odds ratio for the highest risk group was at least as large as that of the second highest risk group.

Given the fitted log odds ratios $\log(\text{OR}_k)$, we disaggregated Naomi estimates of PLHIV $H_{ia}$ on the logit scale using numerically optimisation to obtain the value of $\theta$ minimising the function

$$\hat{\theta} = \underset{\theta \in [-10,10]}{\arg\min} \left( \sum_k (\text{logistic}(\theta + \log(\text{OR}_k)) \cdot N_{iak}) - H_{ia} \right)^2 \tag{19}$$

where $\text{logistic}(x) = \exp(x)/(1+\exp(x))$ such that $\text{logistic}(\hat{\theta} + \log(\text{OR}_k)) = \rho_{iak}$. Fig R - AD in S2 Text show the resulting HIV prevalence estimates by risk group in each country. The number of PLHIV are then obtained by $H_{iak} = \rho_{iak} N_{iak}$, where $N_{iak}$ is the risk group population size.

#### 3.2 Calculation of incidence and expected number of new infections

We calculated HIV incidence $\lambda_{iak}$ and number of new HIV infections $I_{iak}$ stratified according to district, age group and risk group by linear disaggregation

$$I_{ia} = \sum_k I_{iak} = \sum_k \lambda_{iak} N_{iak} \tag{20}$$

$$= 0 + \lambda_{ia2} N_{ia2} + \lambda_{ia3} N_{ia3} + \lambda_{ia4} N_{ia4} \tag{21}$$

$$= \lambda_{ia2} \left( N_{ia2} + \text{RR}_3 N_{ia3} + \text{RR}_4(\lambda_{ia}) N_{ia4} \right). \tag{22}$$

Risk group specific HIV incidence estimates are then given by

$$\lambda_{ia1} = 0, \tag{23}$$

$$\lambda_{ia2} = I_{ia} / \left( N_{ia2} + \text{RR}_3 N_{ia3} + \text{RR}_4(\lambda_{ia}) N_{ia4} \right), \tag{24}$$

$$\lambda_{ia3} = \text{RR}_3 \lambda_{ia2}, \tag{25}$$

$$\lambda_{ia4} = \text{RR}_4(\lambda_{ia}) \lambda_{ia2}. \tag{26}$$

which we evaluated using Naomi model estimates of the number of new HIV infections $I_{ia} = \lambda_{ia} N_{ia}$, HIV infection risk ratios $\{\text{RR}_3, \text{RR}_4(\lambda_{ia})\}$, and risk group population sizes as above. The risk ratio $\text{RR}_4(\lambda_{ia})$ was defined as a function of general population incidence. Fig AE - AQ in S2 Text show the resulting HIV incidence estimates by risk group in each country. The number of new HIV infections are then $I_{iak} = \lambda_{iak} N_{iak}$.

#### 3.3 Calculation of expected new infections reached

We calculated the number of new infections that would be reached prioritising according to each possible stratification of the population–that is for all $2^3 = 8$ possible combinations of stratification by location, age, and risk group. As an illustration, for stratification just by age, we aggregated the number of new HIV infections and HIV incidence as such

$$I_a = \sum_{ik} I_{iak}, \tag{27}$$

$$\lambda_a = I_a / \sum_{ik} N_{iak}. \tag{28}$$

Under this stratification, individuals in each age group $a$ are prioritised according to the highest HIV incidence $\lambda_a$. By cumulatively summing the expected infections, for each fraction of the total population reached we calculated the fraction of total expected new infections that would be reached. Fig AR - BD in S2 Text show the percentage of new infections that would be reached prioritising according to each possible stratification within each country.

This analysis was relatively simple. More involved analyses might consider prioritisation of a hypothetical intervention which has some, possibly varying, probability of preventing HIV acquisition, as well as the costs associated to its roll-out.

DHS. 2012. "Sampling and Household Listing Manual: Demographic and Health Surveys Methodology."

Eaton, Jeffrey W., Laura Dwyer-Lindgren, Steve Gutreuter, Megan O'Driscoll, Oliver Stevens, Sumali Bajaj, Rob Ashton, et al. 2021. "Naomi: a new modelling tool for estimating HIV epidemic indicators at the district level in sub-Saharan Africa." *Journal of the International AIDS Society* 24 (S5): e25788.

Freni-Sterrantino, Anna, Massimo Ventrucci, and Håvard Rue. 2018. "A Note on Intrinsic Conditional Autoregressive Models for Disconnected Graphs." *Spatial and Spatio-Temporal Epidemiology* 26: 25–34.

Hodgins, Caroline, James Stannah, Salome Kuchukhidze, Lycias Zembe, Jeffrey W Eaton, Marie-Claude Boily, and Mathieu Maheu-Giroux. 2022. "Population Sizes, HIV Prevalence, and HIV Prevention Among Men Who Paid for Sex in Sub-Saharan Africa (2000–2020): A Meta-Analysis of 87 Population-Based Surveys." *PLoS Medicine* 19 (1): e1003861.

Johnson, L, and RE Dorrington. 2020. "Thembisa version 4.3: A model for evaluating the impact of HIV/AIDS in South Africa." *View Article*.

Kish, Leslie. 1965. *Survey Sampling*. 04; HN29, K5.

Lumley, Thomas. 2004. "Analysis of Complex Survey Samples." *Journal of Statistical Software, Articles* 9 (8): 1–19. https://doi.org/10.18637/jss.v009.i08.

McCullagh, Peter, and John A Nelder. 1989. *Generalized Linear Models*. Routledge.

Nguyen, Van Kính, and Jeffrey W. Eaton. 2022. "Trends and country-level variation in age at first sex in sub-Saharan Africa among birth cohorts entering adulthood between 1985 and 2020." *BMC Public Health* 22 (1): 1120. https://doi.org/10.1186/s12889-022-13451-y.

Pettit, LI. 1990. "The Conditional Predictive Ordinate for the Normal Distribution." *Journal of the Royal Statistical Society: Series B (Methodological)* 52 (1): 175–84.

Säilynoja, Teemu, Paul-Christian Bürkner, and Aki Vehtari. 2021. "Graphical Test for Discrete Uniformity and Its Applications in Goodness of Fit Evaluation and Multiple Sample Comparison." *arXiv Preprint arXiv:2103.10522*.

Simpson, Daniel, Håvard Rue, Andrea Riebler, Thiago G Martins, and Sigrunn H Sørbye. 2017. "Penalising Model Component Complexity: A Principled, Practical Approach to Constructing Priors." *Statistical Science* 32 (1): 1–28.

Sørbye, Sigrunn Holbek, and Håvard Rue. 2014. "Scaling intrinsic Gaussian Markov random field priors in spatial modelling." *Spatial Statistics* 8: 39–51.

———. 2017. "Penalised Complexity Priors for Stationary Autoregressive Processes." *Journal of Time Series Analysis* 38 (6): 923–35.

Spiegelhalter, David J, Nicola G Best, Bradley P Carlin, and Angelika Van Der Linde. 2002. "Bayesian measures of model complexity and fit." *Journal of the Royal Statistical Society: Series B (Statistical Methodology)* 64 (4): 583–639.

Stevens, Oliver, Keith Sabin, Sonia Arias Garcia, Kalai Willis, Abu Abdul-Quader, Anne McIntyre, Frances Cowan, et al. 2022. "Estimating key population size, HIV prevalence, and ART coverage for sub-Saharan Africa at the national level."

Wamoyi, Joyce, Kirsten Stobeanau, Natalia Bobrova, Tanya Abramsky, and Charlotte Watts. 2016. "Transactional sex and risk for HIV infection in sub-Saharan Africa: a systematic review and meta-analysis." *Journal of the International AIDS Society* 19 (1): 20992.

Watanabe, Sumio. 2013. "A widely applicable Bayesian information criterion." *Journal of Machine Learning Research* 14 (Mar): 867–97.
