## Supplementary material for "Spatio-temporal estimates of HIV risk group proportions for adolescent girls and young women across 13 priority countries in sub-Saharan Africa": S2 Text

### The Global AIDS Strategy

| Prioritisation strata | Criterion |
|---|---|
| Low | 0.3-1.0% incidence and low-risk behaviour, or <0.3% incidence and high-risk behaviour |
| Moderate | 1.0-3.0% incidence and low-risk behaviour, or 0.3-1.0% incidence and high-risk behaviour |
| High | 1.0-3.0% incidence and high-risk behaviour |
| Very high | >3.0% incidence |

Table A: Prioritisation strata according to HIV incidence in the general population and behavioural risk.

| Intervention | Low | Moderate | High | Very High |
|---|---|---|---|---|
| Condoms and lube for those with non-regular partners(s) with unknown STI status and not on PrEP | 50% | 70% | 95% | 95% |
| STI screening and treatment | 10% | 10% | 80% | 80% |
| Access to PEP | - | - | 50% | 90% |
| PrEP use | - | 5% | 50% | 50% |
| Economic empowerment | - | - | 20% | 20% |

Table B: Commitments to be met for each intervention in terms of proportion of the prioritisation strata reached, where "-" represents no commitment.

### Household survey data

#### Included surveys

| | Type | Year | Transactional sex question | Sample size | | | |
|---|---|---|---|---|---|---|---|
| | | | | 15-19 | 20-24 | 25-29 | Total |
| Botswana | | | | | | | |
| | BAIS | 2013 | ✓ | 557 | 588 | 649 | 1794 |
| Total | | | | 557 | 588 | 649 | 1794 |
| Cameroon | | | | | | | |
| | DHS | 2004 | ✗ | 2675 | 2207 | 1732 | 6614 |
| | DHS | 2011 | ✗ | 3588 | 3115 | 2655 | 9358 |
| | PHIA | 2017 | ✗ | 2620 | 2339 | 2259 | 7218 |
| | DHS | 2018 | ✓ | 3349 | 2463 | 2345 | 8157 |
| Total | | | | 12232 | 10124 | 8991 | 31347 |
| Kenya | | | | | | | |
| | DHS | 2003 | ✗ | 1819 | 1709 | 1391 | 4919 |
| | DHS | 2008 | ✗ | 1767 | 1743 | 1419 | 4929 |
| | DHS | 2014 | ✗ | 2861 | 2534 | 2858 | 8253 |
| Total | | | | 6447 | 5986 | 5668 | 18101 |
| Lesotho | | | | | | | |
| | DHS | 2004 | ✗ | 1761 | 1455 | 1026 | 4242 |
| | DHS | 2009 | ✗ | 1833 | 1543 | 1194 | 4570 |
| | DHS | 2014 | ✗ | 1537 | 1292 | 1067 | 3896 |
| | PHIA | 2017 | ✓ | 1156 | 1202 | 1054 | 3412 |
| Total | | | | 6287 | 5492 | 4341 | 16120 |
| Mozambique | | | | | | | |
| | AIS | 2009 | ✗ | 1031 | 1106 | 987 | 3124 |
| | DHS | 2011 | ✗ | 2932 | 2299 | 2206 | 7437 |
| | AIS | 2015 | ✗ | 1552 | 1389 | 1080 | 4021 |
| Total | | | | 5515 | 4794 | 4273 | 14582 |
| Malawi | | | | | | | |
| | DHS | 2000 | ✗ | 2914 | 2998 | 2358 | 8270 |
| | DHS | 2004 | ✗ | 2407 | 2823 | 2135 | 7365 |
| | DHS | 2010 | ✗ | 5031 | 4387 | 4309 | 13727 |
| | DHS | 2015 | ✓ | 5273 | 5094 | 3976 | 14343 |
| | PHIA | 2016 | ✓ | 1646 | 1934 | 1511 | 5091 |
| Total | | | | 17271 | 17236 | 14289 | 48796 |
| Namibia | | | | | | | |
| | DHS | 2000 | ✗ | 1427 | 1313 | 1098 | 3838 |
| | DHS | 2006 | ✗ | 2203 | 1869 | 1544 | 5616 |
| | DHS | 2013 | ✗ | 1852 | 1709 | 1481 | 5042 |
| | PHIA | 2017 | ✓ | 1491 | 1525 | 1370 | 4386 |
| Total | | | | 6973 | 6416 | 5493 | 18882 |
| Eswatini | | | | | | | |
| | DHS | 2006 | ✗ | 1265 | 1027 | 731 | 3023 |
| | PHIA | 2017 | ✗ | 1031 | 895 | 811 | 2737 |

| | | | | | | | |
|---|---|---|---|---|---|---|---|
| Total | | | | 2296 | 1922 | 1542 | 5760 |
| Tanzania | | | | | | | |
| | AIS | 2003 | ✗ | 1466 | 1377 | 1270 | 4113 |
| | AIS | 2007 | ✗ | 2137 | 1676 | 1509 | 5322 |
| | DHS | 2010 | ✗ | 2221 | 1860 | 1613 | 5694 |
| | AIS | 2012 | ✗ | 2474 | 1923 | 1815 | 6212 |
| | PHIA | 2016 | ✓ | 2999 | 2845 | 2521 | 8365 |
| Total | | | | 11297 | 9681 | 8728 | 29706 |
| Uganda | | | | | | | |
| | DHS | 2000 | ✗ | 1687 | 1541 | 1326 | 4554 |
| | DHS | 2006 | ✗ | 1948 | 1660 | 1404 | 5012 |
| | AIS | 2011 | ✗ | 2451 | 2164 | 1921 | 6536 |
| | DHS | 2011 | ✗ | 2025 | 1664 | 1614 | 5303 |
| | DHS | 2016 | ✓ | 4276 | 3782 | 3014 | 11072 |
| | PHIA | 2016 | ✗ | 3289 | 3059 | 2574 | 8922 |
| Total | | | | 15676 | 13870 | 11853 | 41399 |
| South Africa | | | | | | | |
| | DHS | 2016 | ✓ | 1505 | 1408 | 1397 | 4310 |
| Total | | | | 1505 | 1408 | 1397 | 4310 |
| Zambia | | | | | | | |
| | DHS | 2007 | ✗ | 1598 | 1405 | 1373 | 4376 |
| | DHS | 2013 | ✗ | 3685 | 3036 | 2789 | 9510 |
| | PHIA | 2016 | ✓ | 2120 | 2045 | 1619 | 5784 |
| | DHS | 2018 | ✓ | 3112 | 2687 | 2166 | 7965 |
| Total | | | | 10515 | 9173 | 7947 | 27635 |
| Zimbabwe | | | | | | | |
| | DHS | 1999 | ✗ | 1467 | 1230 | 1011 | 3708 |
| | DHS | 2005 | ✗ | 2128 | 1943 | 1438 | 5509 |
| | DHS | 2010 | ✗ | 1963 | 1796 | 1679 | 5438 |
| | DHS | 2015 | ✓ | 2154 | 1777 | 1646 | 5577 |
| | PHIA | 2016 | ✓ | 2114 | 1817 | 1573 | 5504 |
| Total | | | | 9826 | 8563 | 7347 | 25736 |
| Total | | | | 106397 | 95253 | 82518 | 284168 |

Table C: All of the surveys that we used in our analysis and their sample sizes, disaggregated by respondent age.

| Survey | Exclusion reason |
|---|---|
| MOZ2003DHS | No GPS coordinates available to place survey clusters within districts. |
| TZA2015DHS | Insufficient sexual behaviour questions. |
| UGA2004AIS | Unable to download region boundaries. |
| ZMB2002DHS | No GPS coordinates available to place survey clusters within districts. |

Table D: All of that surveys that were excluded from our analysis.

#### Spatial analysis levels

| Country | Number of areas | Analysis level |
|---|---|---|
| Botswana | 27 | 3 |
| Cameroon | 58 | 2 |
| Kenya | 47 | 2 |
| Lesotho | 10 | 1 |
| Mozambique | 161 | 3 |
| Malawi | 33 | 5 |
| Namibia | 38 | 2 |
| Eswatini | 4 | 1 |
| Tanzania | 195 | 4 |
| Uganda | 136 | 3 |
| South Africa | 52 | 2 |
| Zambia | 116 | 2 |
| Zimbabwe | 63 | 2 |

Table E: The numer of areas and analysis levels for each country that were used in our analysis.

#### Survey questions and risk group allocation

| Variable(s) | Description |
|---|---|
| v501 | Current marital status of the respondent. |
| v529 | Computed time since last sexual intercourse. |
| v531 | Age at first sexual intercourse–imputed. |
| v766b | Number of sexual partners during the last 12 months (including husband). |
| v767[a, b, c] | Relationship with last three sexual partners. Options are: spouse, boyfriend not living with respondent, other friend, casual acquaintance, relative, commercial sex worker, live-in partner, other. |
| v791a | Had sex in return for gifts, cash or anything else in the past 12 months. Asked only to women 15-24 who are not in a union. |

Table F: AIS, BAIS and DHS survey questions.

| Variable(s) | Description |
|---|---|
| part12monum | Number of sexual partners during the last 12 months (including husband). |
| part12modkr | Reason for leaving part12monum blank. |
| partlivew[1, 2, 3] | Does the person you had sex with live in this household? |
| partrelation[1, 2, 3] | Relationship with last three sexual partners. Options are: husband, live-in partner, partner (not living with), ex-spouse/partner, friend/acquaintance, sex worker, sex worker client, stranger, other, don't know, refused. |
| sellsx12mo | Had sex for money and/or gifts in the last 12 months. |
| buysx12mo | Paid money or given gifts for sex in the last 12 months. |

Table G: PHIA survey questions.

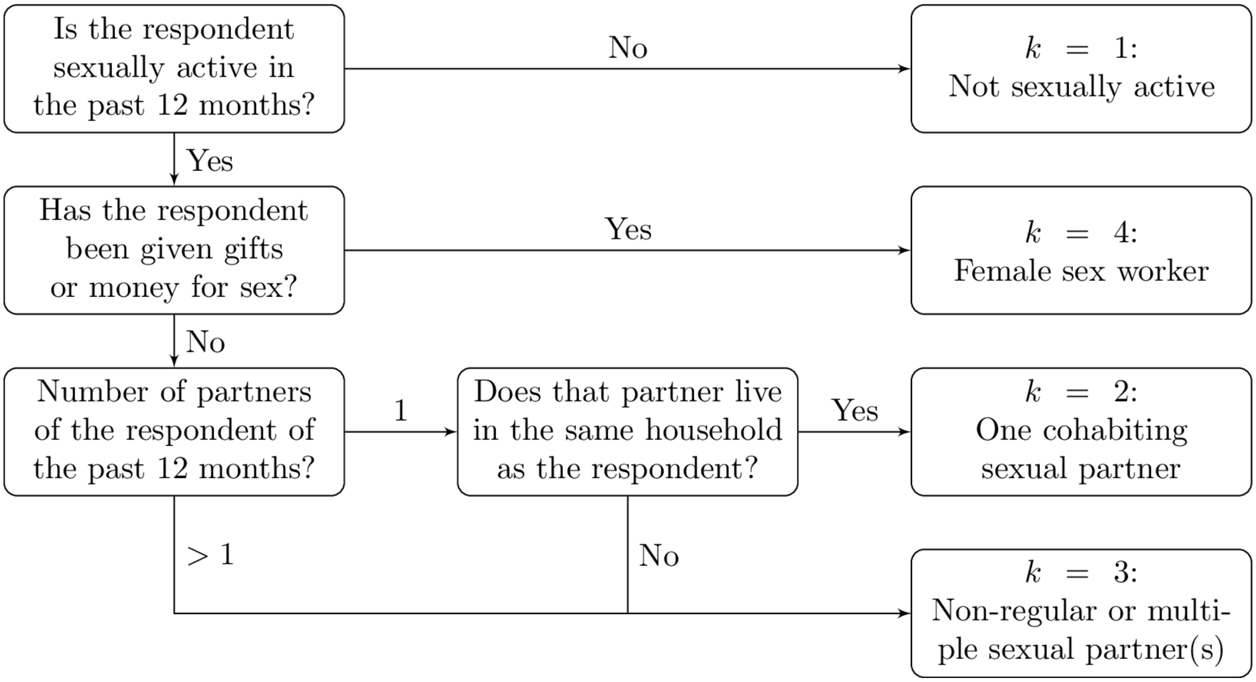

Figure A: Flowchart describing allocation of respondents to risk groups.

#### Miscellaneous figures

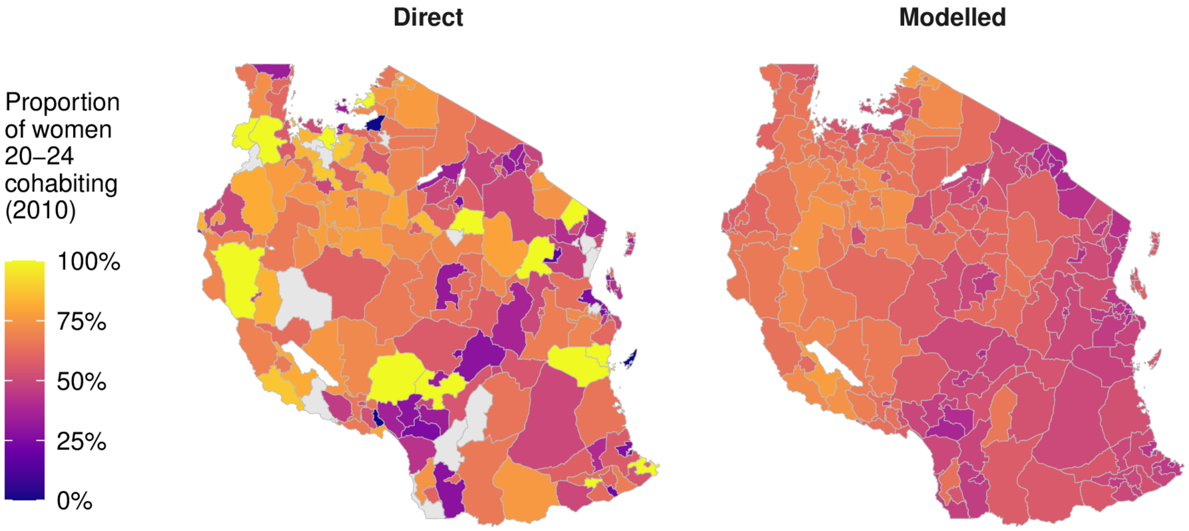

Figure B: Illustration of our model results for AGYW 20-24 in Tanzania in 2010 in the cohabiting risk group. Compared to the direct survey results, our spatio-temporally smoothed estimates more plausibly represent district-level heterogeneity, as well as imputing any districts with missing data.

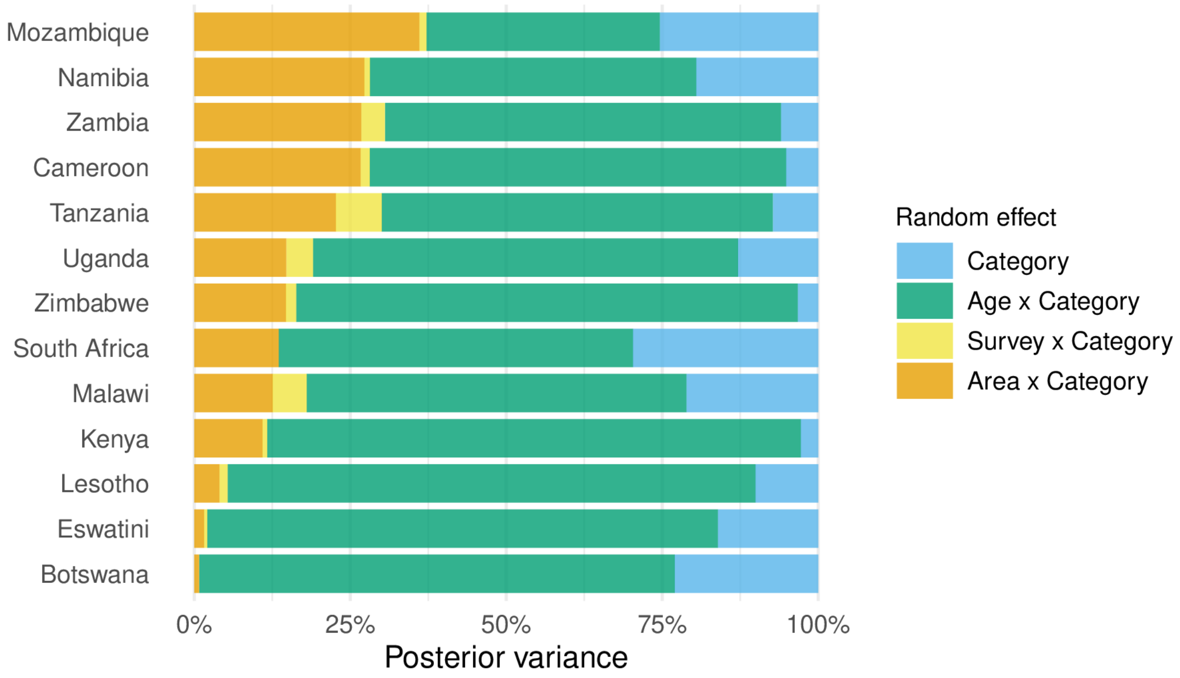

Figure C: Proportion of variance explained by each random effect (Sobol' indices) when the multinomial regression model is fit to each country individually. In this setting, country-category random effects are not included in the model and year-category random effects are replaced by survey-category random effects (for countries with surveys in multiple years). Countries are ordered by the proportion of their variance which is explained by the area-category random effects.

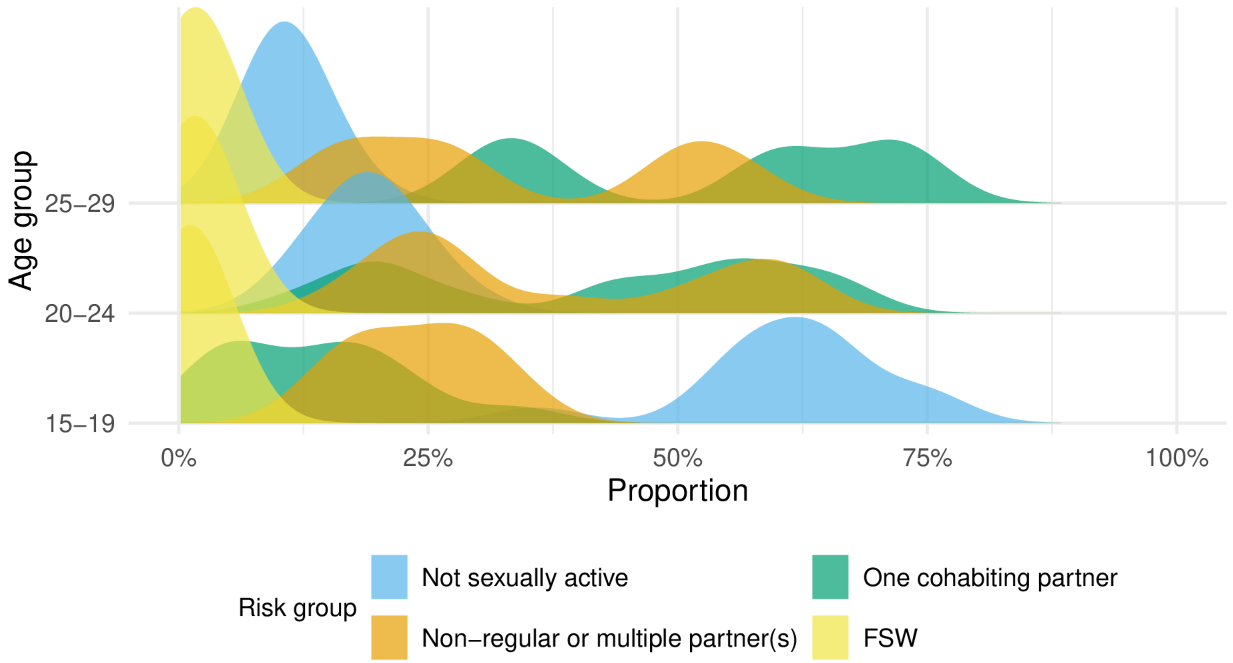

Figure D: The posterior density of national-level risk group proportions by age, illustrating the bi-modality of the cohabiting partner and non-regular and multiple partner(s) risk groups.

### Country-specific figures

#### Comparison of direct and modelled risk group estimates

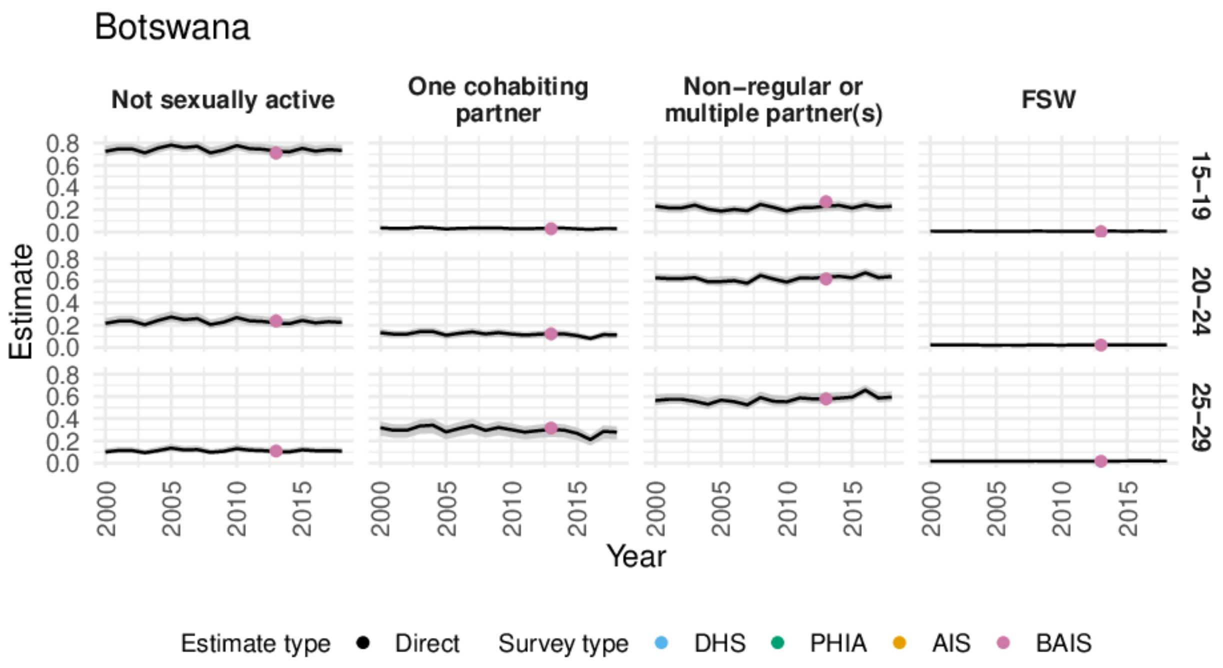

Figure E: Comparison of modelled and direct national-level estimates in 1999-2018 in Botswana. Estimates are described as "partially direct" when there are no surveys containing a transactional sex question in a country-age-group and we instead used modelled logistic regression estimates to differentiate the direct estimates.

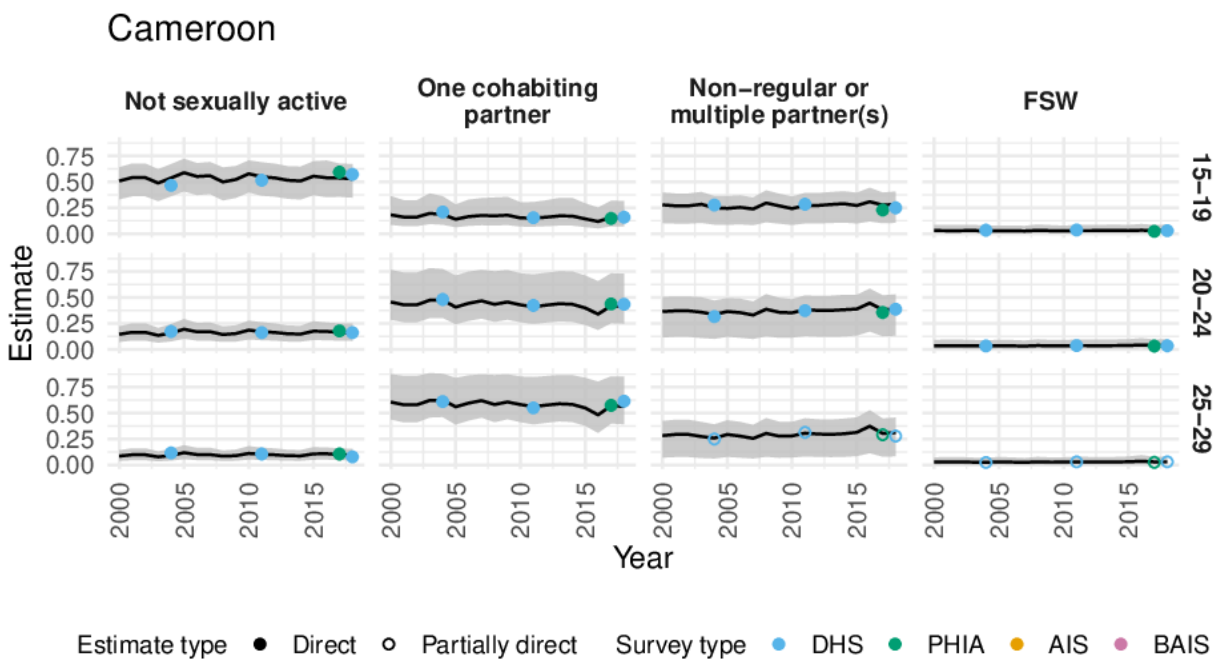

Figure F: Comparison of modelled and direct national-level estimates in 1999-2018 in Cameroon. Estimates are described as "partially direct" when there are no surveys containing a transactional sex question in a country-age-group and we instead used modelled logistic regression estimates to differentiate the direct estimates.

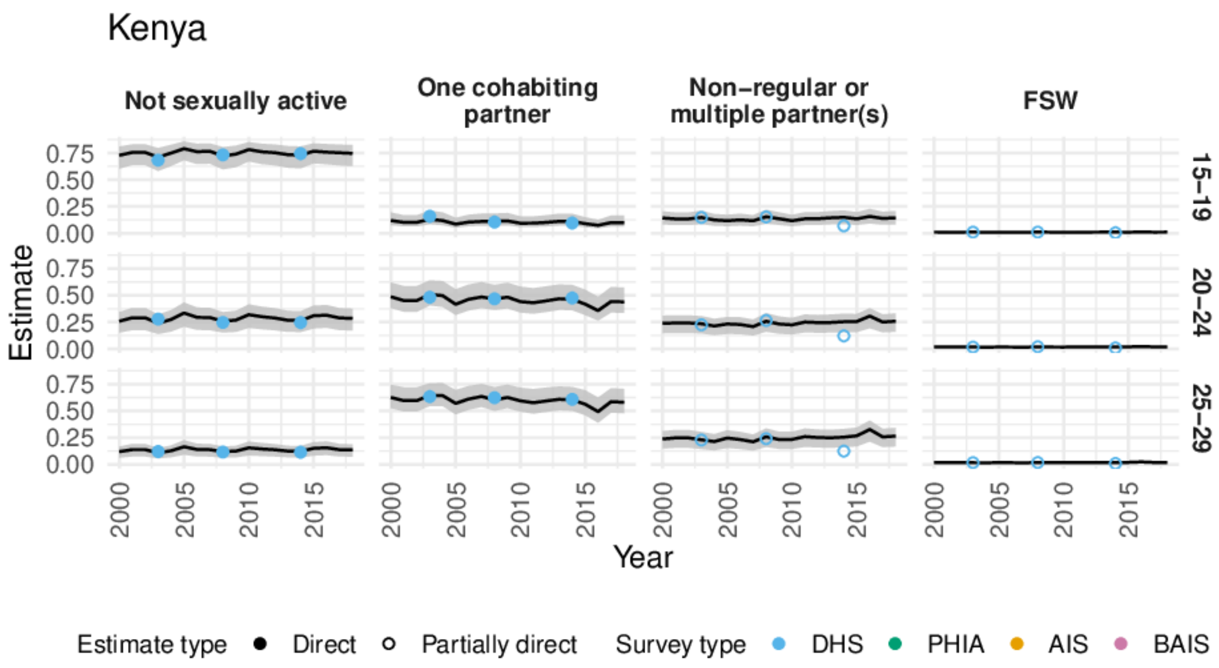

Figure G: Comparison of modelled and direct national-level estimates in 1999-2018 in Kenya. Estimates are described as "partially direct" when there are no surveys containing a transactional sex question in a country-age-group and we instead used modelled logistic regression estimates to differentiate the direct estimates.

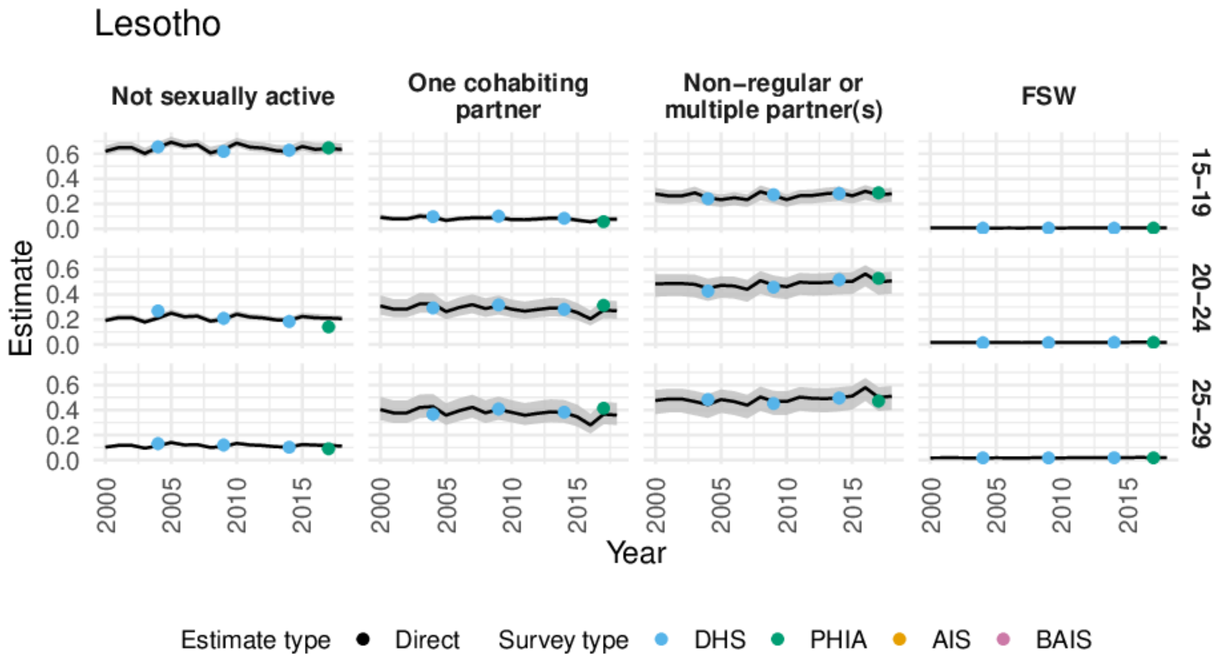

Figure H: Comparison of modelled and direct national-level estimates in 1999-2018 in Lesotho. Estimates are described as "partially direct" when there are no surveys containing a transactional sex question in a country-age-group and we instead used modelled logistic regression estimates to differentiate the direct estimates.

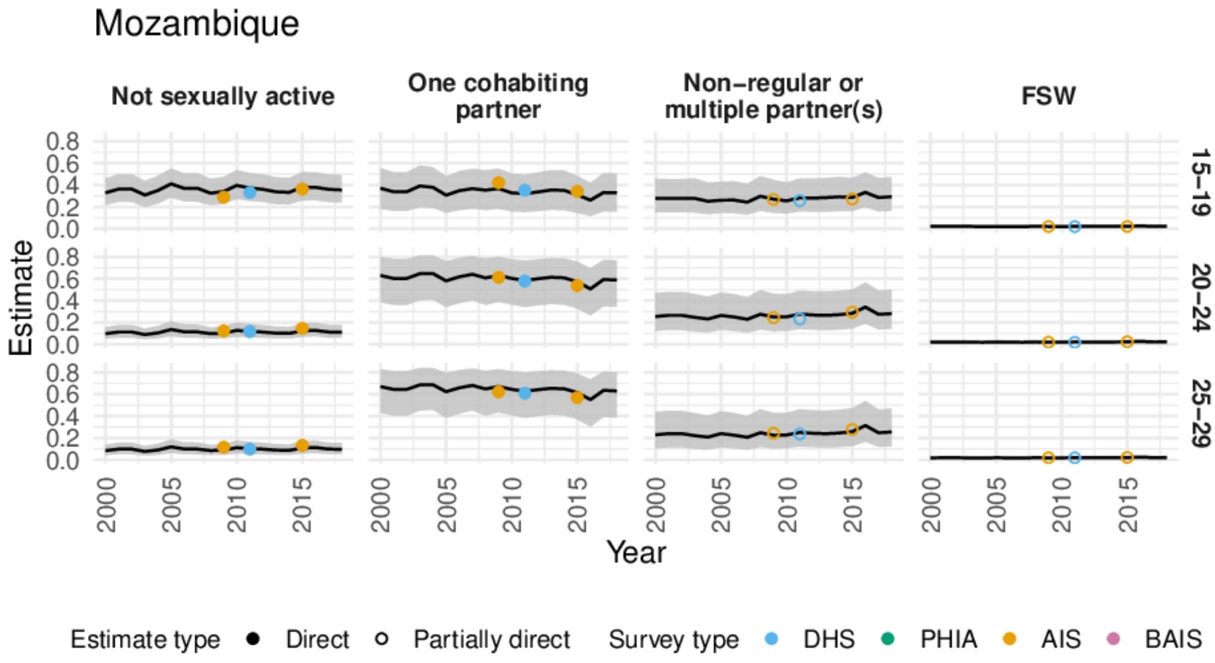

Figure I: Comparison of modelled and direct national-level estimates in 1999-2018 in Mozambique. Estimates are described as "partially direct" when there are no surveys containing a transactional sex question in a country-age-group and we instead used modelled logistic regression estimates to differentiate the direct estimates.

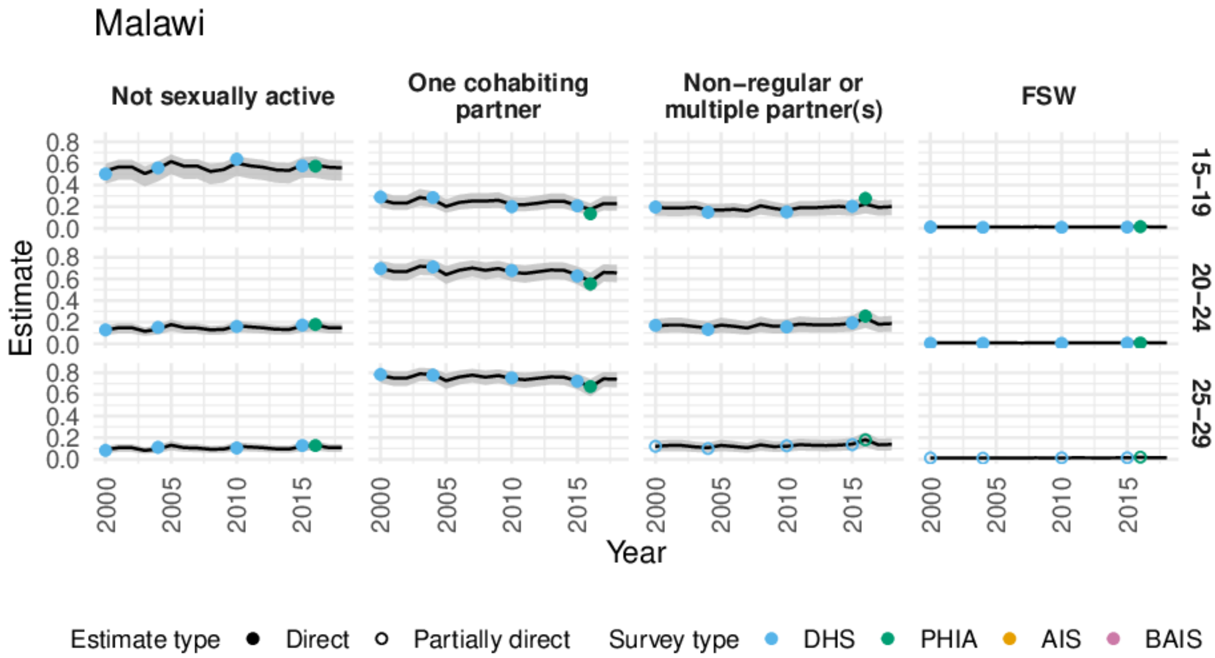

Figure J: Comparison of modelled and direct national-level estimates in 1999-2018 in Malawi. Estimates are described as "partially direct" when there are no surveys containing a transactional sex question in a country-age-group and we instead used modelled logistic regression estimates to differentiate the direct estimates.

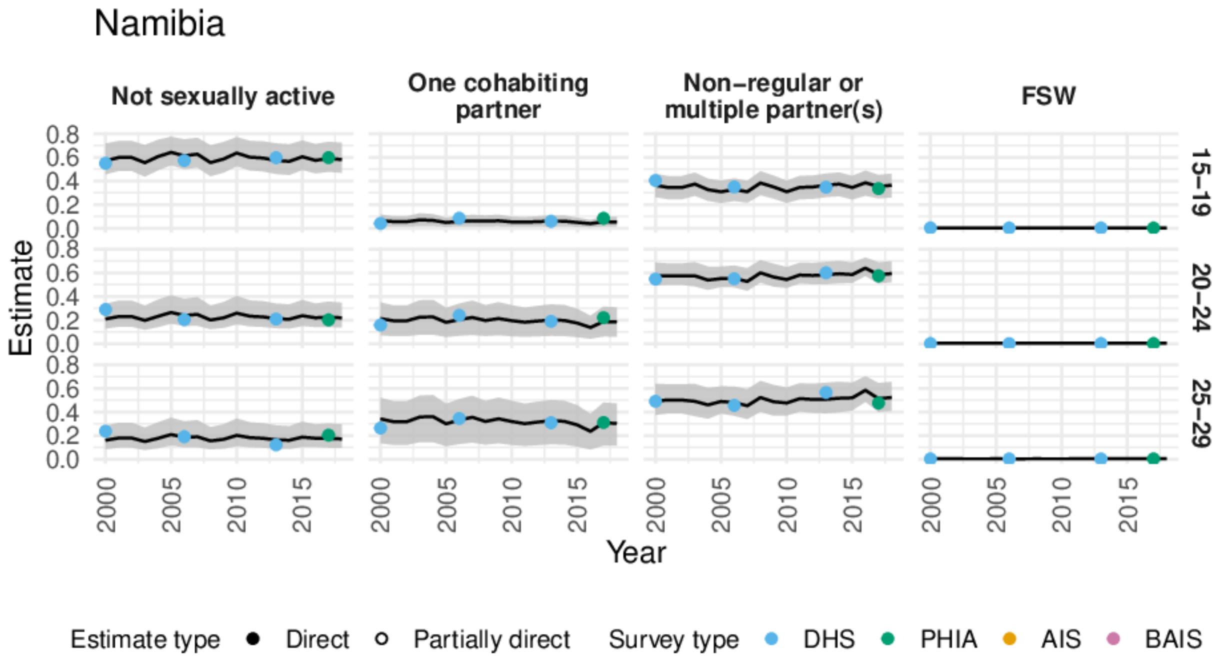

Figure K: Comparison of modelled and direct national-level estimates in 1999-2018 in Namibia. Estimates are described as "partially direct" when there are no surveys containing a transactional sex question in a country-age-group and we instead used modelled logistic regression estimates to differentiate the direct estimates.

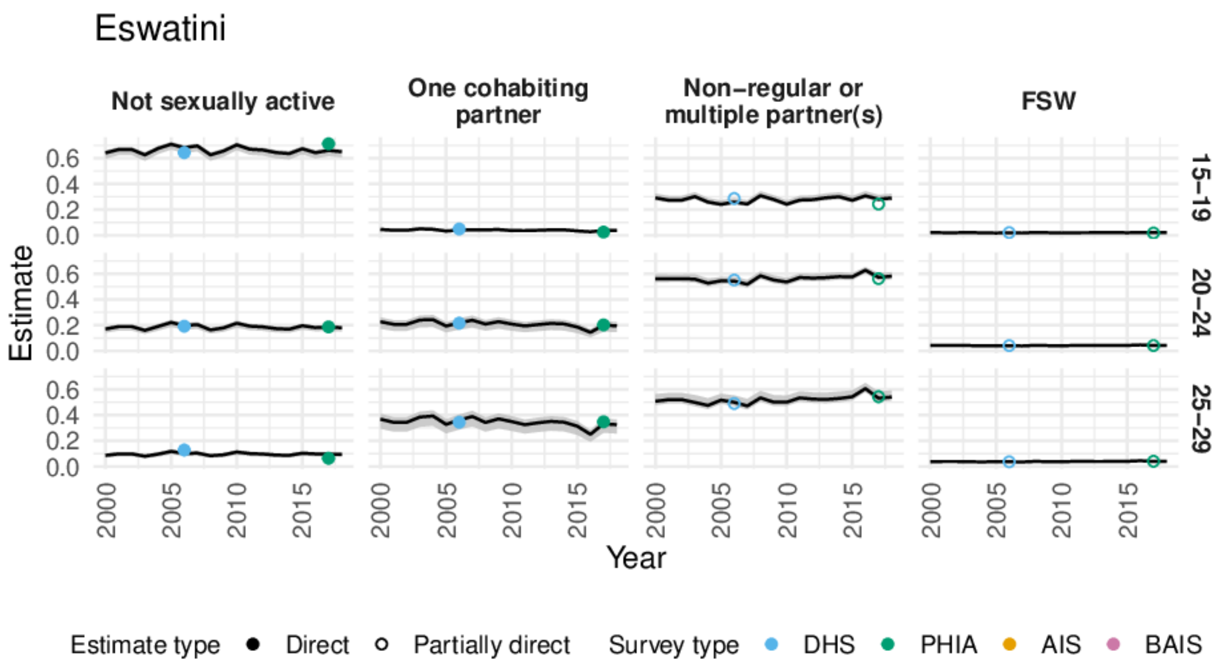

Figure L: Comparison of modelled and direct national-level estimates in 1999-2018 in Eswatini. Estimates are described as "partially direct" when there are no surveys containing a transactional sex question in a country-age-group and we instead used modelled logistic regression estimates to differentiate the direct estimates.

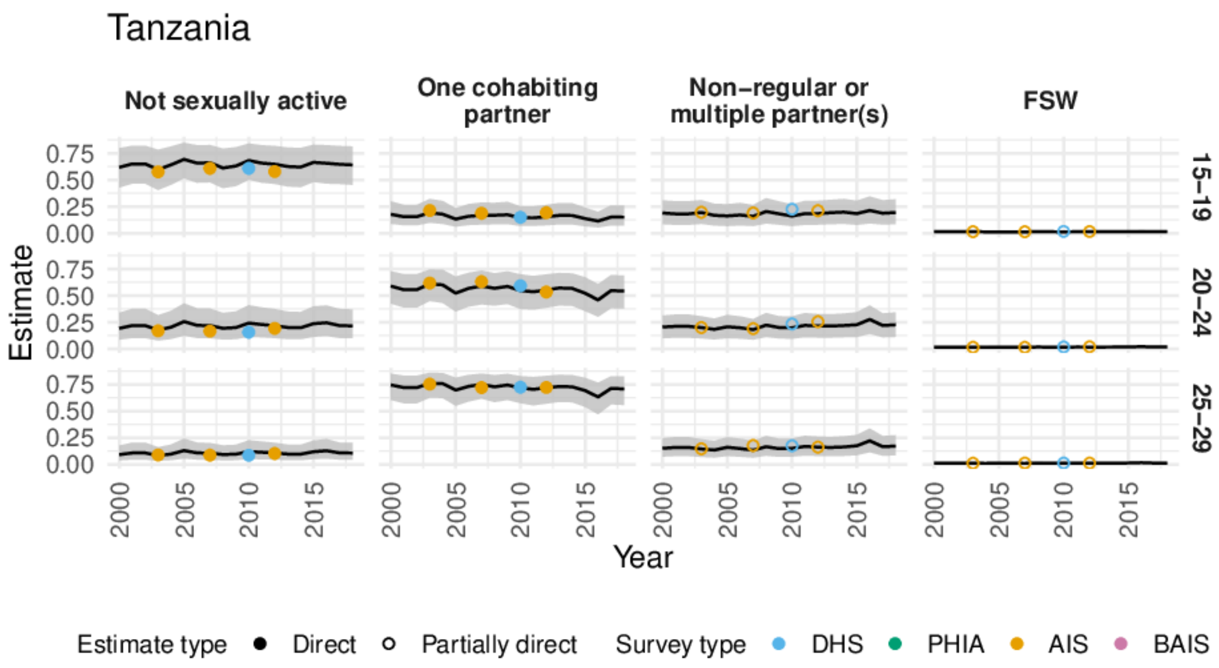

Figure M: Comparison of modelled and direct national-level estimates in 1999-2018 in Tanzania. Estimates are described as "partially direct" when there are no surveys containing a transactional sex question in a country-age-group and we instead used modelled logistic regression estimates to differentiate the direct estimates.

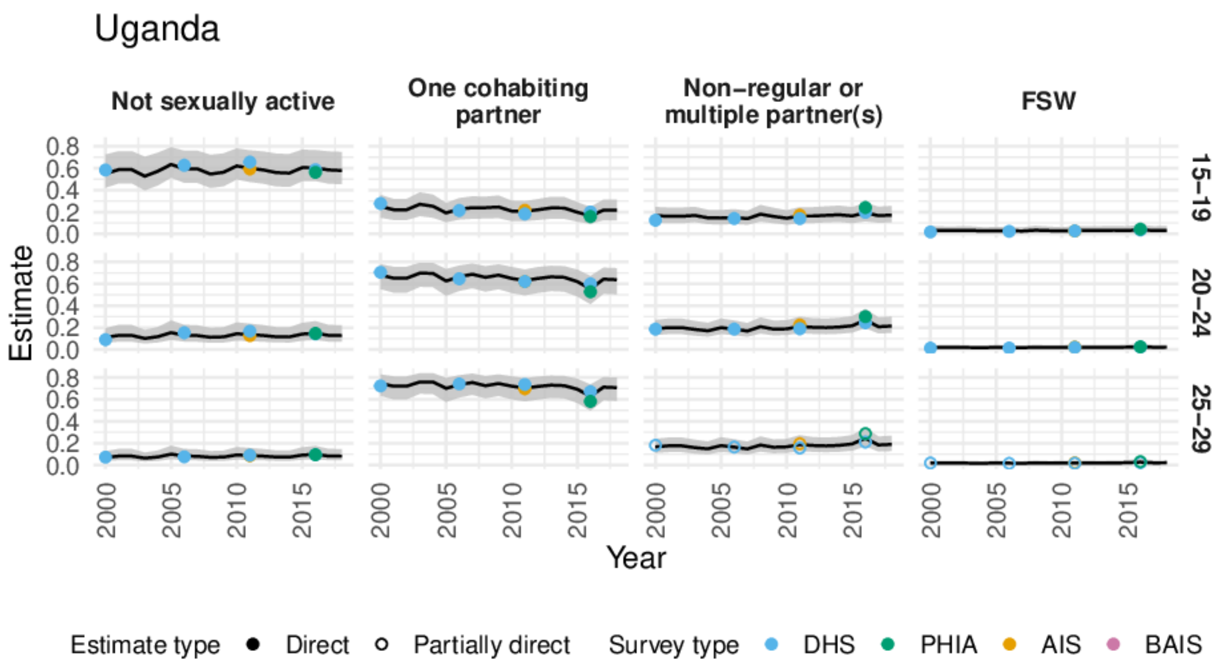

Figure N: Comparison of modelled and direct national-level estimates in 1999-2018 in Uganda. Estimates are described as "partially direct" when there are no surveys containing a transactional sex question in a country-age-group and we instead used modelled logistic regression estimates to differentiate the direct estimates.

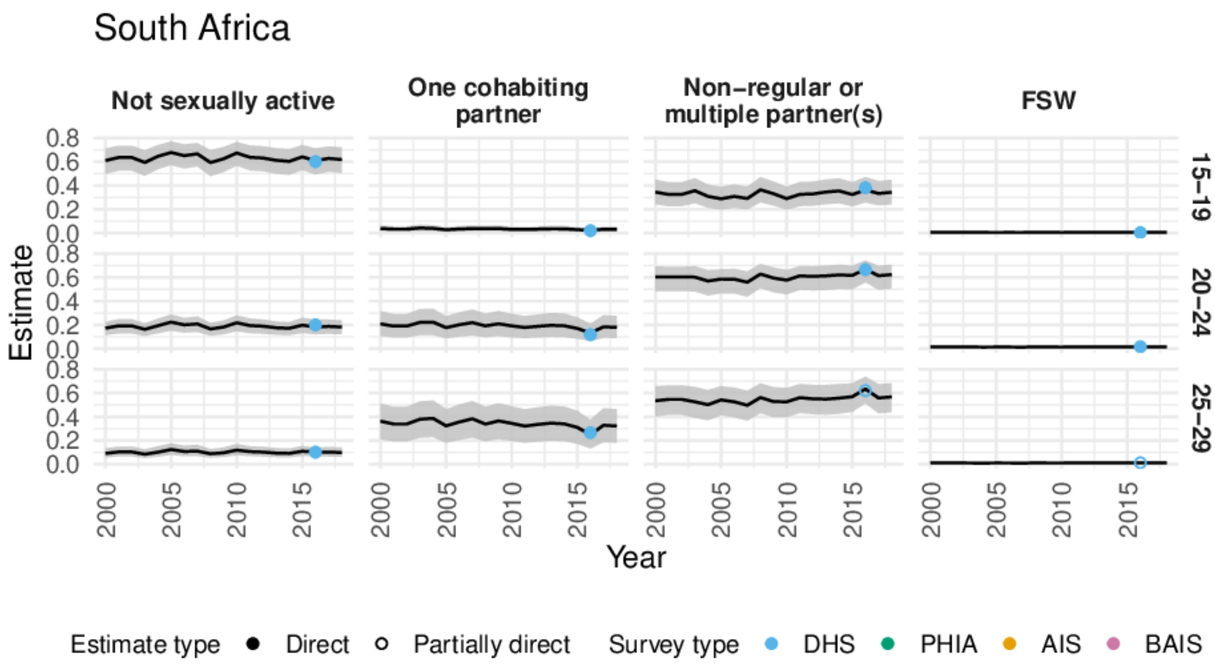

Figure O: Comparison of modelled and direct national-level estimates in 1999-2018 in South Africa. Estimates are described as "partially direct" when there are no surveys containing a transactional sex question in a country-age-group and we instead used modelled logistic regression estimates to differentiate the direct estimates.

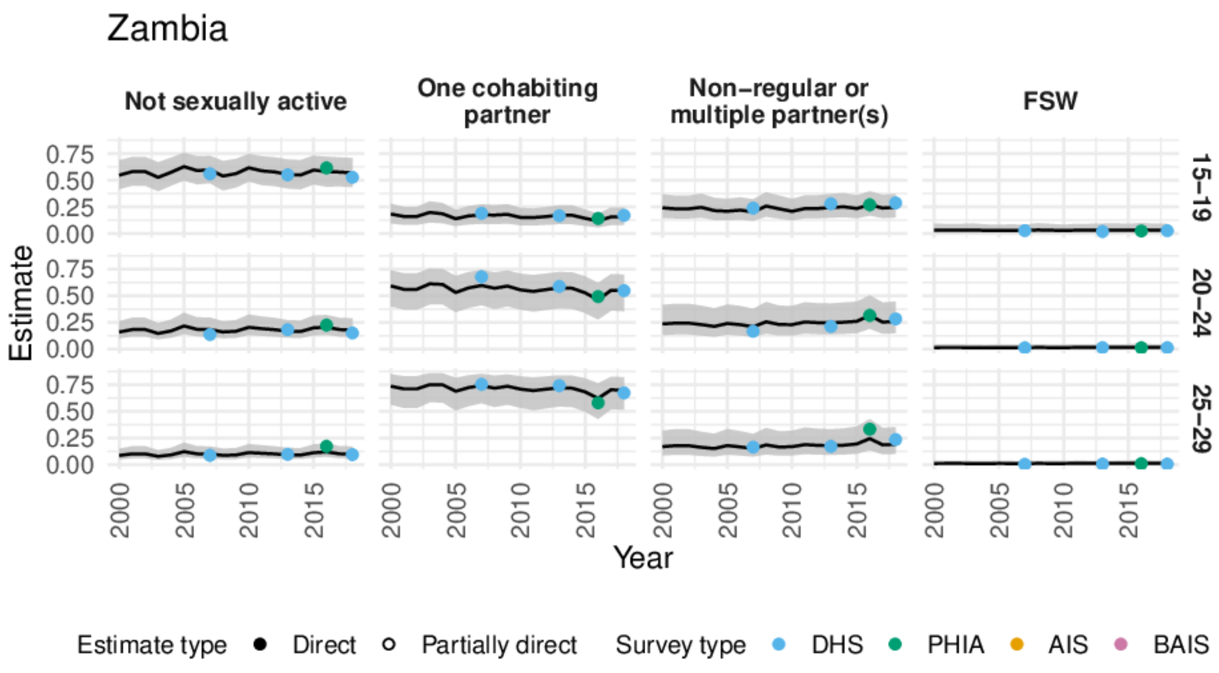

Figure P: Comparison of modelled and direct national-level estimates in 1999-2018 in Zambia. Estimates are described as "partially direct" when there are no surveys containing a transactional sex question in a country-age-group and we instead used modelled logistic regression estimates to differentiate the direct estimates.

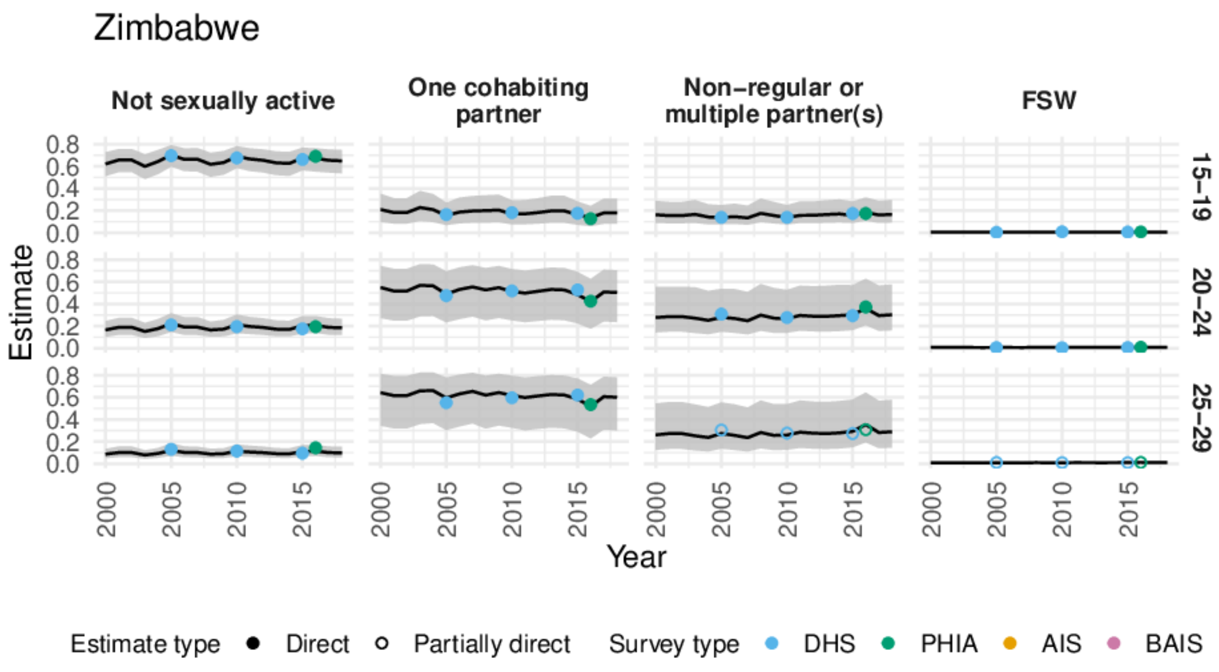

Figure Q: Comparison of modelled and direct national-level estimates in 1999-2018 in Zimbabwe. Estimates are described as "partially direct" when there are no surveys containing a transactional sex question in a country-age-group and we instead used modelled logistic regression estimates to differentiate the direct estimates.

**HIV prevalence**

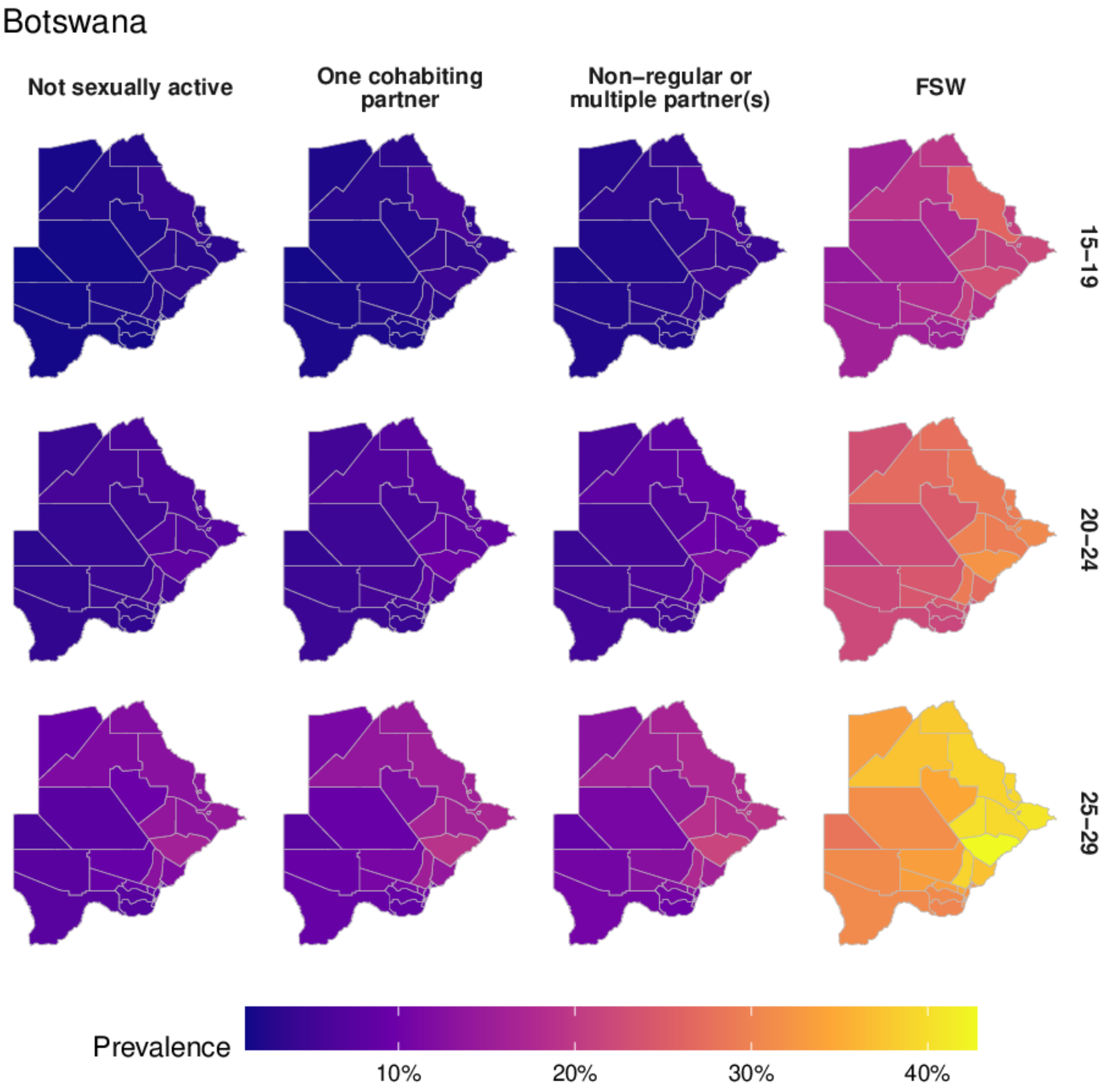

Figure R: District-level HIV prevalence for each of the risk groups in 2018 in Botswana.

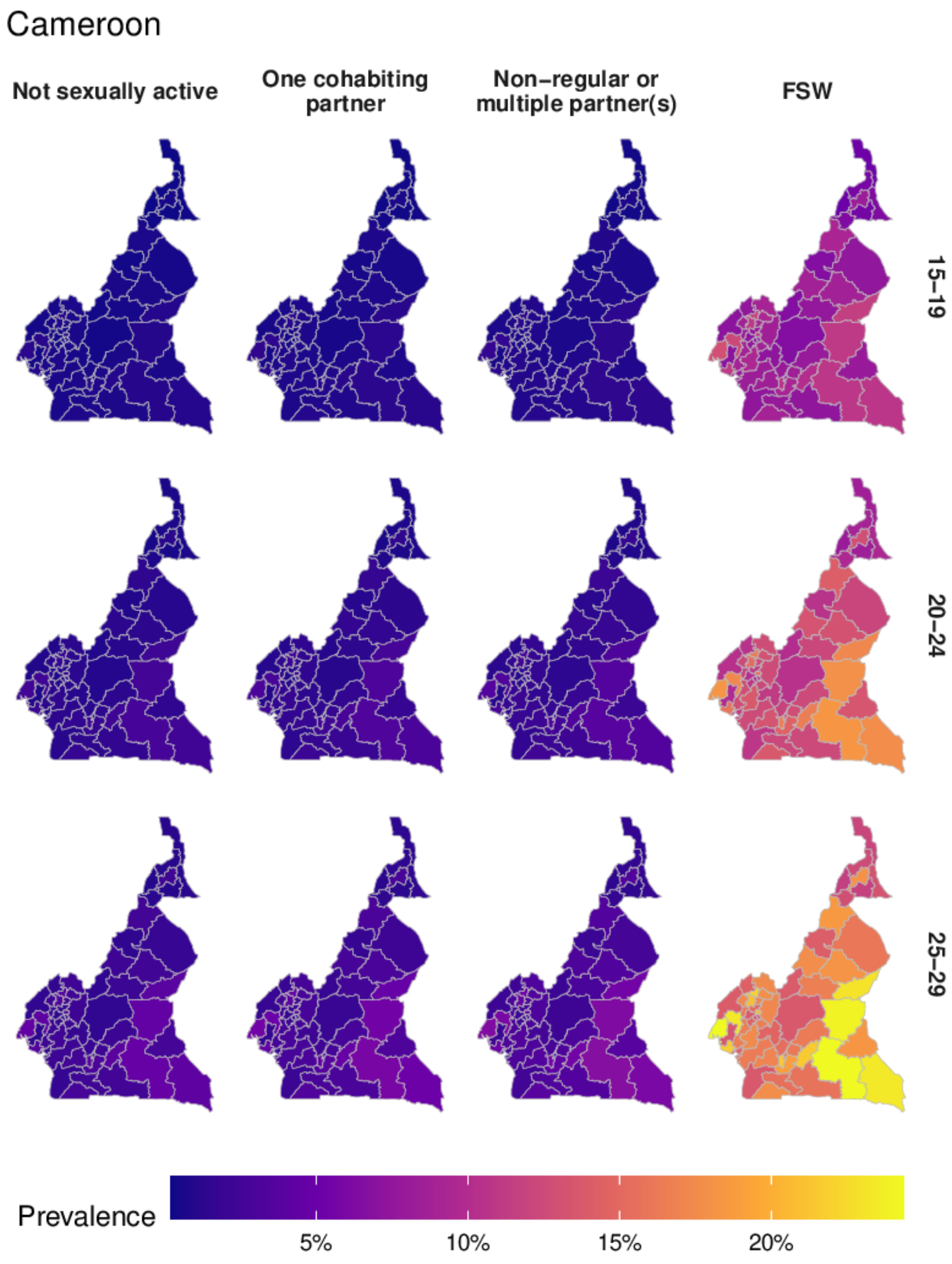

Figure S: District-level HIV prevalence for each of the risk groups in 2018 in Cameroon.

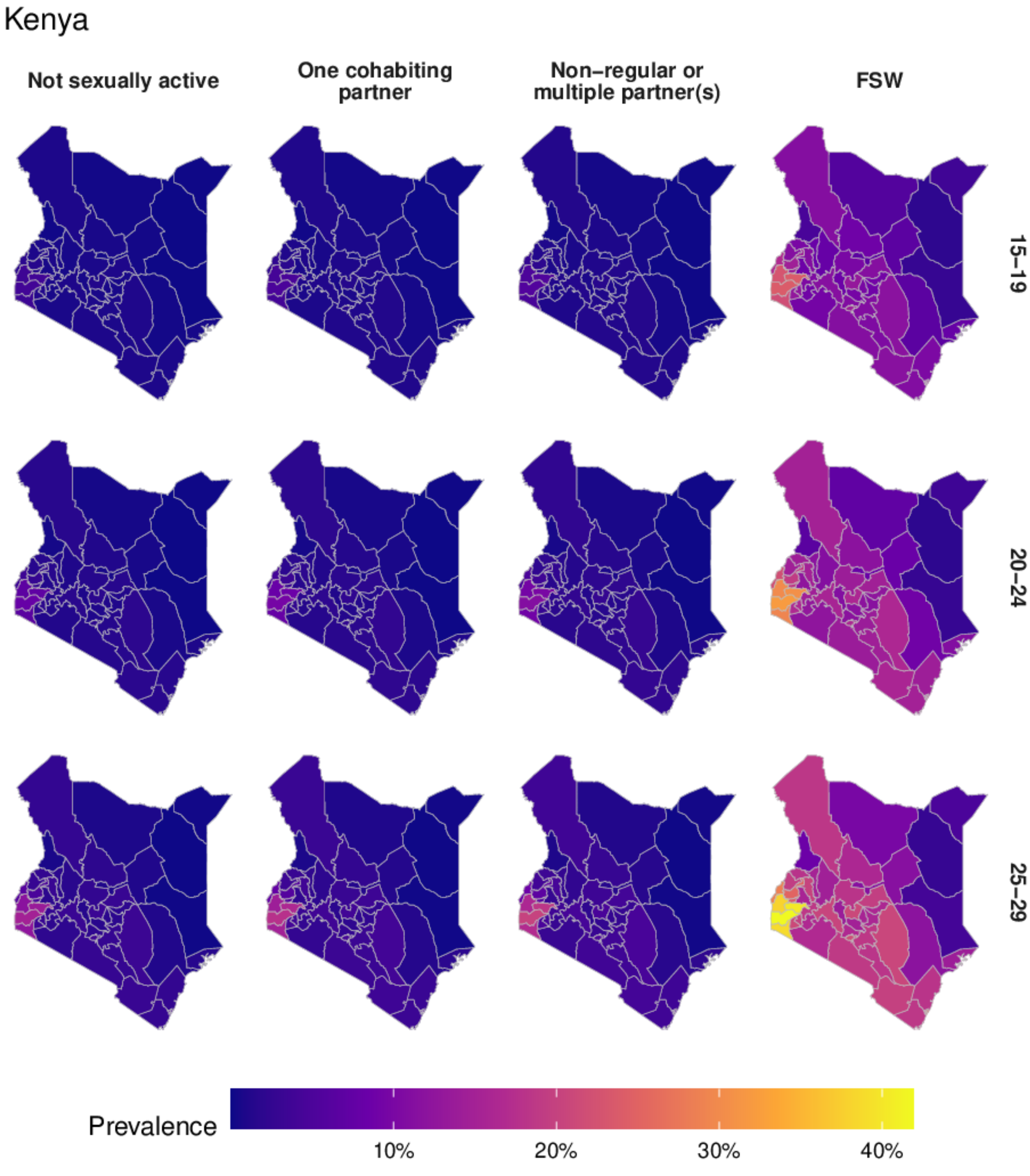

Figure T: District-level HIV prevalence for each of the risk groups in 2018 in Kenya.

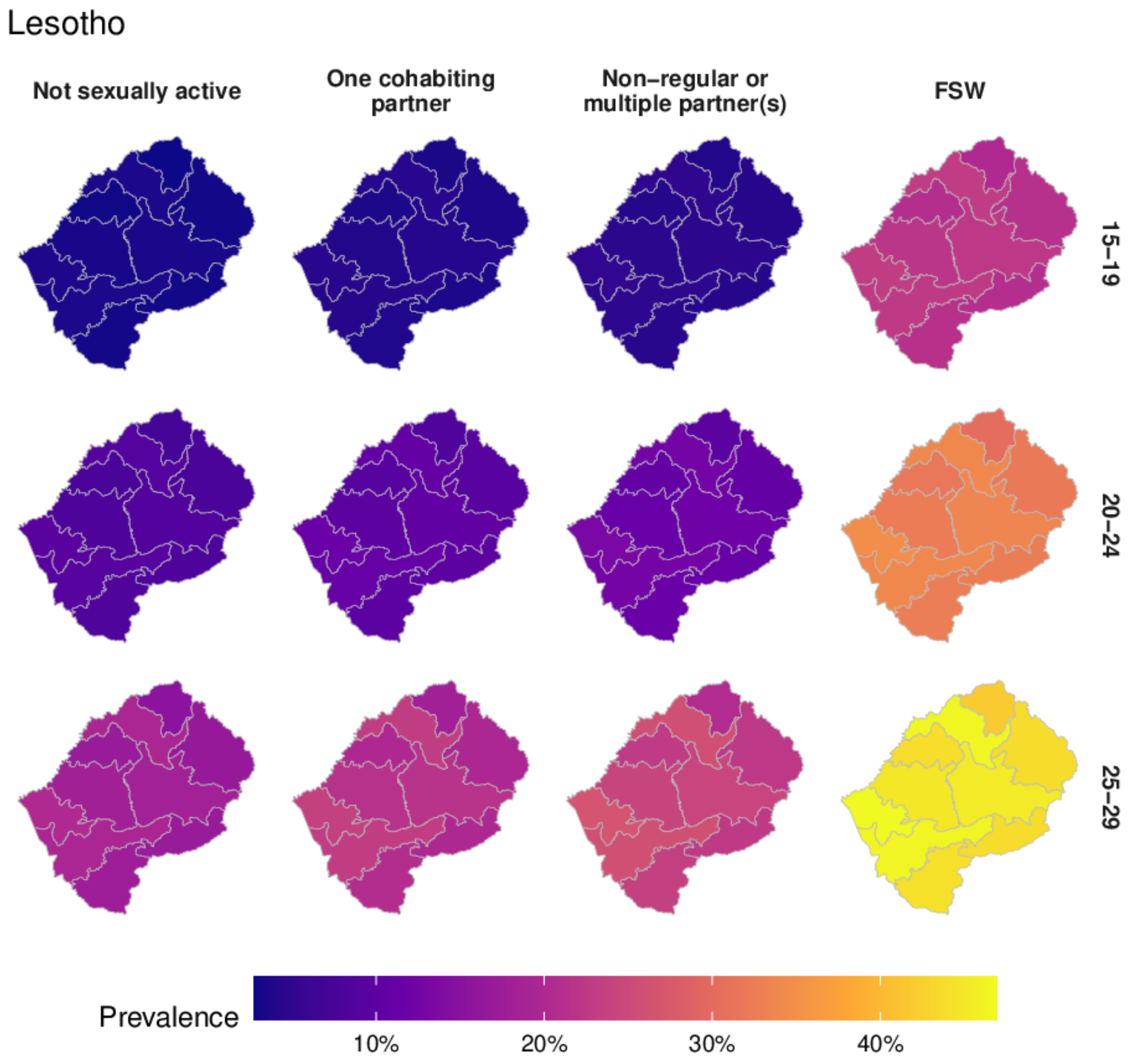

Figure U: District-level HIV prevalence for each of the risk groups in 2018 in Lesotho.

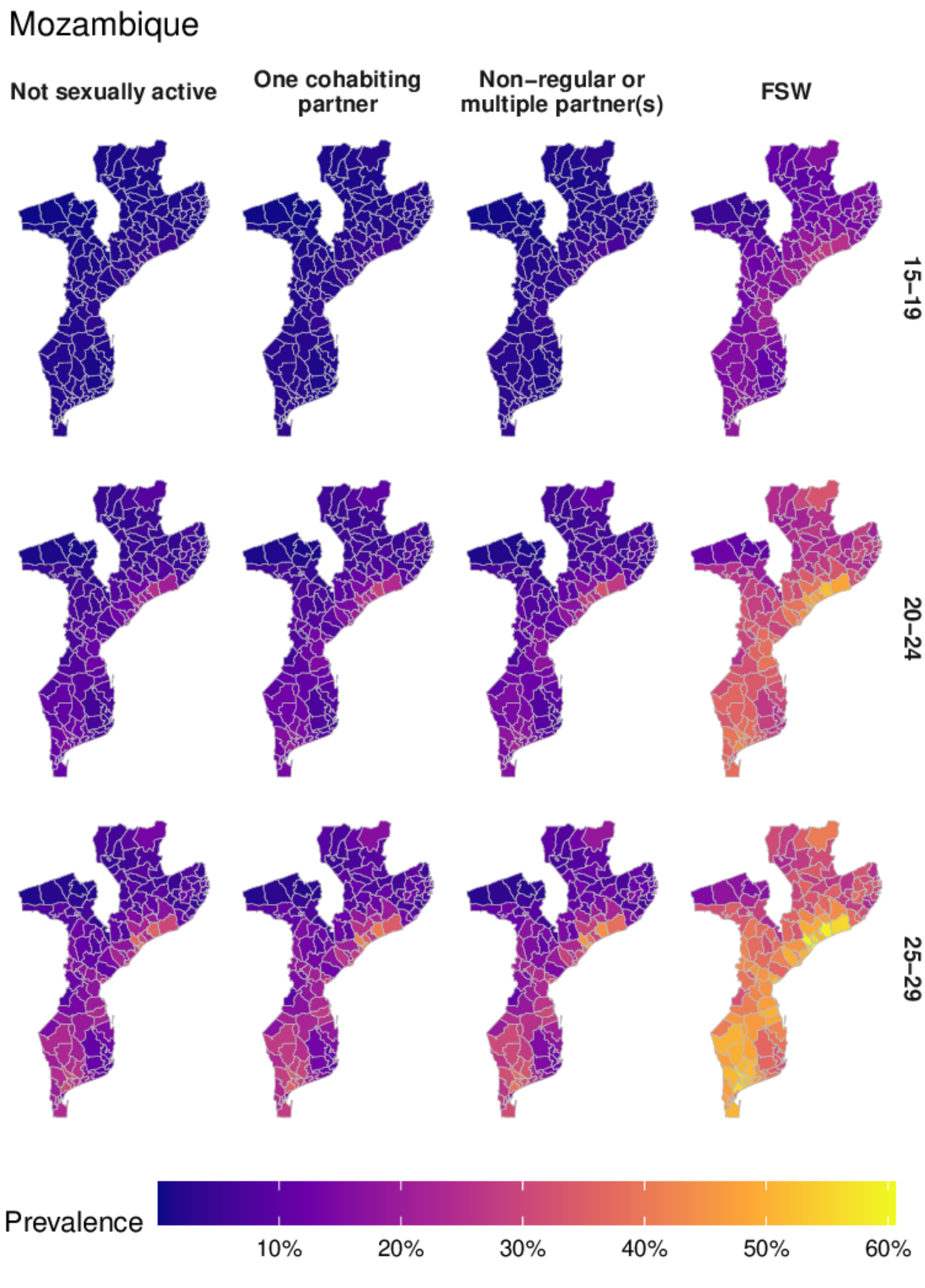

Figure V: District-level HIV prevalence for each of the risk groups in 2018 in Mozambique.

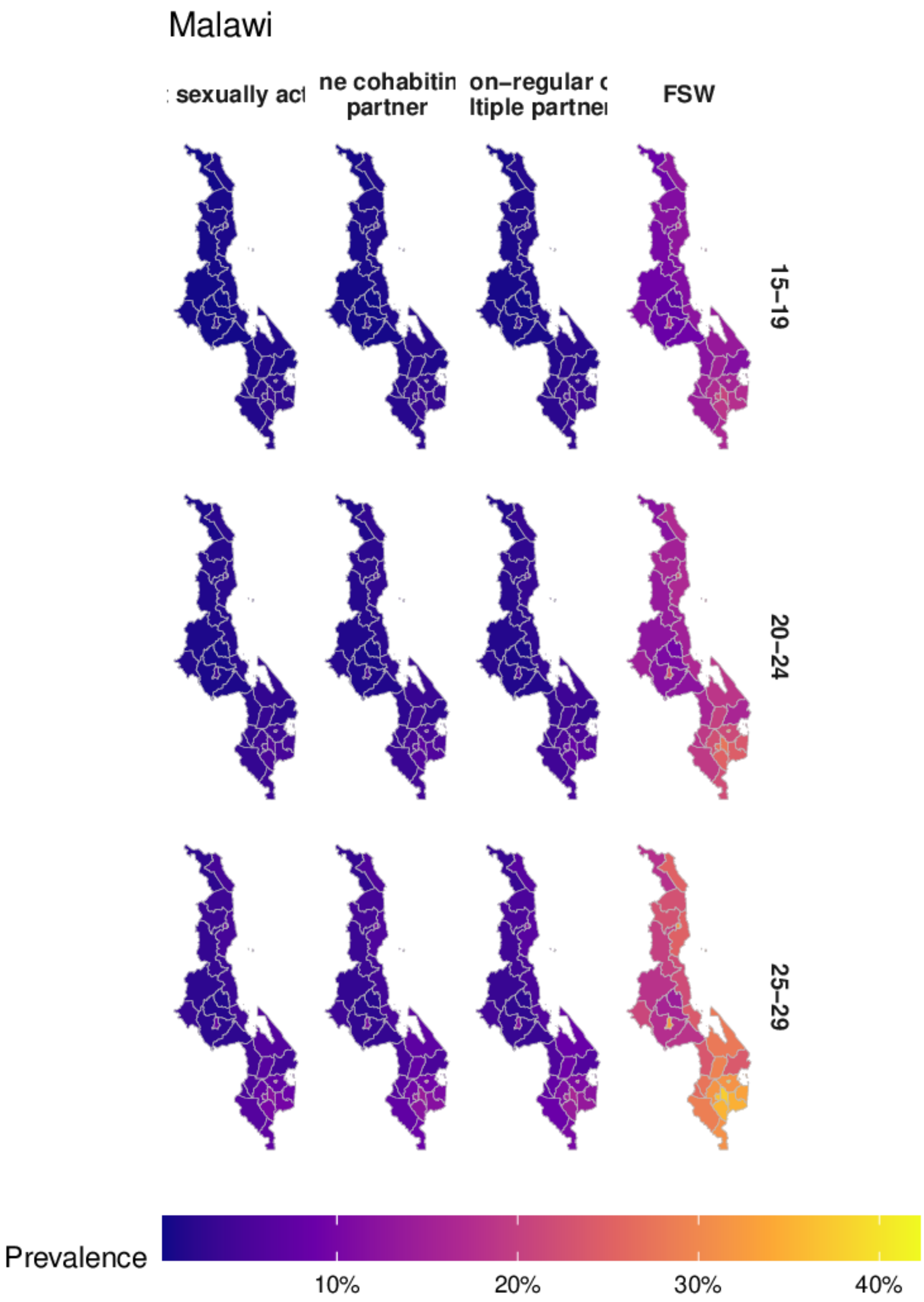

Figure W: District-level HIV prevalence for each of the risk groups in 2018 in Malawi.

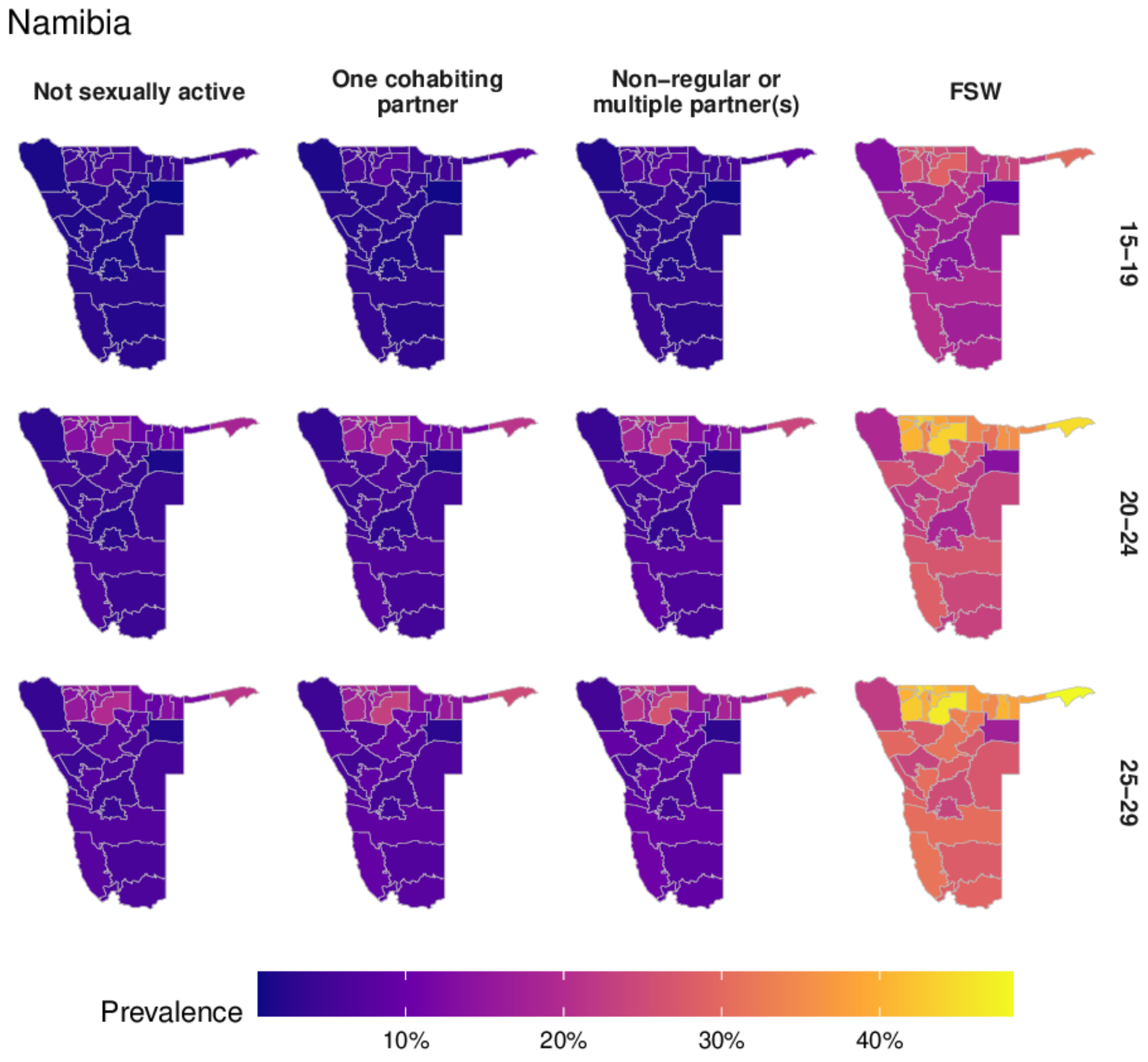

Figure X: District-level HIV prevalence for each of the risk groups in 2018 in Namibia.

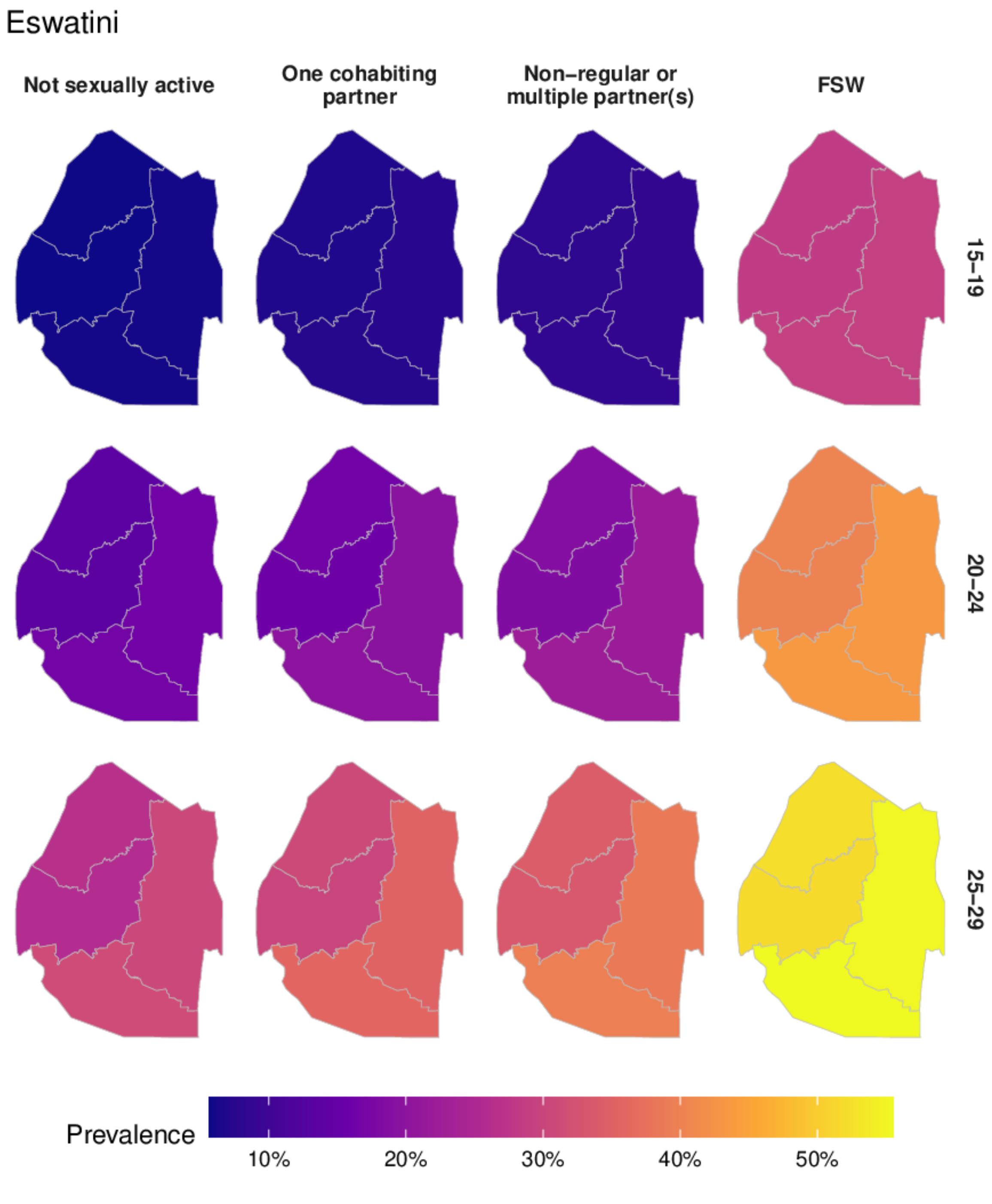

Figure Y: District-level HIV prevalence for each of the risk groups in 2018 in Eswatini.

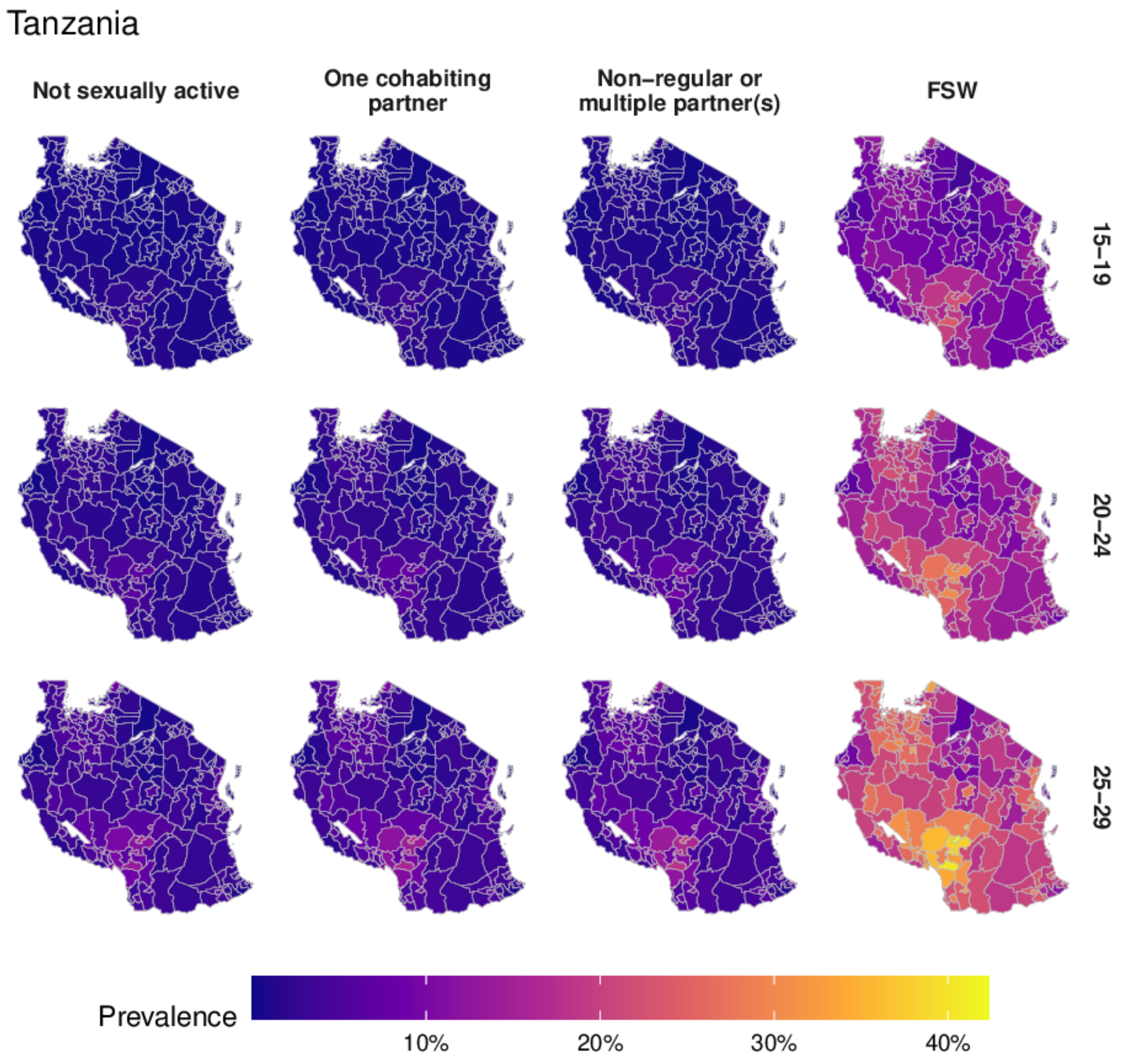

Figure Z: District-level HIV prevalence for each of the risk groups in 2018 in Tanzania.

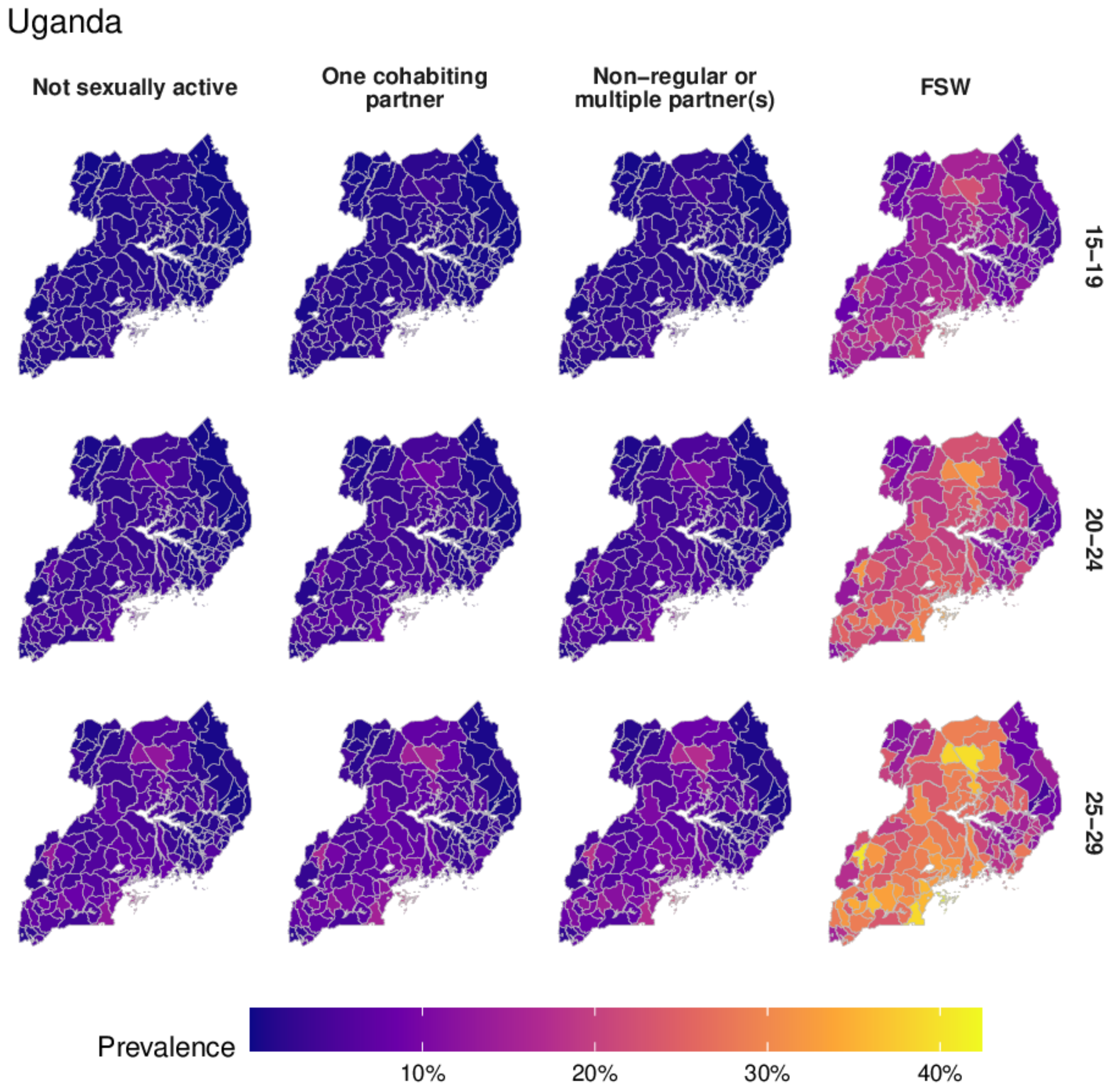

Figure AA: District-level HIV prevalence for each of the risk groups in 2018 in Uganda.

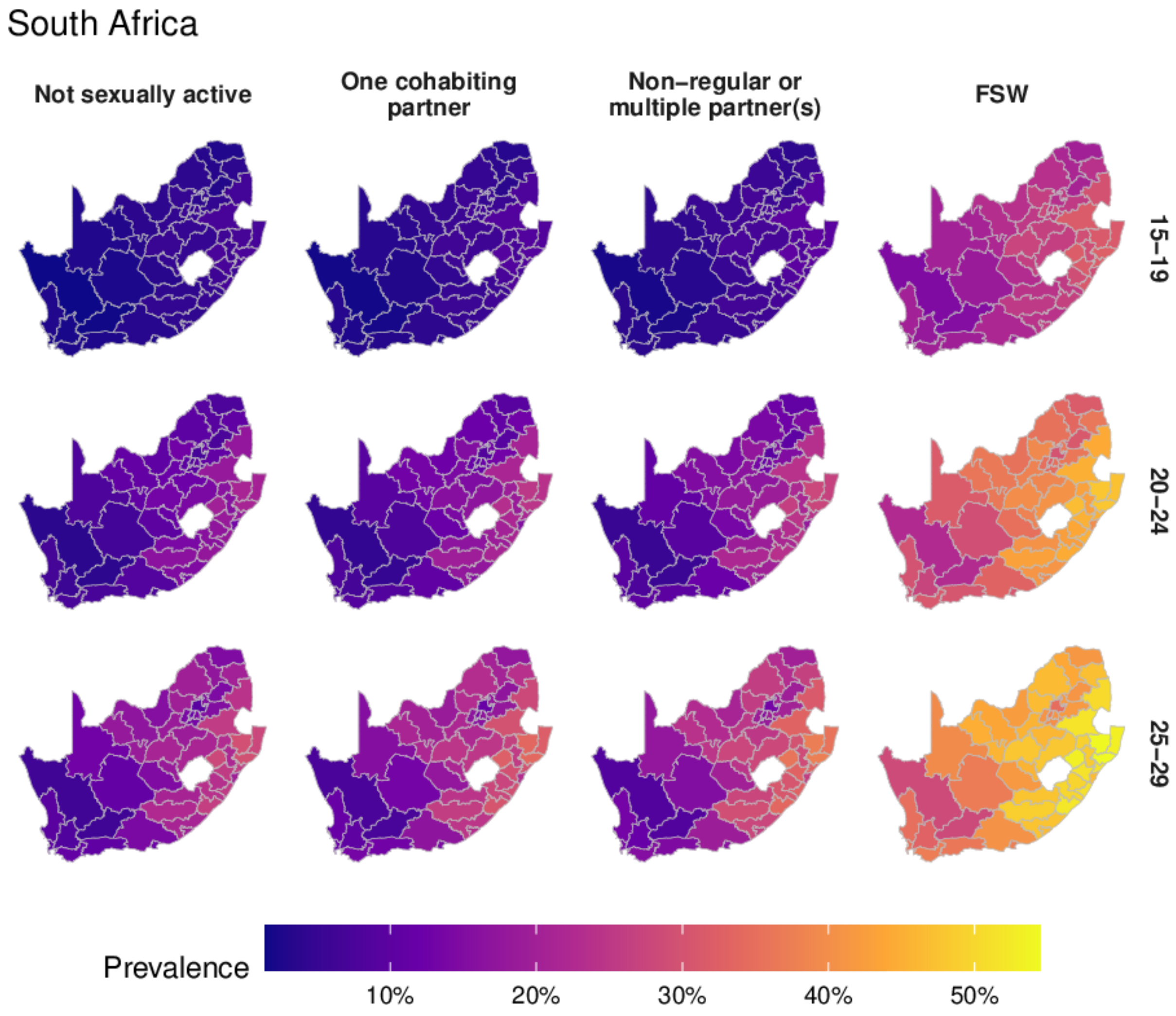

Figure AB: District-level HIV prevalence for each of the risk groups in 2018 in South Africa.

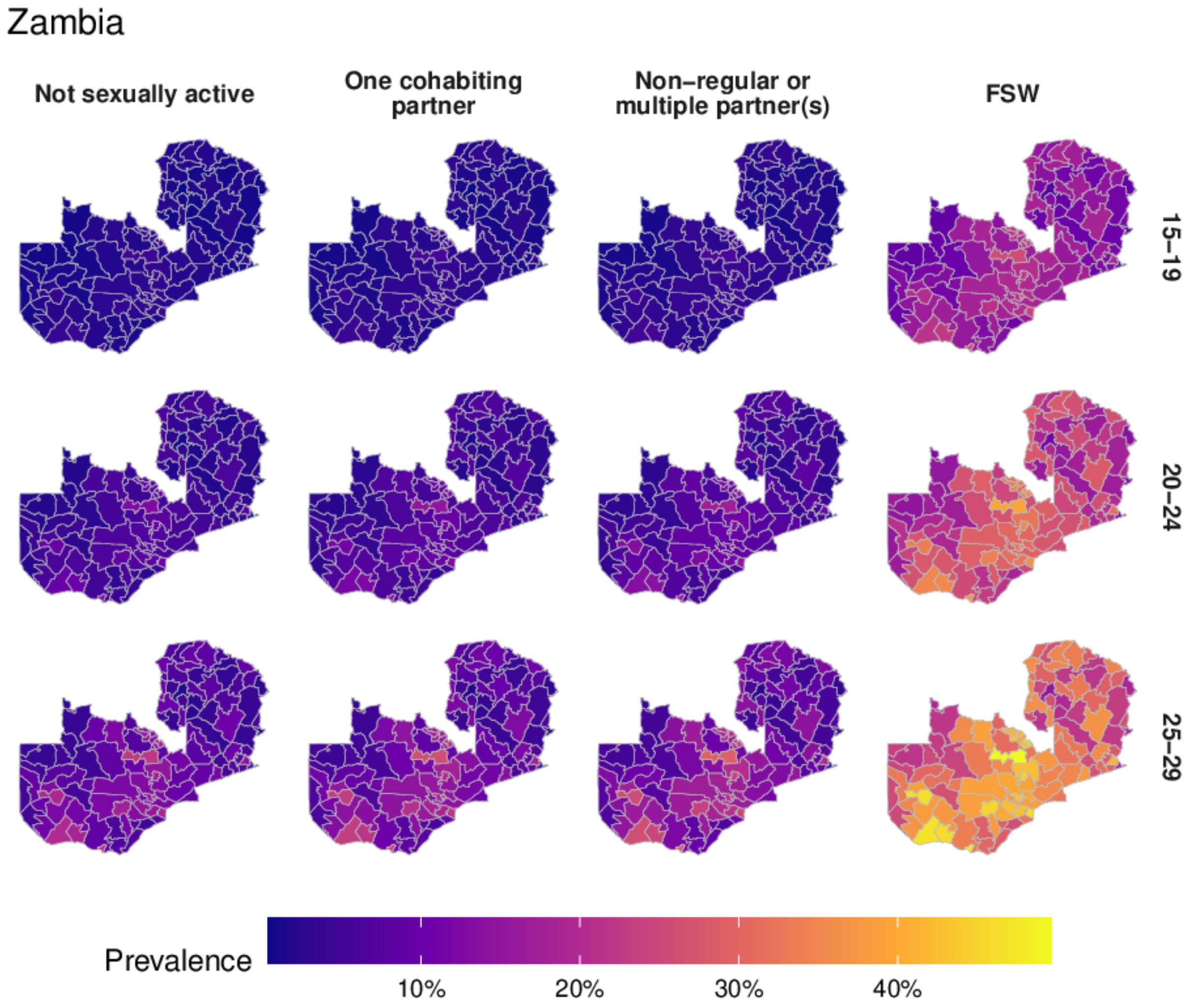

Figure AC: District-level HIV prevalence for each of the risk groups in 2018 in Zambia.

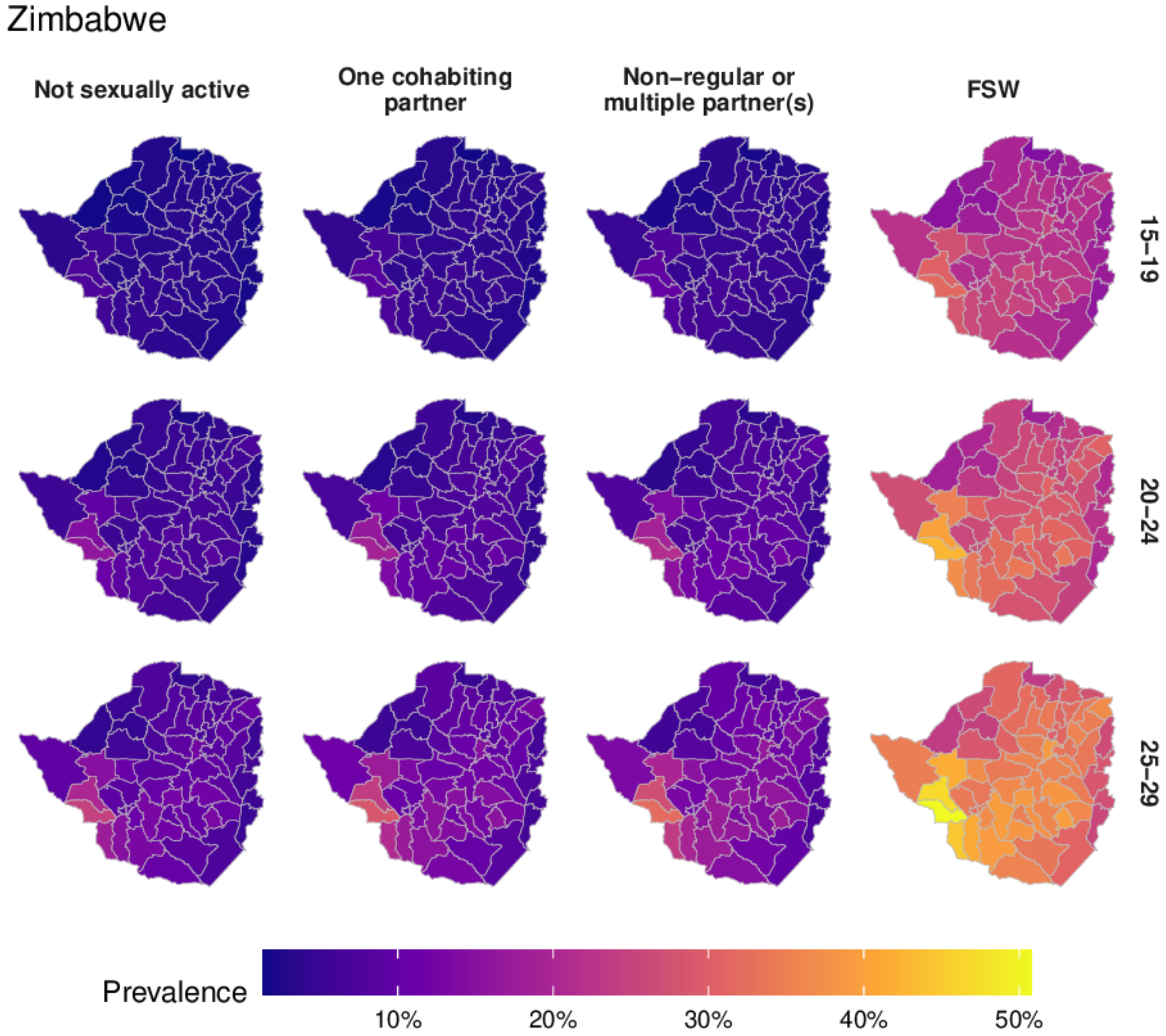

Figure AD: District-level HIV prevalence for each of the risk groups in 2018 in Zimbabwe.

**HIV incidence**

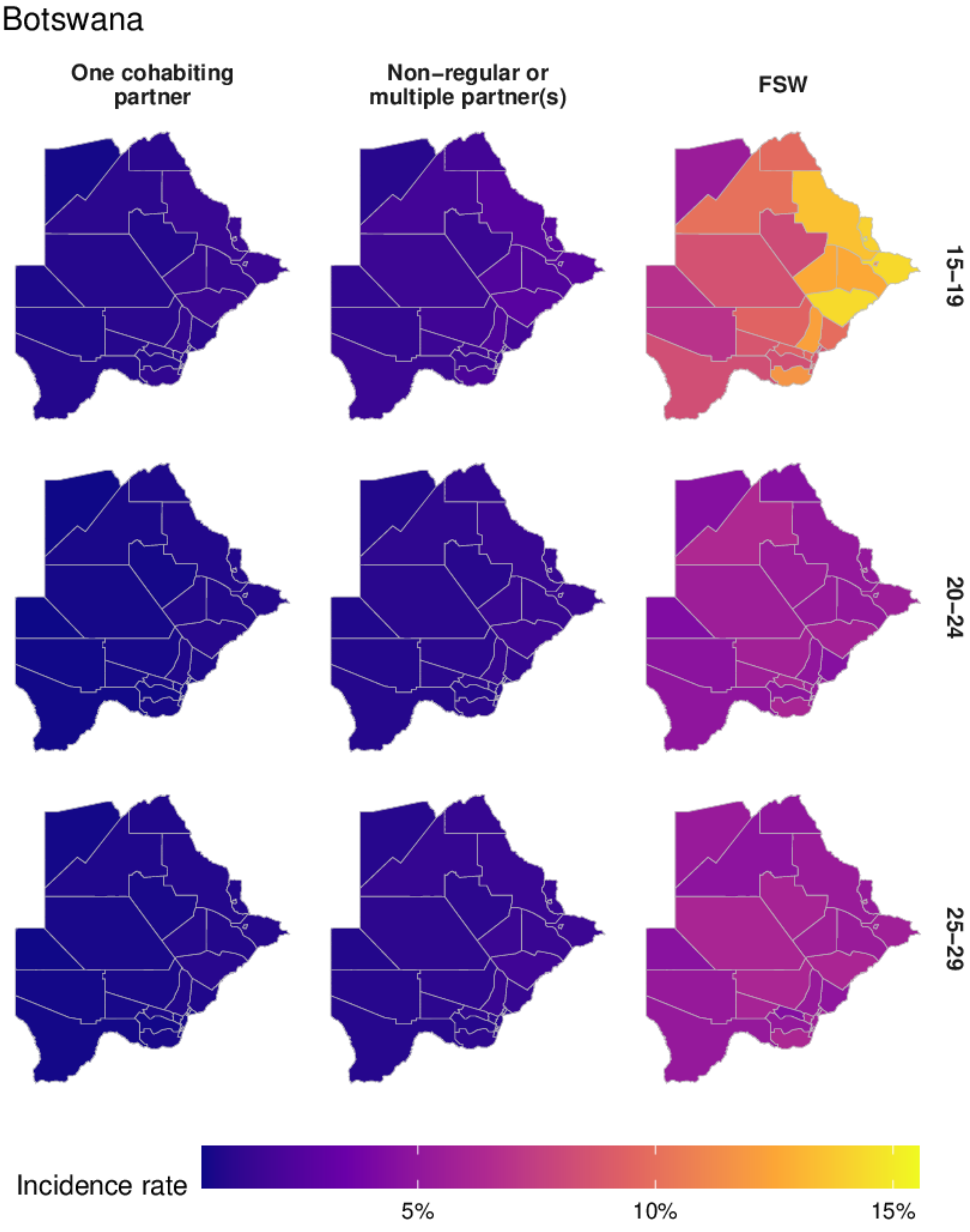

Figure AE: District-level HIV incidence for each of the risk groups in 2018 in Botswana.

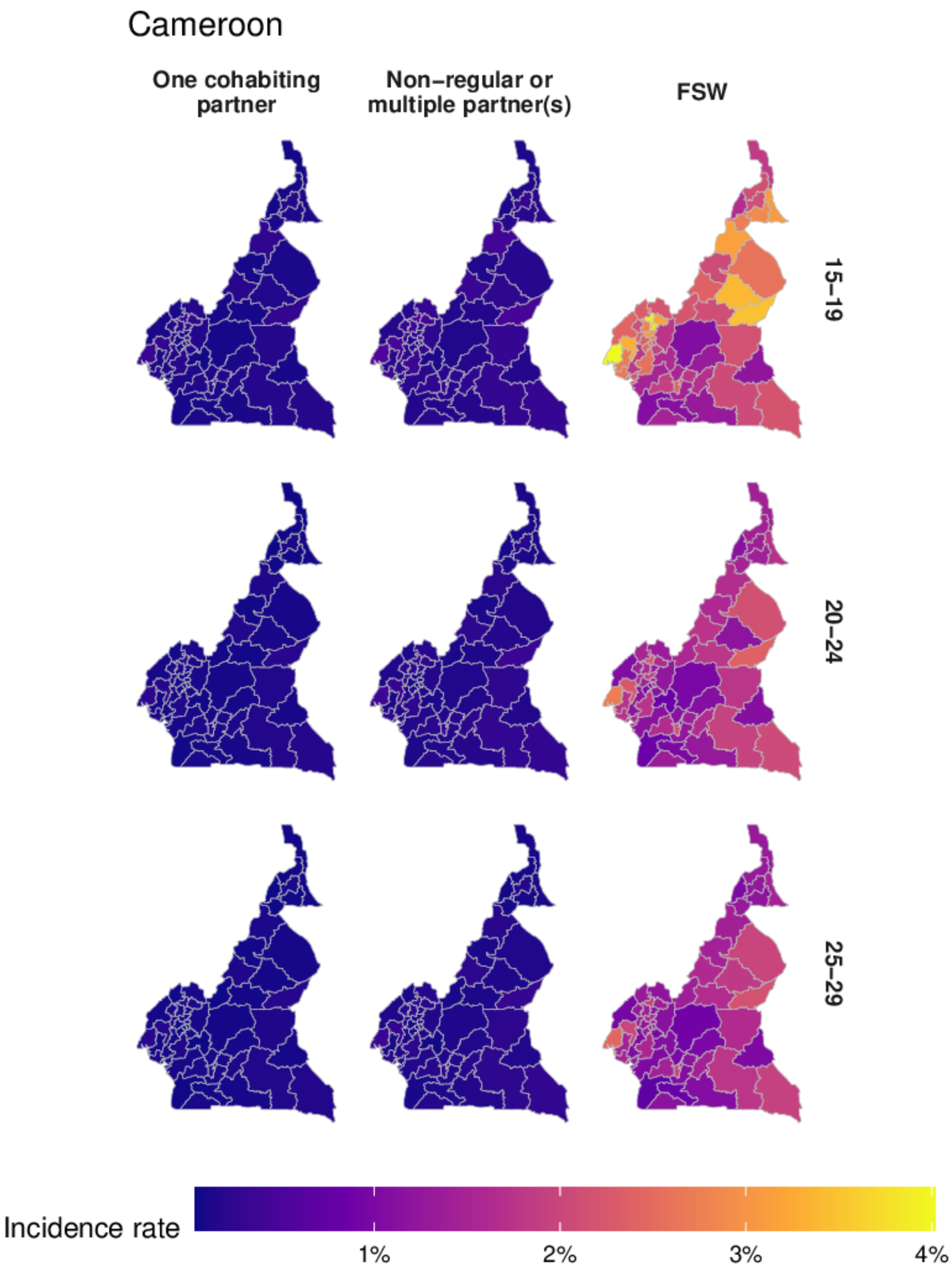

Figure AF: District-level HIV incidence for each of the risk groups in 2018 in Cameroon.

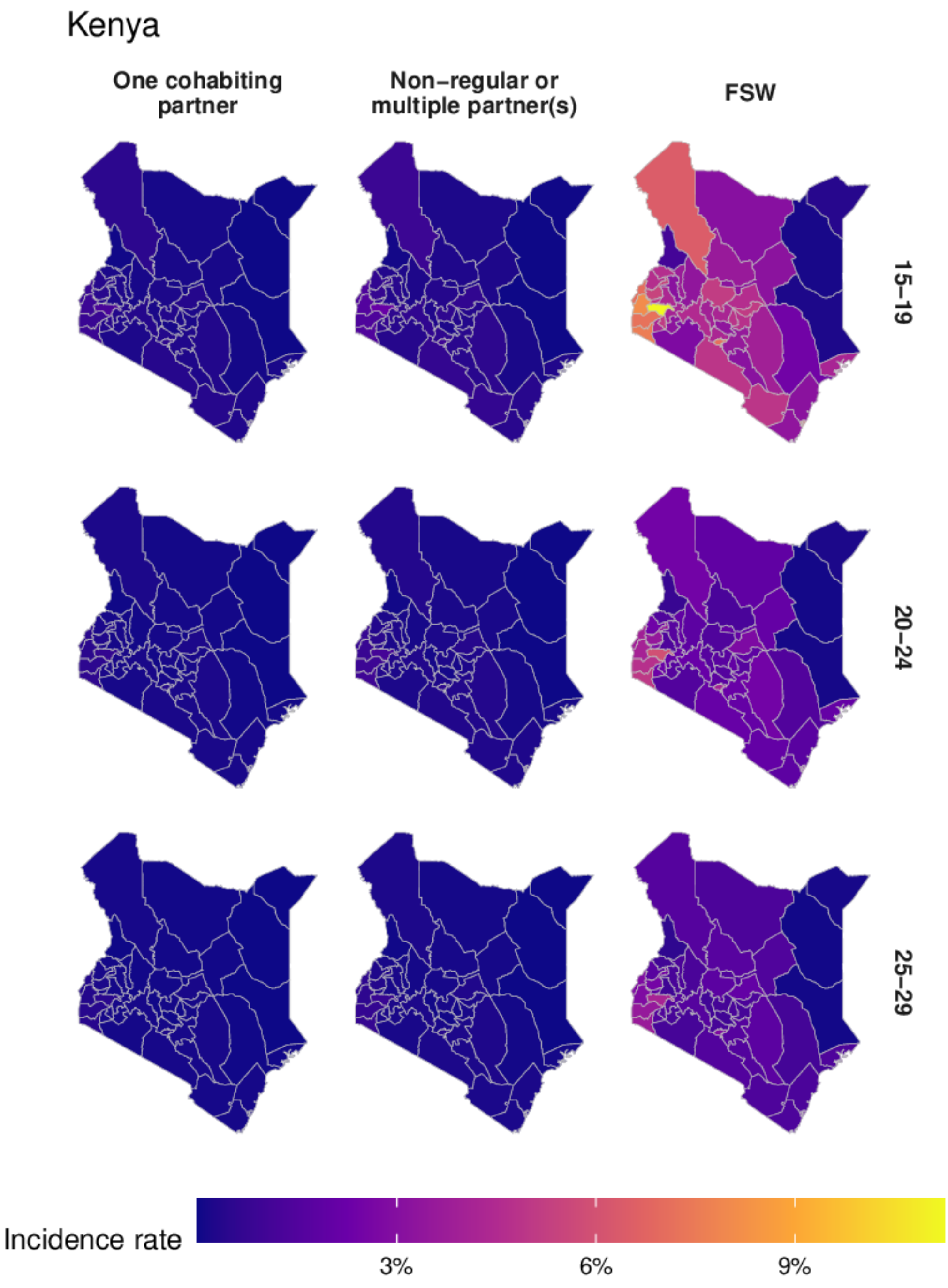

Figure AG: District-level HIV incidence for each of the risk groups in 2018 in Kenya.

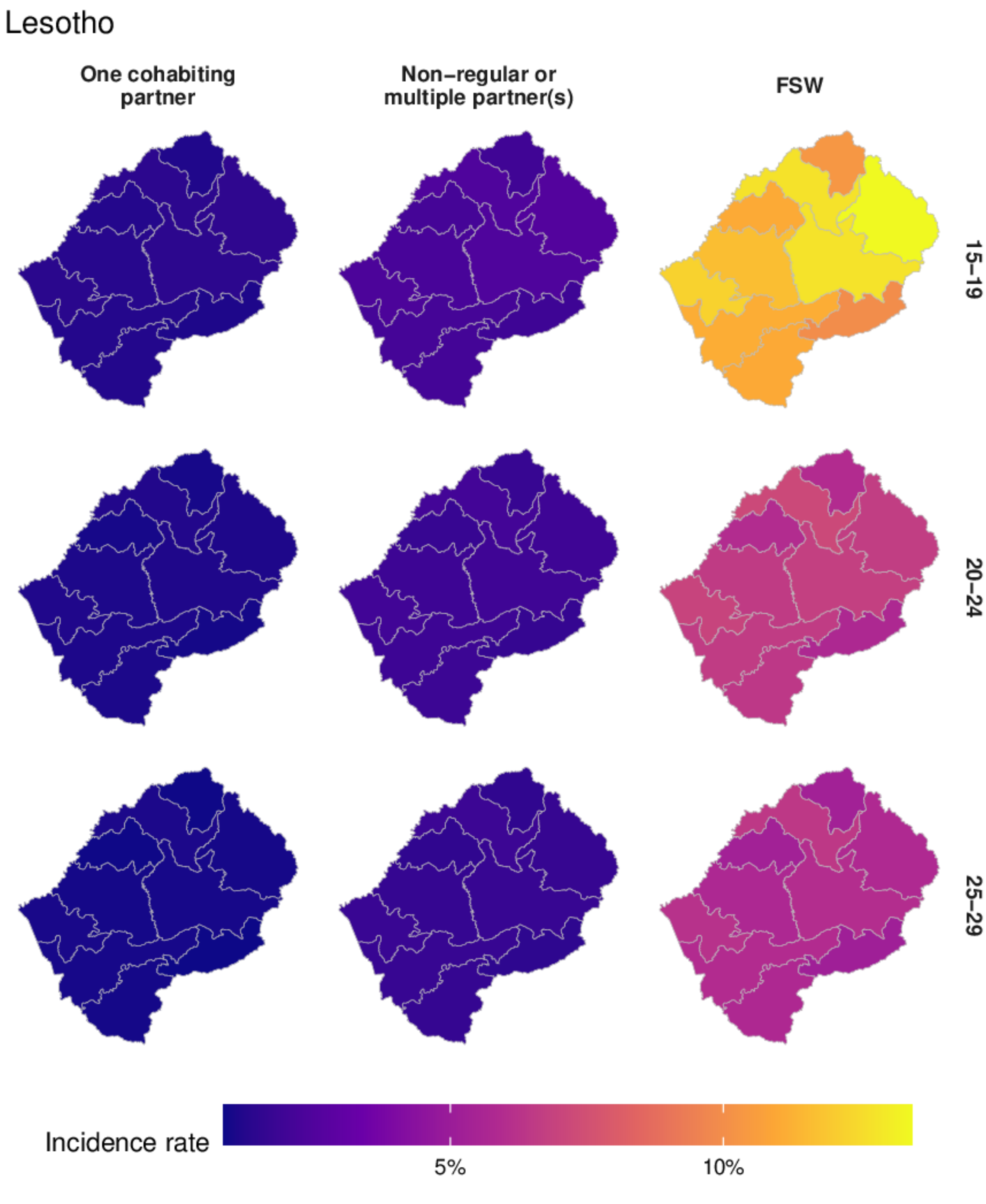

Figure AH: District-level HIV incidence for each of the risk groups in 2018 in Lesotho.

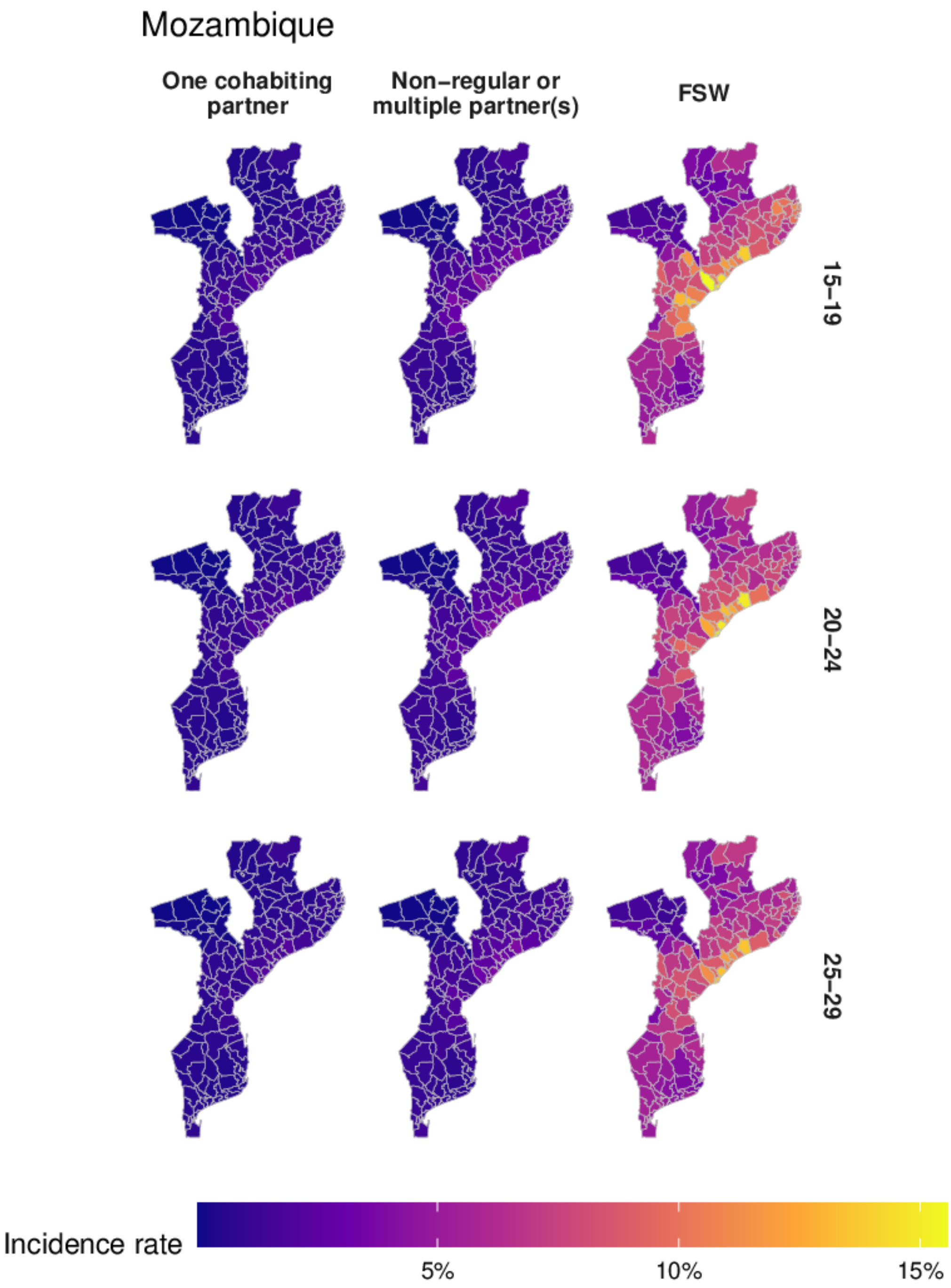

Figure AI: District-level HIV incidence for each of the risk groups in 2018 in Mozambique.

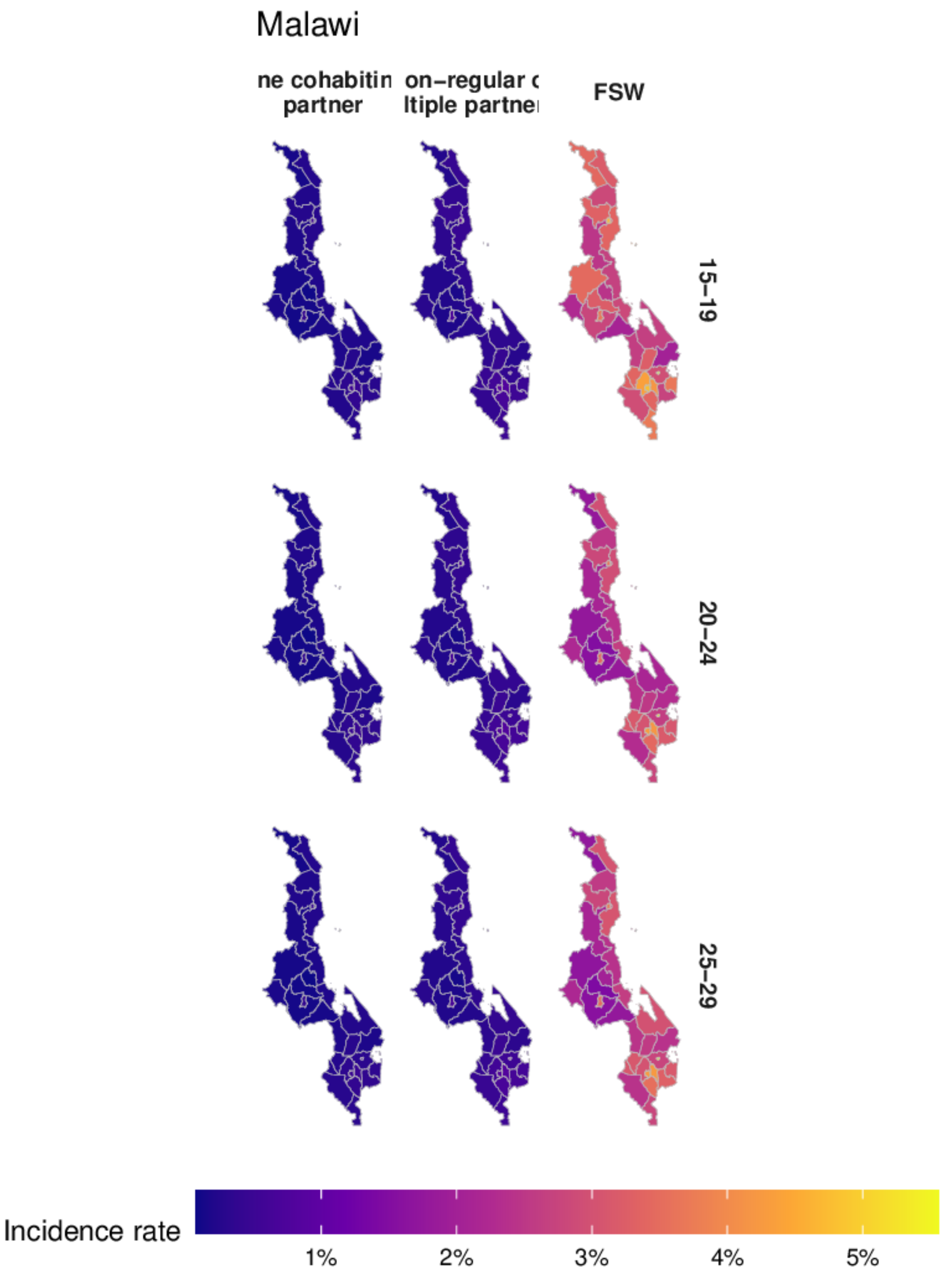

Figure AJ: District-level HIV incidence for each of the risk groups in 2018 in Malawi.

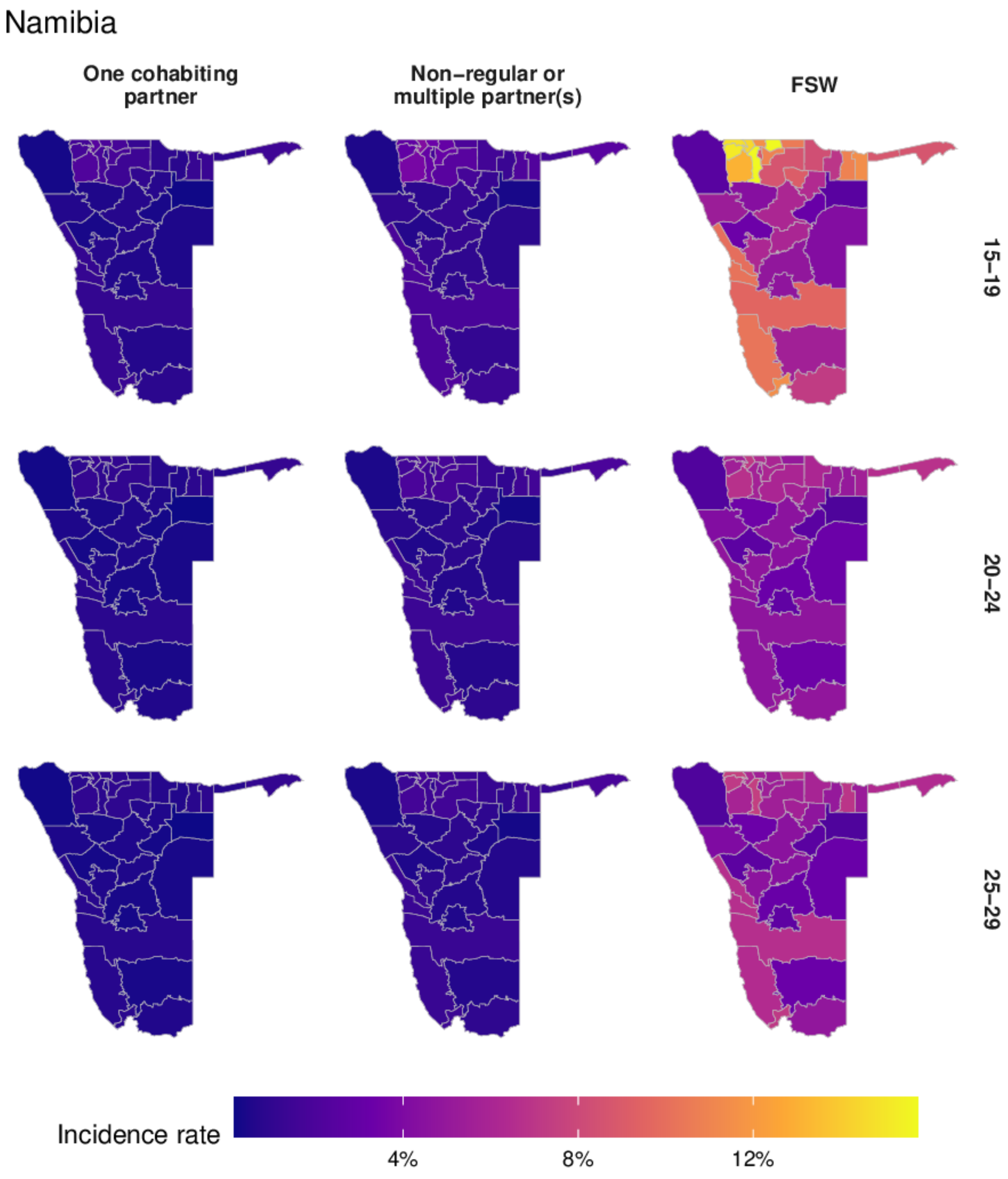

Figure AK: District-level HIV incidence for each of the risk groups in 2018 in Namibia.

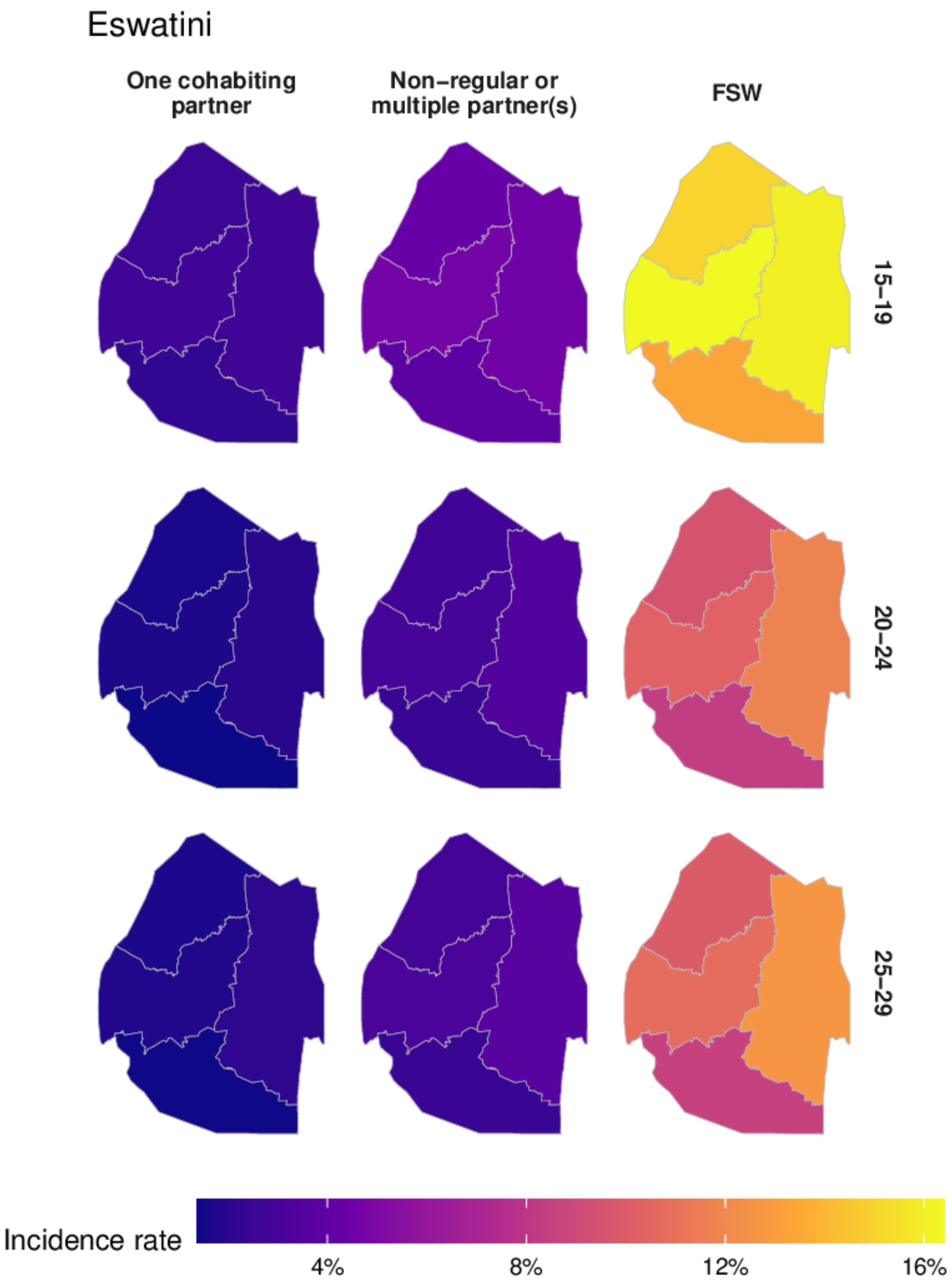

Figure AL: District-level HIV incidence for each of the risk groups in 2018 in Eswatini.

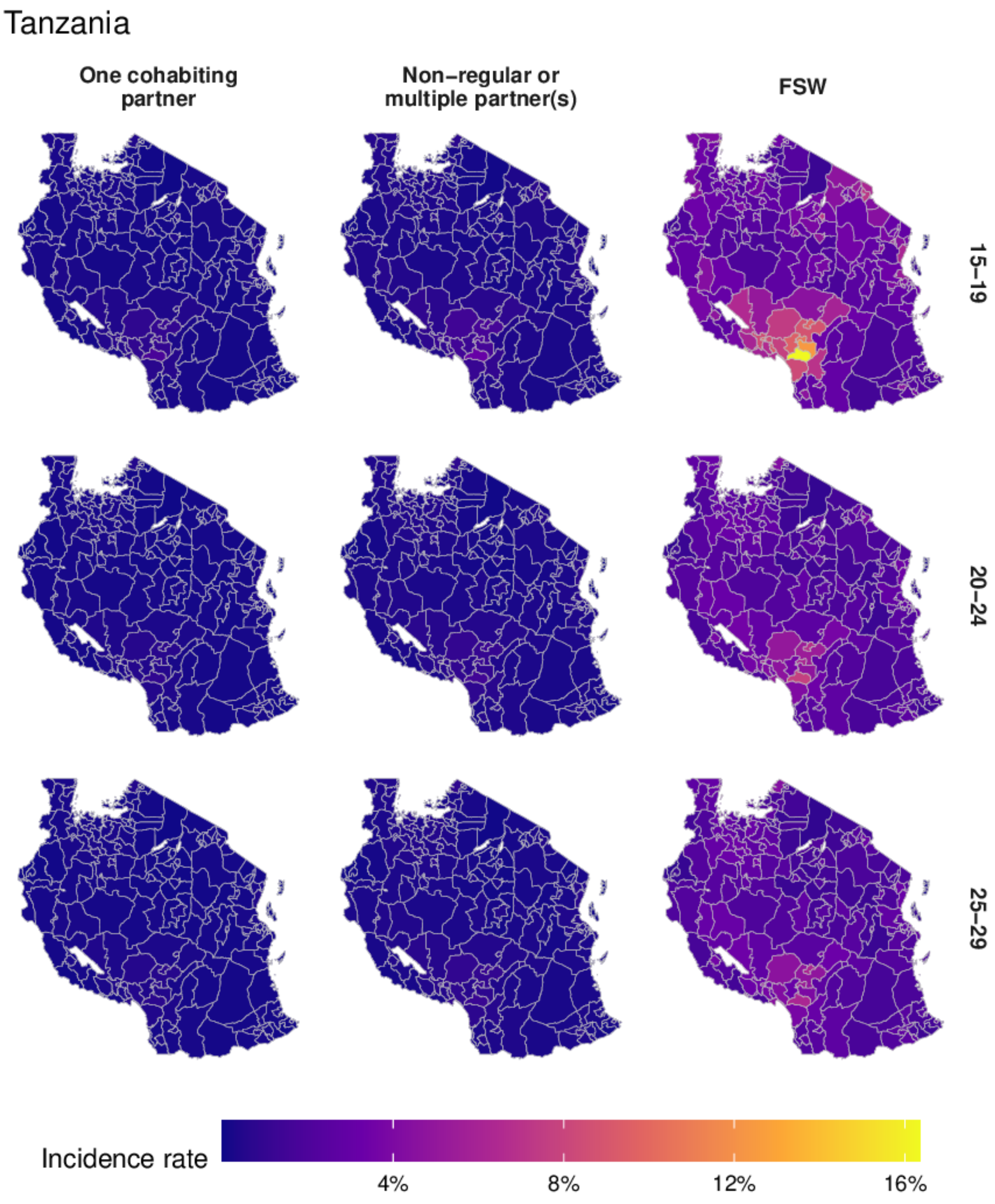

Figure AM: District-level HIV incidence for each of the risk groups in 2018 in Tanzania.

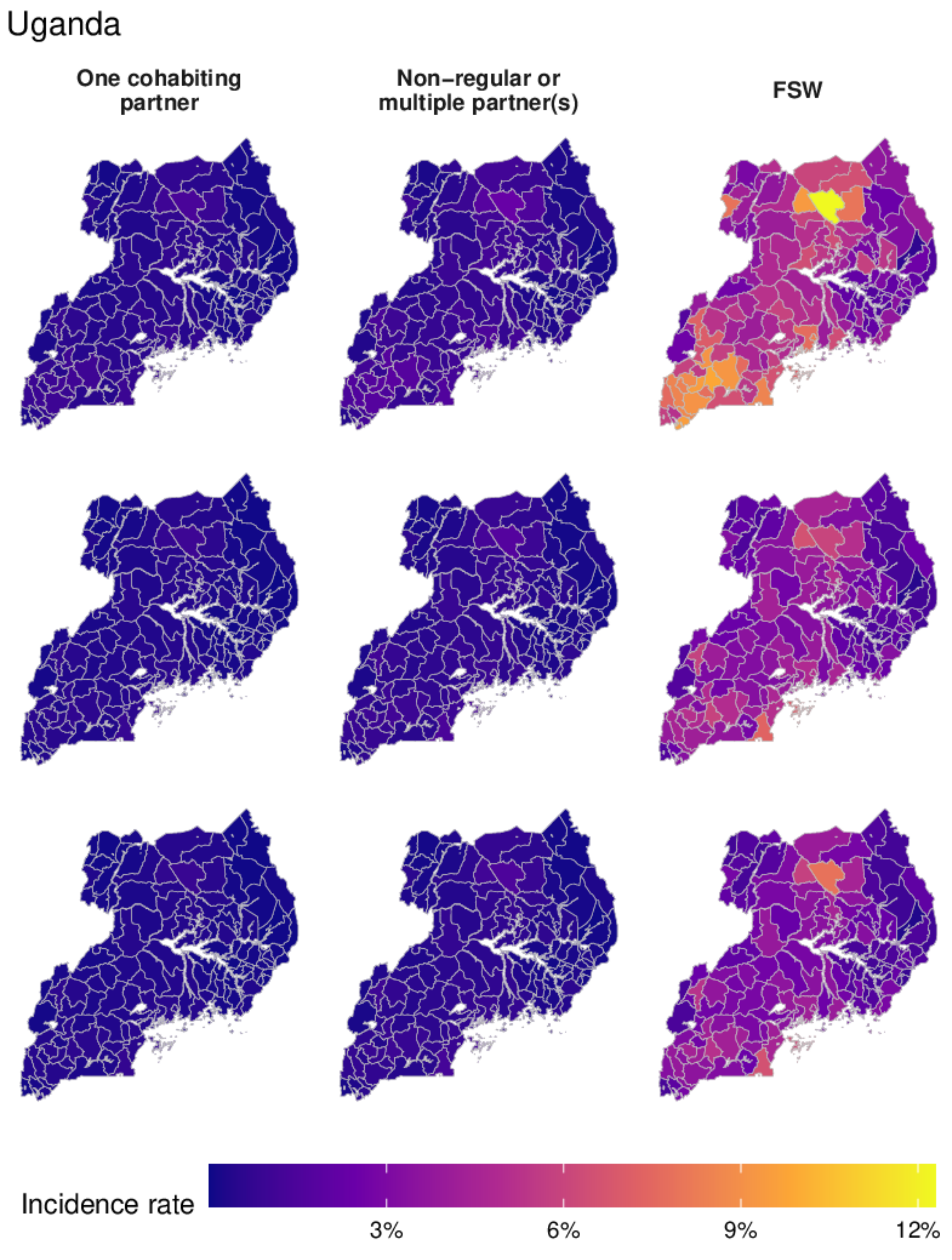

Figure AN: District-level HIV incidence for each of the risk groups in 2018 in Uganda.

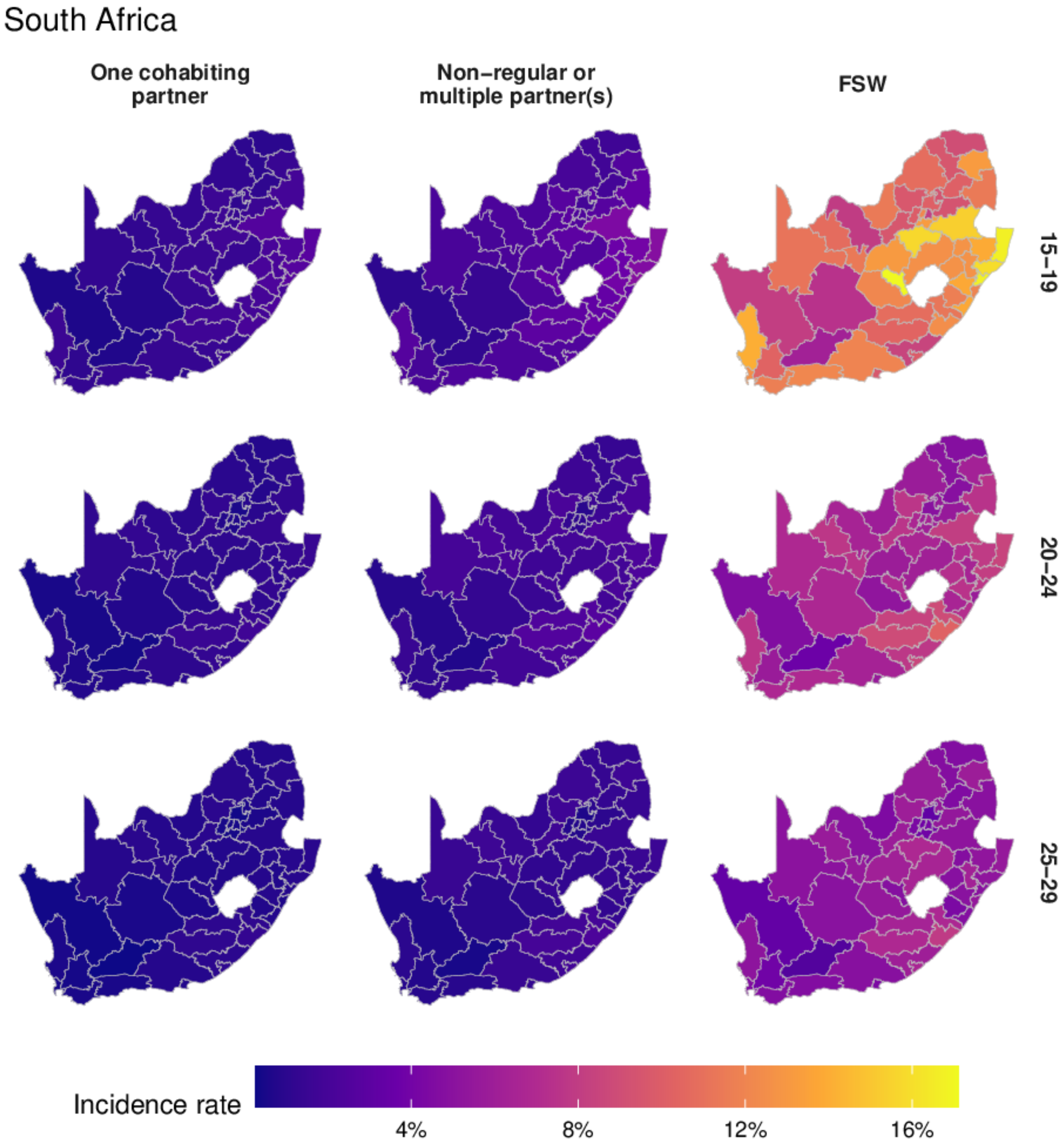

Figure AO: District-level HIV incidence for each of the risk groups in 2018 in South Africa.

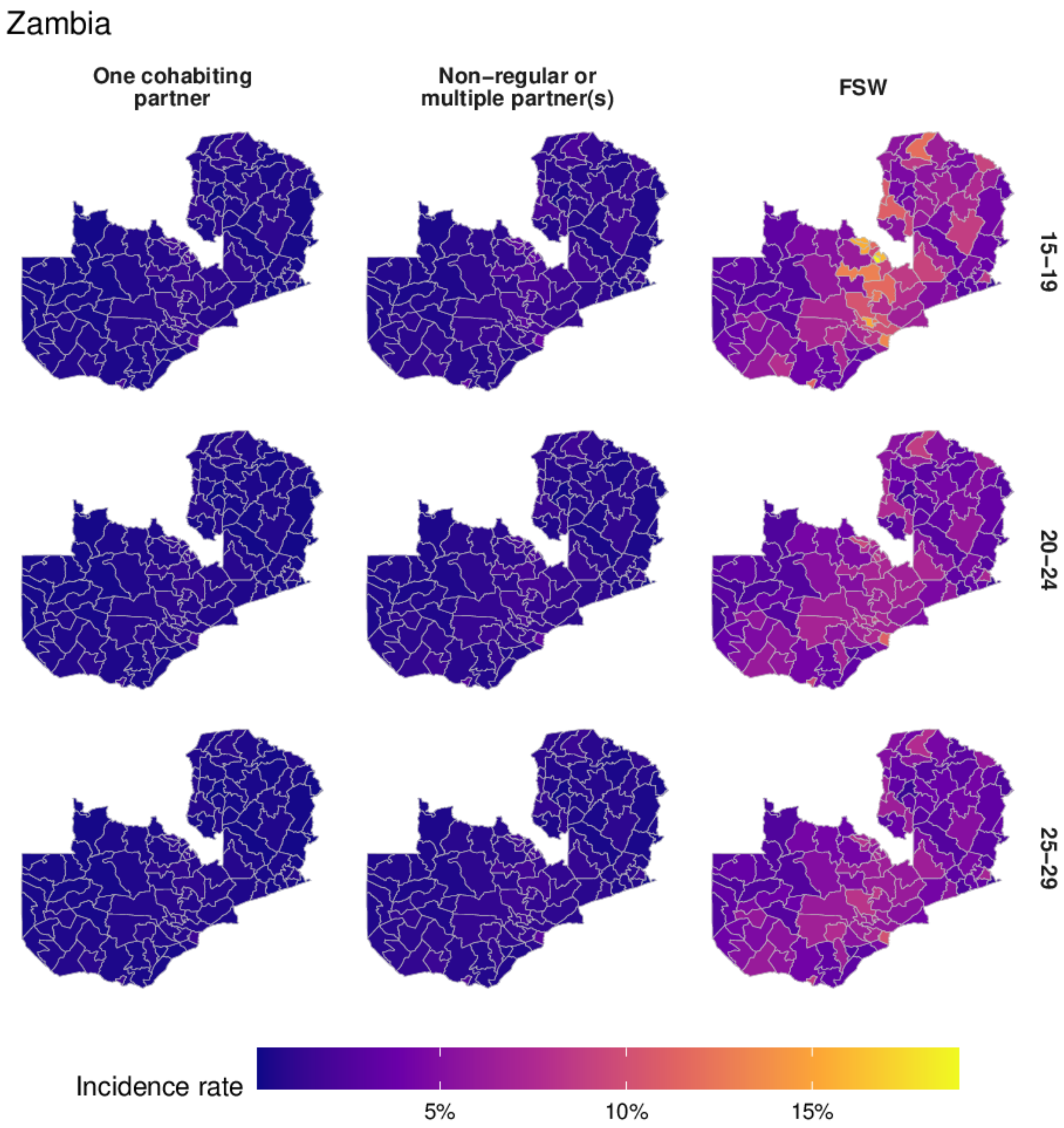

Figure AP: District-level HIV incidence for each of the risk groups in 2018 in Zambia.

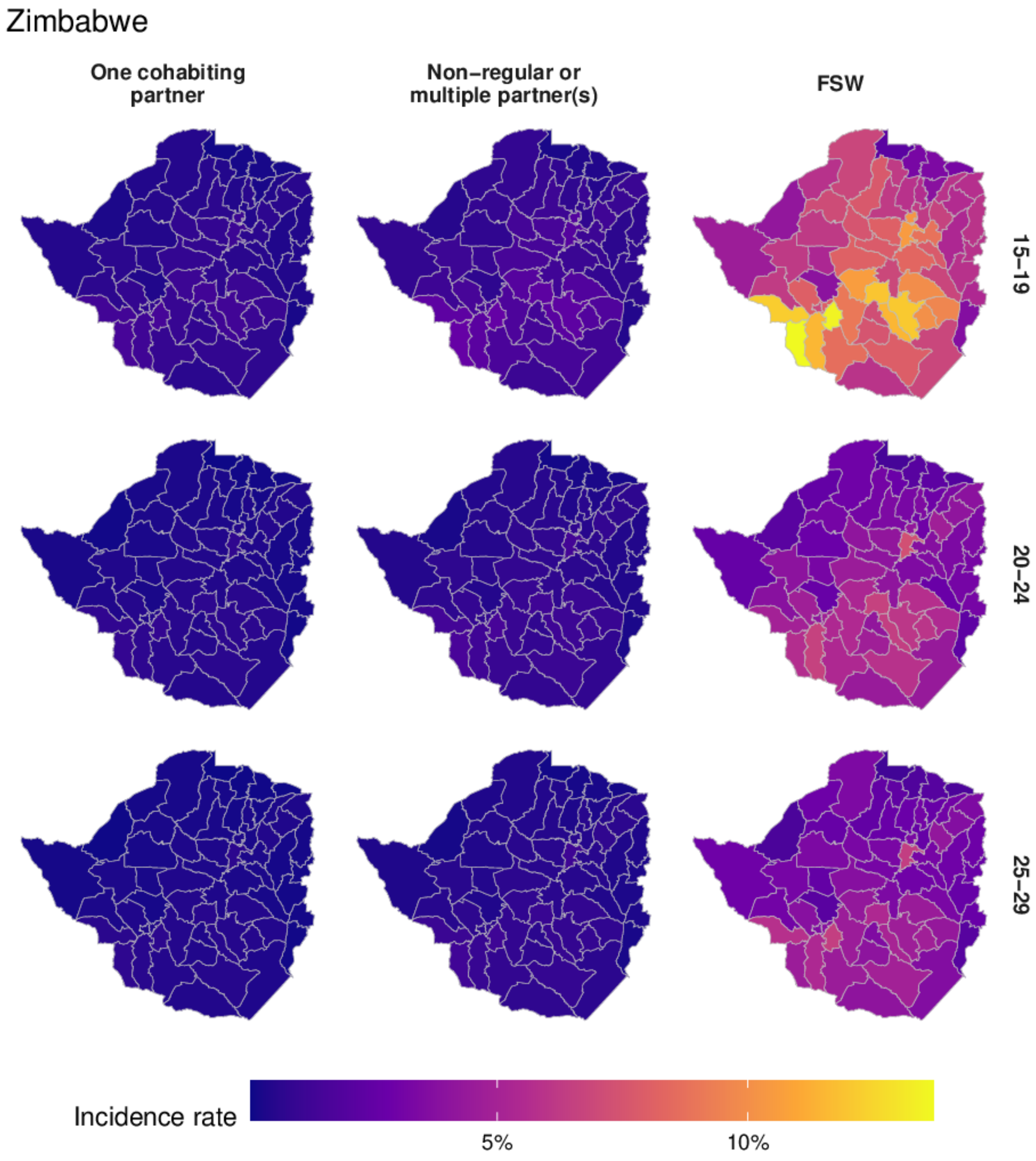

Figure AQ: District-level HIV incidence for each of the risk groups in 2018 in Zimbabwe.

**Expected new infections reached**

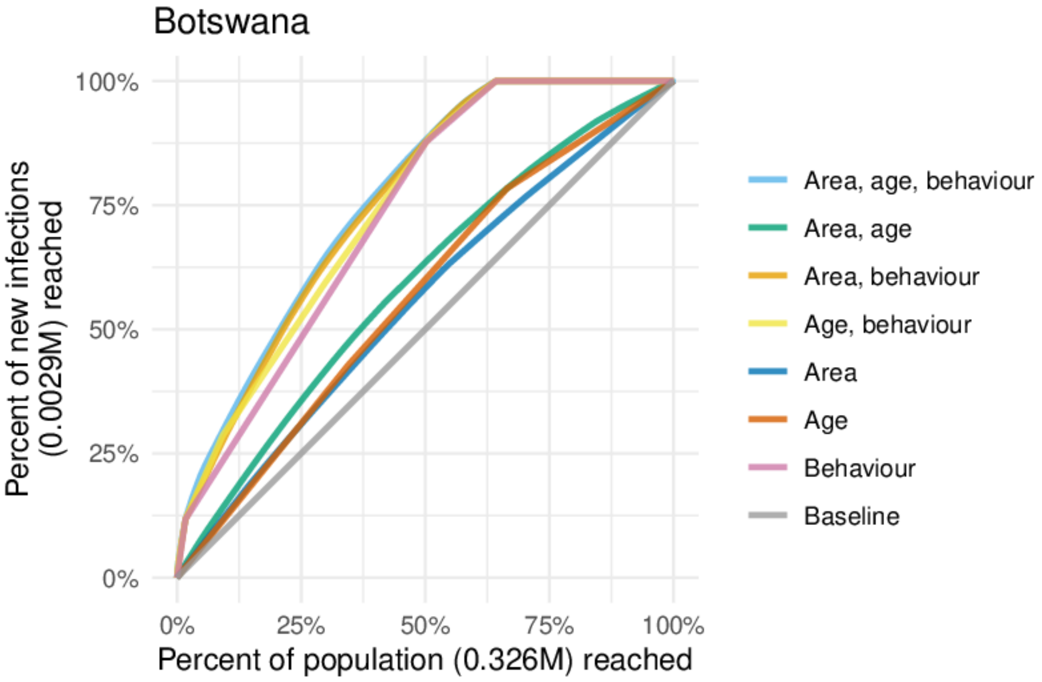

Figure AR: Percentage of expected new infections reached taking a variety of risk stratification approaches against the percentage of at risk population reached in Botswana.

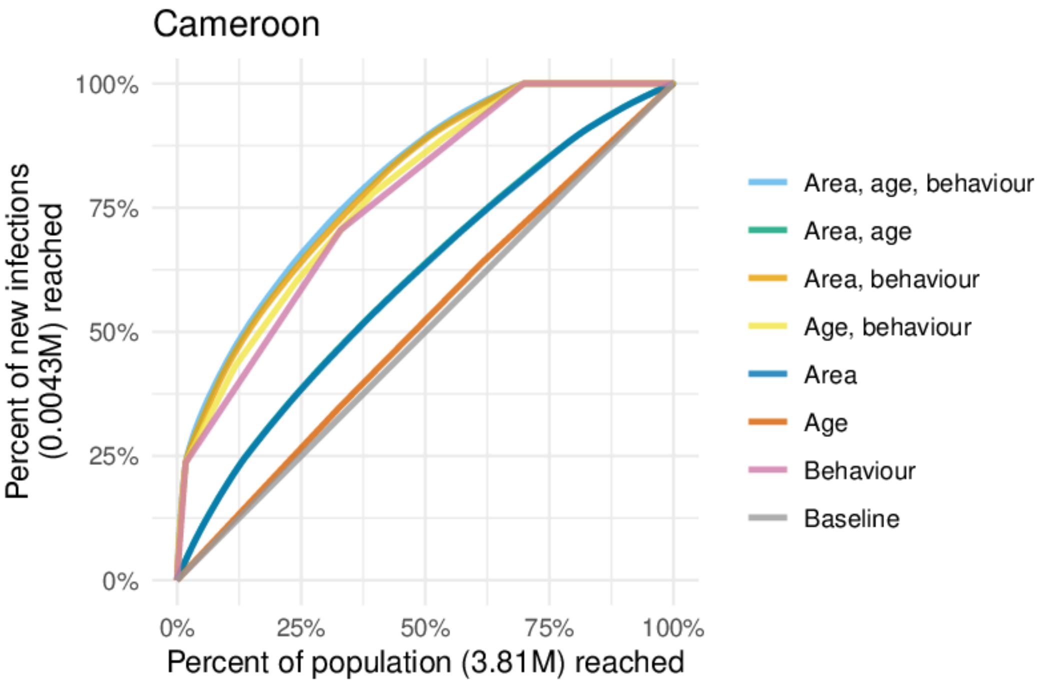

Figure AS: Percentage of expected new infections reached taking a variety of risk stratification approaches against the percentage of at risk population reached in Cameroon.

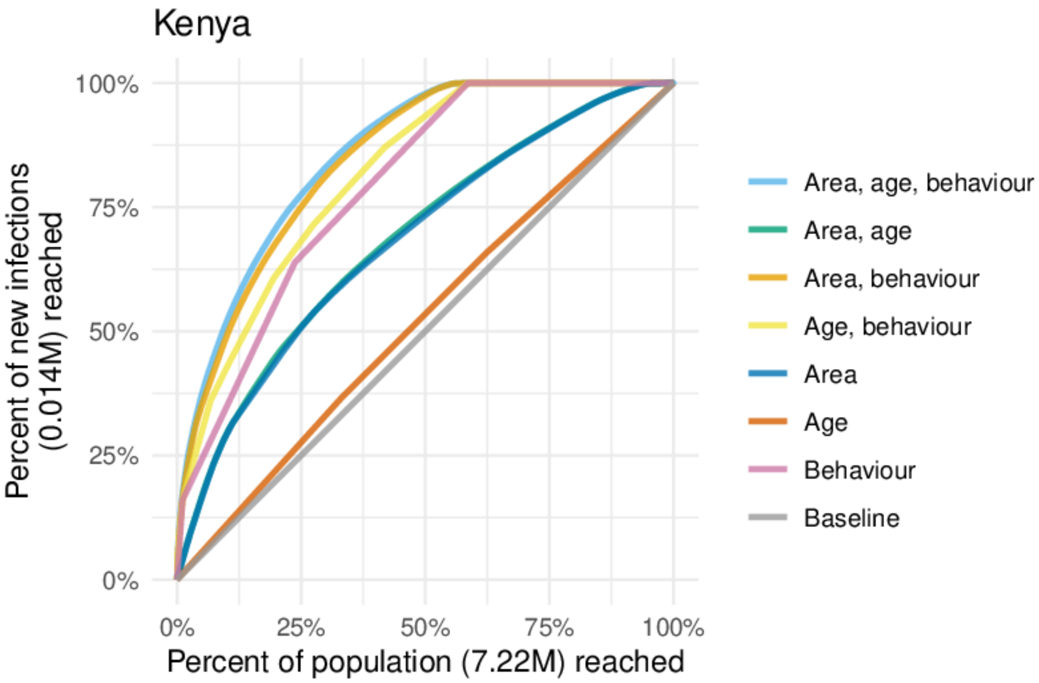

Figure AT: Percentage of expected new infections reached taking a variety of risk stratification approaches against the percentage of at risk population reached in Kenya.

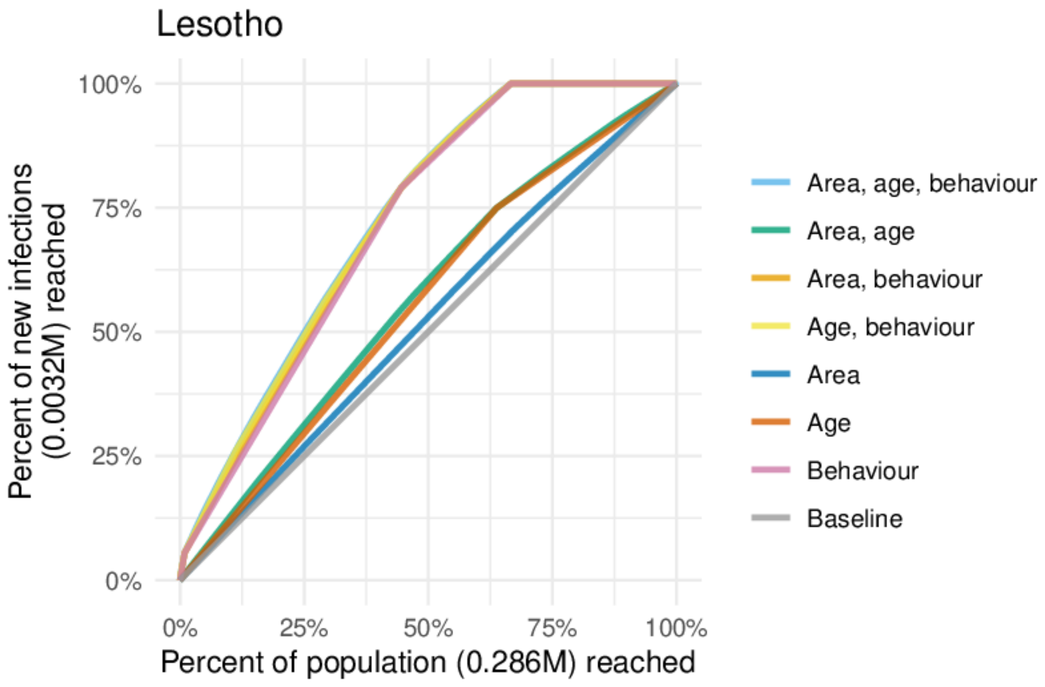

Figure AU: Percentage of expected new infections reached taking a variety of risk stratification approaches against the percentage of at risk population reached in Lesotho.

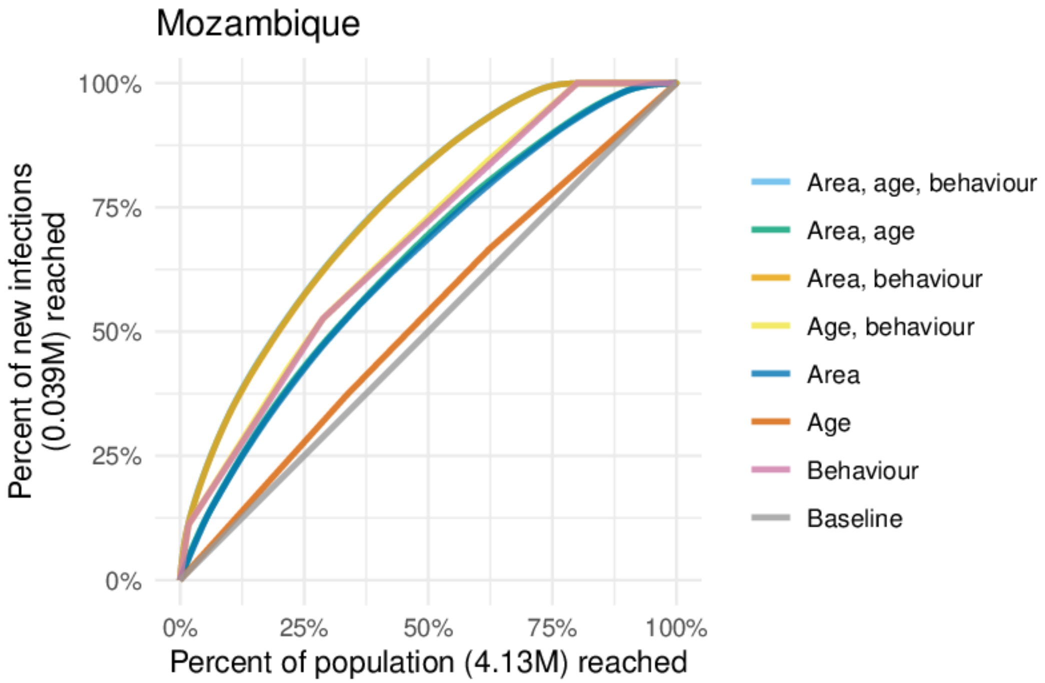

Figure AV: Percentage of expected new infections reached taking a variety of risk stratification approaches against the percentage of at risk population reached in Mozambique.

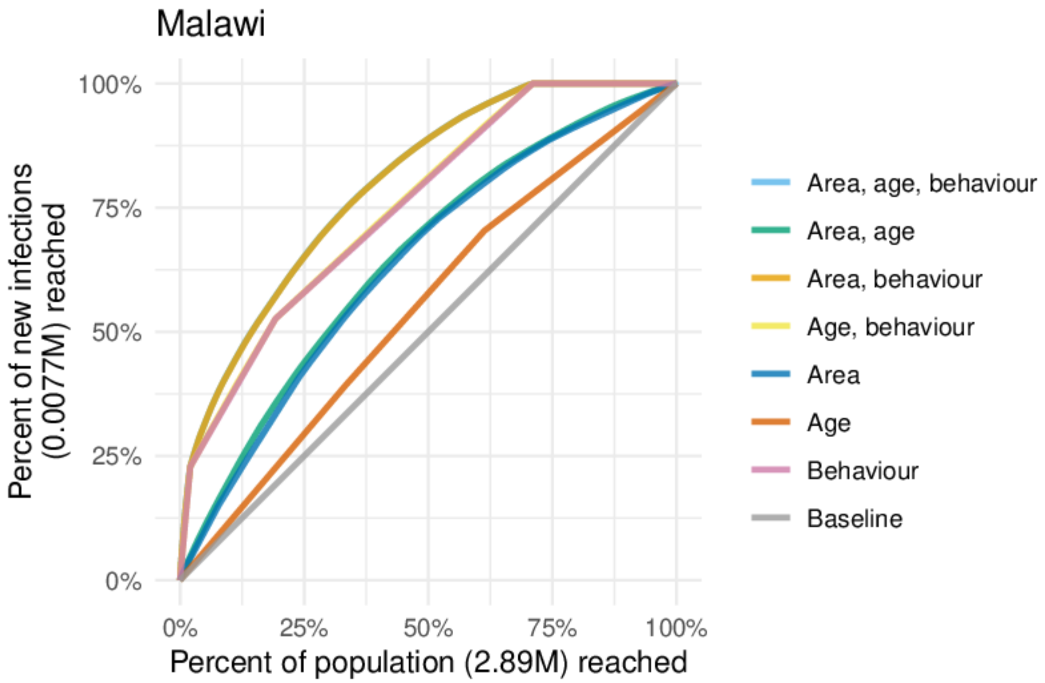

Figure AW: Percentage of expected new infections reached taking a variety of risk stratification approaches against the percentage of at risk population reached in Malawi.

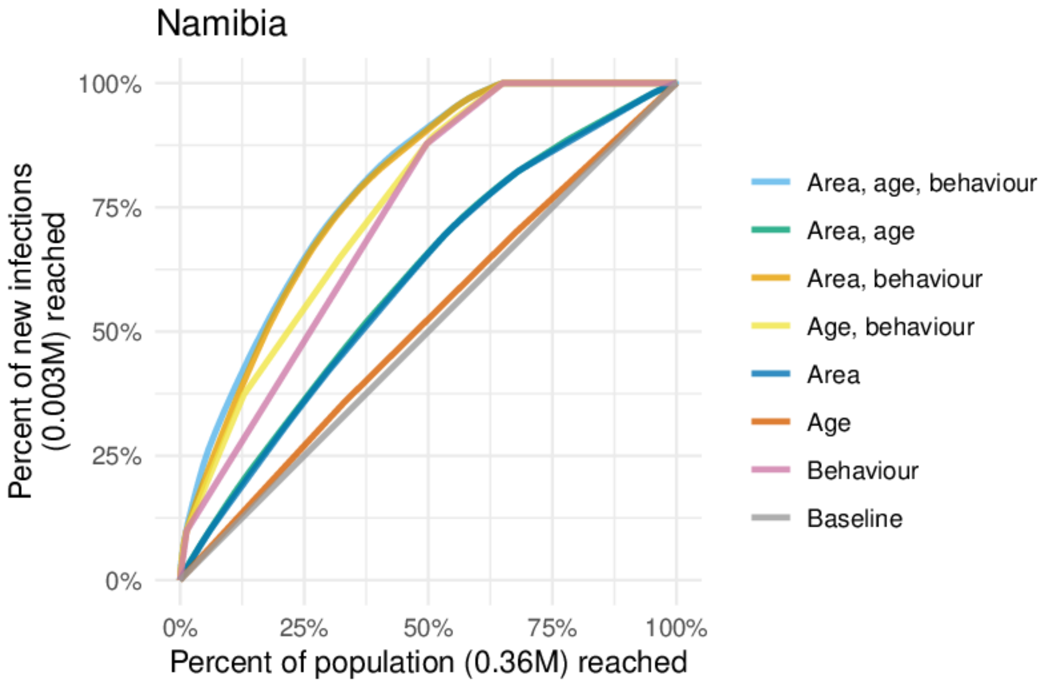

Figure AX: Percentage of expected new infections reached taking a variety of risk stratification approaches against the percentage of at risk population reached in Namibia.

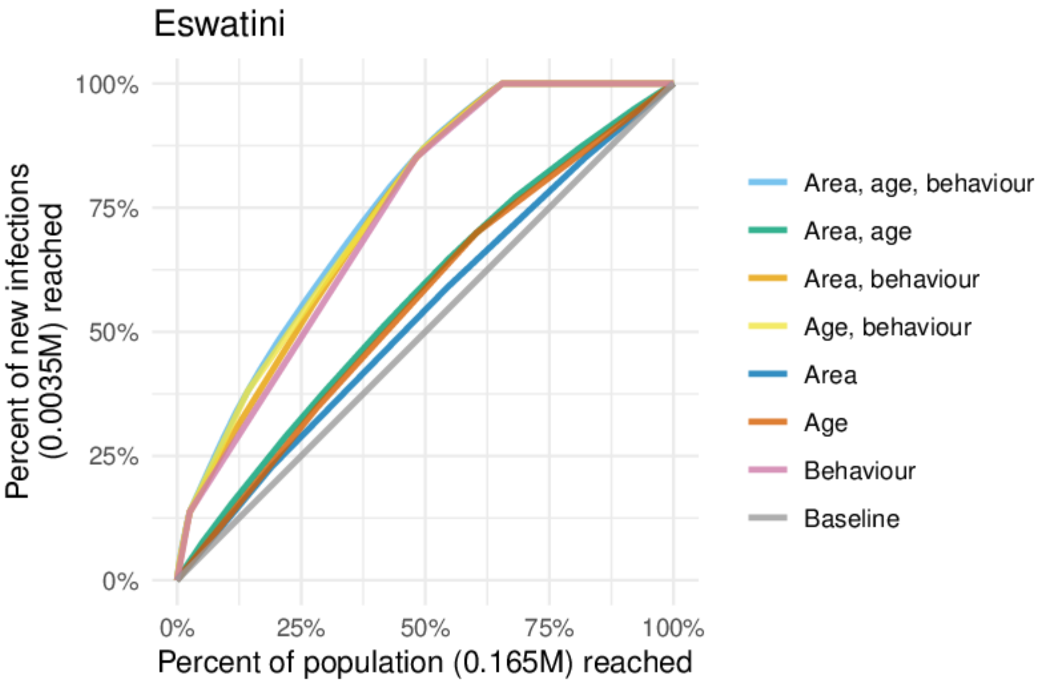

Figure AY: Percentage of expected new infections reached taking a variety of risk stratification approaches against the percentage of at risk population reached in Eswatini.

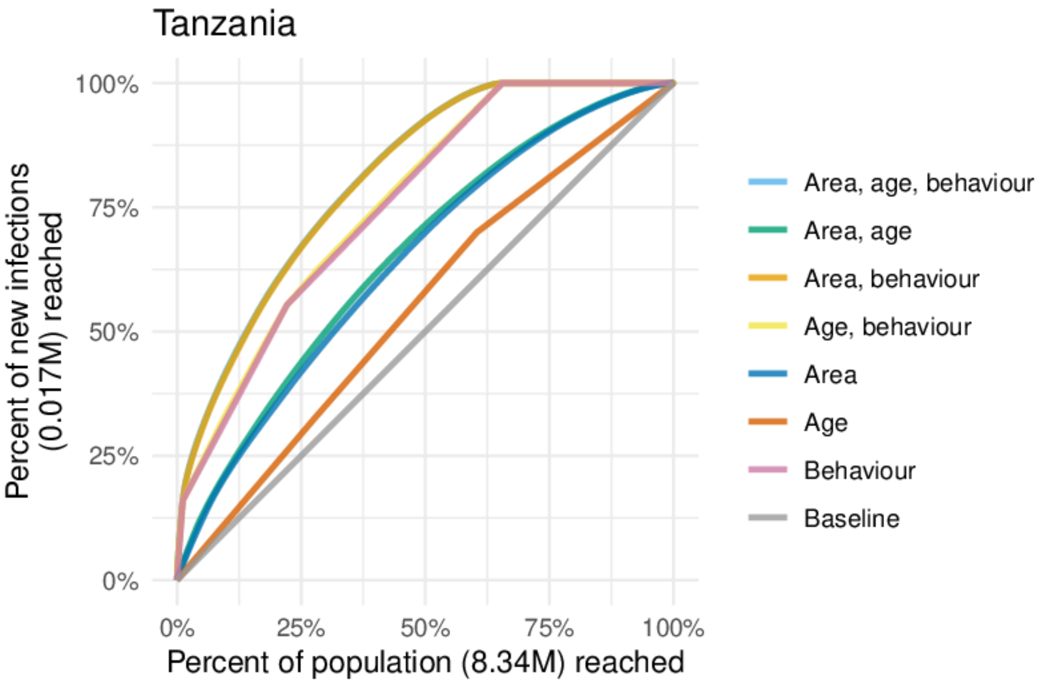

Figure AZ: Percentage of expected new infections reached taking a variety of risk stratification approaches against the percentage of at risk population reached in Tanzania.

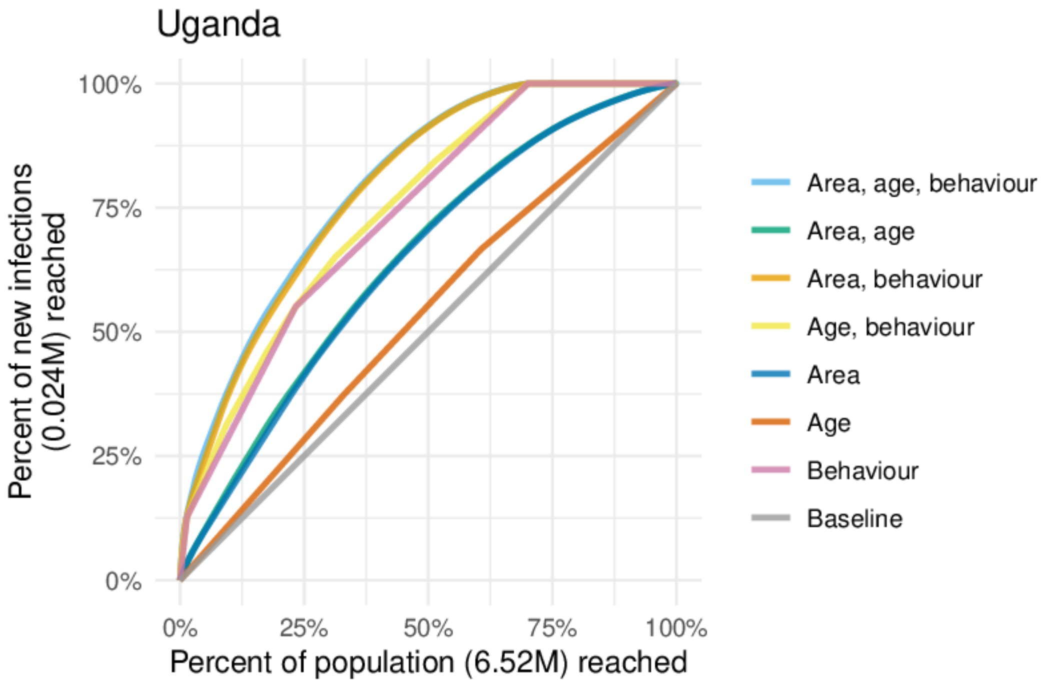

Figure BA: Percentage of expected new infections reached taking a variety of risk stratification approaches against the percentage of at risk population reached in Uganda.

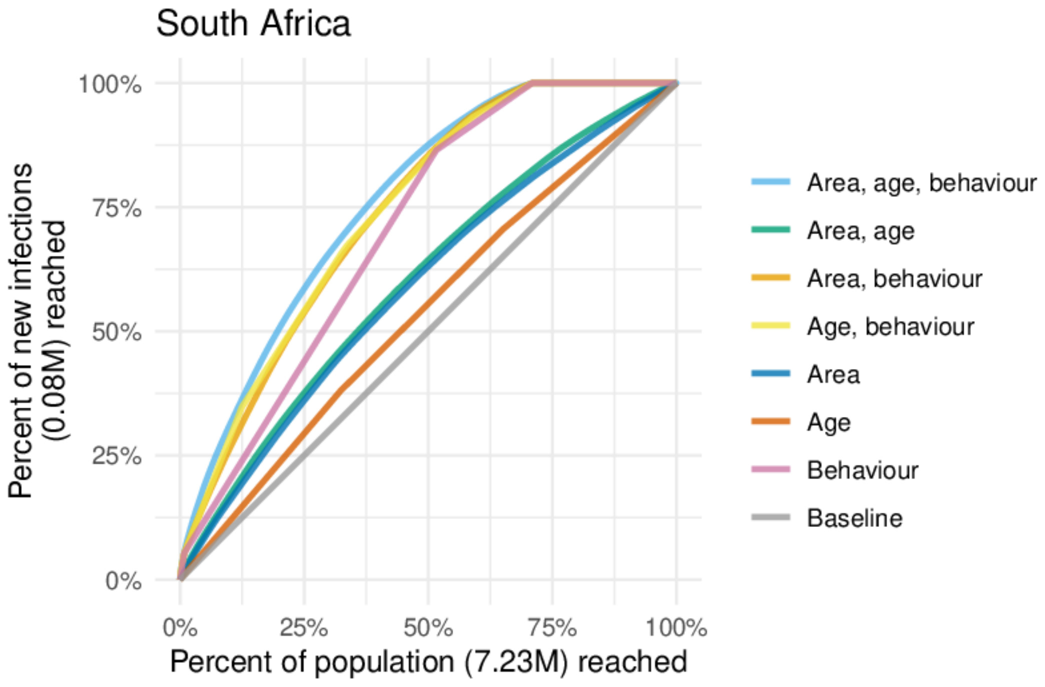

Figure BB: Percentage of expected new infections reached taking a variety of risk stratification approaches against the percentage of at risk population reached in South Africa.

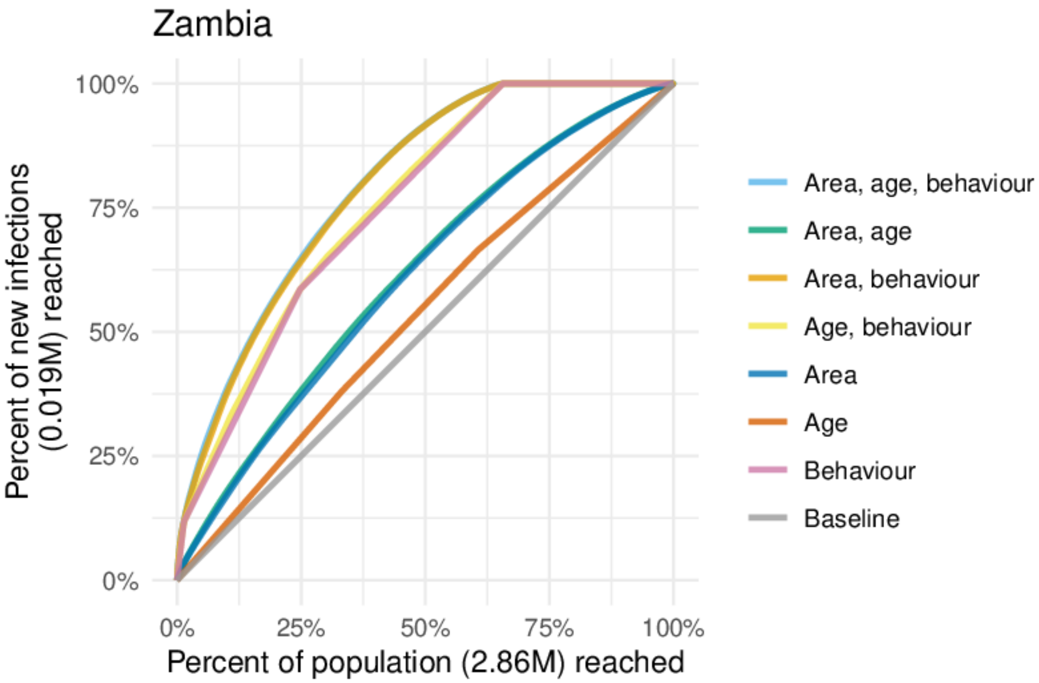

Figure BC: Percentage of expected new infections reached taking a variety of risk stratification approaches against the percentage of at risk population reached in Zambia.

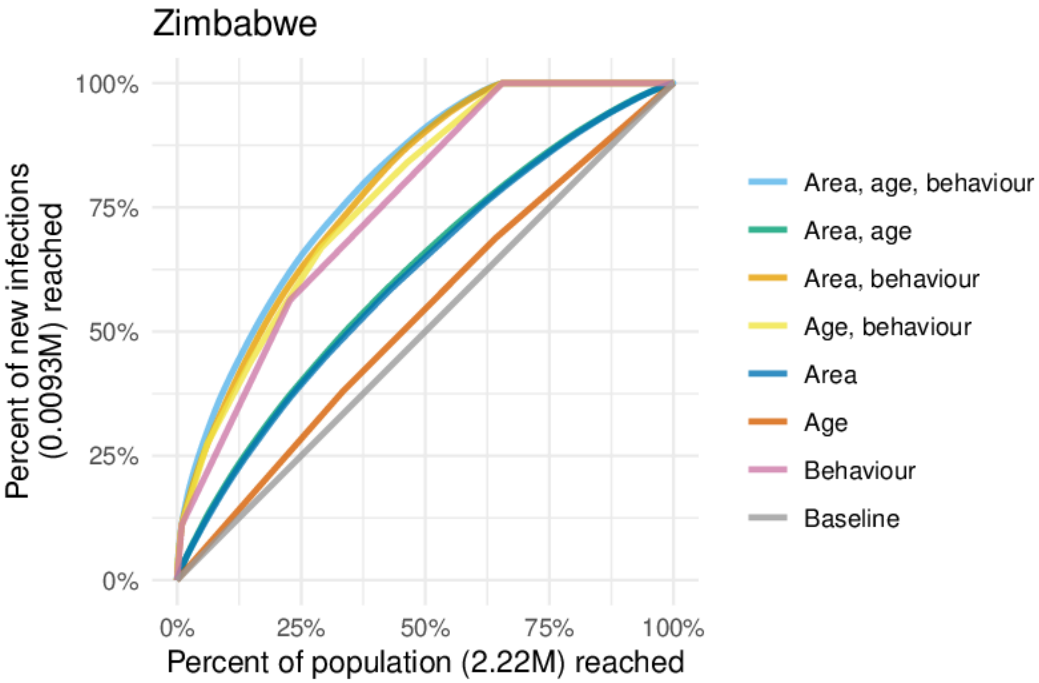

Figure BD: Percentage of expected new infections reached taking a variety of risk stratification approaches against the percentage of at risk population reached in Zimbabwe.
